# A Natural Language Processing Algorithm for Classifying Suicidal Behaviors in Alzheimer’s Disease and Related Dementia Patients: Development and Validation Using Electronic Health Records Data

**DOI:** 10.1101/2023.07.21.23292976

**Authors:** Kimia Zandbiglari, Hamid Reza Hasanzadeh, Pareeta Kotecha, Ruba Sajdeya, Amie J Goodin, Tianze Jiao, Farzana I Adiba, Mamoun T. Mardini, Jiang Bian, Masoud Rouhizadeh

## Abstract

This study aimed to develop a natural language processing algorithm (NLP) using machine learning (ML) and Deep Learning (DL) techniques to identify and classify documentation of suicidal behaviors in patients with Alzheimer’s disease and related dementia (ADRD). We utilized MIMIC-III and MIMIC-IV datasets and identified ADRD patients and subsequently those with suicide ideation using relevant International Classification of Diseases (ICD) codes. We used cosine similarity with ScAN (Suicide Attempt and Ideation Events Dataset) to calculate semantic similarity scores of ScAN with extracted notes from MIMIC for the clinical notes. The notes were sorted based on these scores, and manual review and categorization into eight suicidal behavior categories were performed. The data were further analyzed using conventional ML and DL models, with manual annotation as a reference. The tested classifiers achieved classification results close to human performance with up to 98% precision and 98% recall of suicidal ideation in the ADRD patient population. Our NLP model effectively reproduced human annotation of suicidal ideation within the MIMIC dataset. These results establish a foundation for identifying and categorizing documentation related to suicidal ideation within ADRD population, contributing to the advancement of NLP techniques in healthcare for extracting and classifying clinical concepts, particularly focusing on suicidal ideation among patients with ADRD. Our study showcased the capability of a robust NLP algorithm to accurately identify and classify documentation of suicidal behaviors in ADRD patients.

## Background and Significance

Suicide is a significant public health concern, with the crucial challenge being the identification of individuals at risk, in order to reduce suicide rates. Concurrently, there has been an increase in suicide rates, accounting for over 45,900 deaths in 2020 alone [1,2]. The Census Bureau report suggests that by 2030, the United States will see a significant rise in the older adult population [3]. Notably, suicide rates among older adults are highest, with the Baby Boomer generation particularly susceptible, as illustrated in Figure 1. This showcases suicide rates by age groups according to the Center for Disease and Control (CDC)[1,4]. This combined surge in both the older adult population and suicide rates across all age groups elevates suicide to an urgent public health issue.

**Figure 1:**
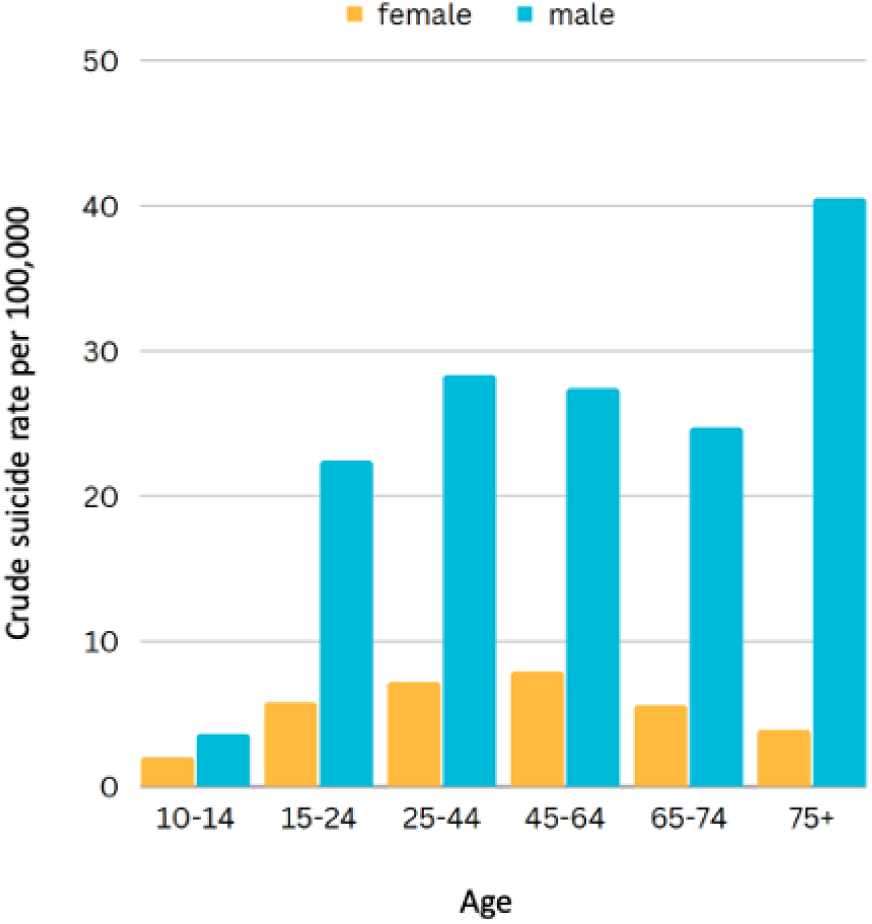
Suicide rates by age group in male and female population in 2020 according to CDC [1]

Alzheimer’s disease and related forms of dementia are among the top fears for an aging population [5,6]. Post-diagnosis, these conditions often evoke feelings of loss, anger, and confusion, which can be devastating for some individuals, potentially leading to suicidal tendencies [7,8]. Research indicates that the risk of suicide tends to increase with age both in the United States and globally [4,6,9,10]. Specifically, in 2018, the suicide rate among adults over 65 was 17.36 per 100,000, outpacing the rate in the general population (14.21 per 100,000) [4]. Further, studies highlight that the risk is most potent within the first year following an Alzheimer’s Disease and Related Dementias (AD/ADRD) diagnosis, with an alarming 54% increase in suicide rates compared to the general population. The risk peaks at a 243% increase in the first 90 days post-diagnosis, which could likely be attributed to a combination of negative feelings and risk factors resulting from the diagnosis, and this risk varies within the AD/ADRD population [6].

Certain risk factors, such as mental or substance use disorders and pain conditions, are significantly heightened in the AD/ADRD population. Individuals with pre-existing mental disorders experience more than triple the suicide rates compared to those with AD/ADRD alone. Given the escalating suicide rates in the US, the increasing AD/ADRD population, and the aging of the Baby Boomer generation, it is a public health priority to address this. A critical objective for healthcare systems is identifying those at the highest risk [6].

While clinical assessments can provide valuable insights into patients’ mental states, they are subjective and can be difficult to interpret accurately. Manual review of clinical notes is a time-consuming task that is susceptible to human error, irreproducibility, and unfeasibility for large-sized health databases, thus complicating comprehensive suicide risk assessment [11]. One source of inaccuracy in manual chart review originates from the varying levels of training and expertise among chart reviewers, which could introduce bias into the chart reviewing outcome. This is due to the fact that reviewers have not undergone uniform training and may not be adhering to a standardized guideline during the review process [12].

To tackle these issues, there is an escalating need for objective and efficient methods to identify patients at risk for suicide using clinical notes. Natural Language Processing (NLP) emerges as a powerful tool to extract medical concepts related to suicidal behaviors from unstructured clinical narratives, enabling healthcare providers to make more informed intervention decisions and enhance patient outcomes [13]. However, there are currently no NLP algorithms available to identify and categorize documentation of suicidal behavior in the ADRD population.

To ensure dependable results, it’s crucial to establish a solid annotation guideline that secures its quality. This research undertakes a comprehensive approach by incorporating guidelines from reliable sources such as the Food and Drug Administration (FDA) [14], the World Health Organization (WHO) [15], and the Medline Plus Medical Encyclopedia [16]. By leveraging these trusted guidelines, the research aims to achieve the utmost level of accuracy in defining suicidal behaviors, mental disorders, and overdoses. This careful selection of guidelines augments the reliability of the annotation phase, thus ensuring the overall quality of the annotations.

This research focuses on developing a comprehensive and dependable pipeline by incorporating multi-label classification. By using these techniques, the study aims to achieve simultaneous classification, providing a more robust and accurate approach. This allows for a more extensive coverage of information and boosts the reliability of the overall pipeline. The research specifically centers on utilizing clinical notes to identify patients at risk for suicide, with particular attention on individuals diagnosed with AD/ADRD.

## Materials and Methodology

### Study cohort and EHR-narrative preprocessing phase

For this study, we utilized the Medical Information Mart for Intensive Care (MIMIC) databases: MIMIC-III and MIMIC-IV, which are widely used, open-access databases that offer valuable resources for medical research and healthcare analytics. MIMIC databases comprise de-identified electronic health records (EHRs) of patients admitted to the intensive care units (ICUs) at a large academic medical center in Boston, Massachusetts, United States. MIMIC-IV, the latest version of the database, builds on the success of MIMIC-III, offering an expanded collection of de-identified EHR data from adult and neonatal ICUs. This enhancement broadens the potential for research in critical care and allows for investigations into a wider range of clinical questions [17]. Both MIMIC-III and MIMIC-IV have proven instrumental in advancing medical research and healthcare analytics by providing researchers with rich and diverse datasets that can be employed for various studies [17,18].

In the context of extracting patients with ADRD diagnoses from the MIMIC-III and MIMIC-IV databases, we employed a specific set of International Classification of Disease (ICD) codes [19] related to ADRD. These codes, covering both ICD-9 and ICD-10, were utilized to identify patients with ADRD diagnoses within the databases. Through these codes, we were able to extract a subset of patients with documented ADRD diagnoses from the extensive dataset. Further, within this extracted group of patients, our focus was on identifying instances of suicide-related information. To achieve this, we used approximately 1,500 ICD codes associated with suicide. Spanning both ICD-9 and ICD-10 code sets, these allowed us to identify patients within the ADRD population who had documented instances related to suicide. These data and diagnosis codes provided invaluable information for us to predict and categorize the risk of suicide among these patients. The relevant ICD codes are available in Appendices A and B.

In a recently published paper, researchers introduced ScAN (Suicide Attempt and Ideation Events Dataset), a comprehensive dataset that is publicly available. It encompasses 12,759 electronic health record (EHR) notes and includes 19,960 unique annotations, focusing specifically on suicide attempts and ideation events [20]. We used cosine similarity measure to calculate the similarity of sentences among patients diagnosed with ADRD who experienced a suicidal ideation or event, using the data from MIMIC-III and MIMIC-IV datasets with the sentences available in ScAN. Cosine similarity is a measure employed in semantic models to determine the similarity between two vectors representing textual data. It calculates the cosine of the angle between the vectors, where a value of 1 indicates perfect similarity and 0 indicates no similarity. This measure is commonly used in tasks like document similarity, information retrieval, and recommendation systems [21]. Using cosine similarity, we extracted 334,228 snippets from 11,267 notes of 267 identified patients. These snippets were then sorted based on their similarity scores, which ranged from 0.9967 to 0.1211.

### Annotation Process

To maintain consistency and accuracy in our annotations, we established written guidelines, provided relevant examples, and trained expert annotators to adhere to these guidelines when labeling the extracted notes. The annotation process involved two rounds of manual labeling, with eight key properties assessed for each matching snippet to ensure accurate and consistent assignment of labels. Table 1 presents these key properties and their respective definitions.

**Table 1:**
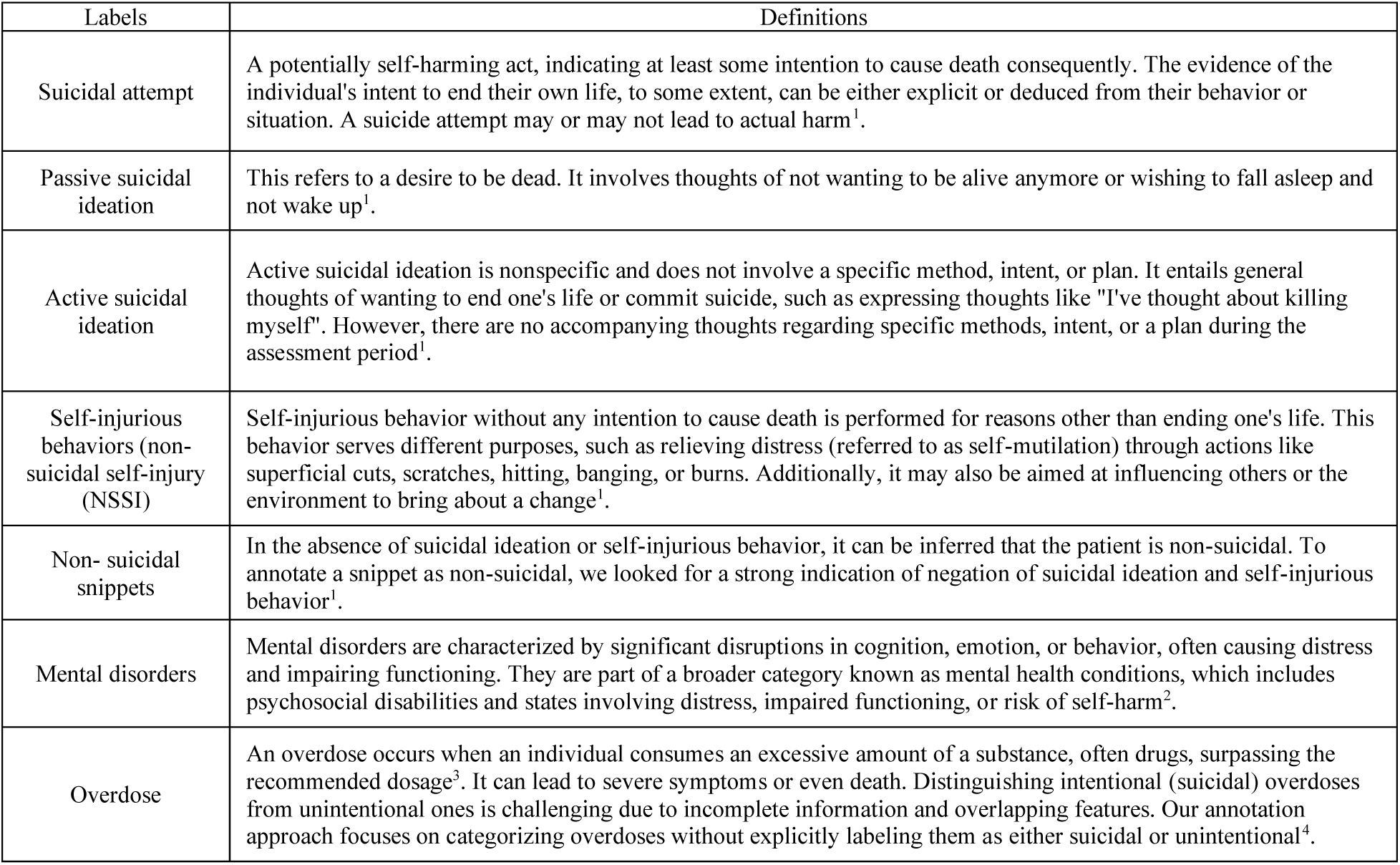
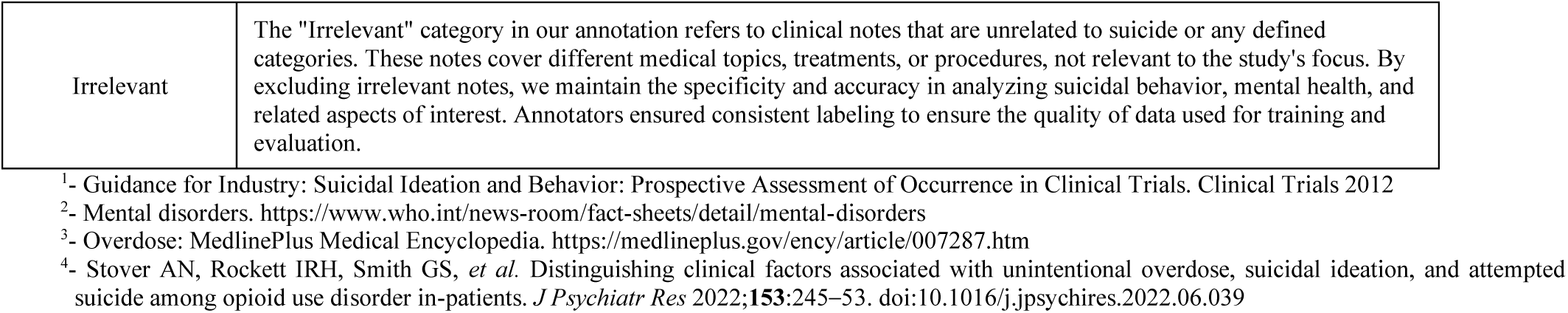
Key properties (labels) and definitions for suicidal behavior used for the annotation of clinical notes.

In the first round of annotation, we meticulously reviewed and labeled 150 note snippets, ensuring dual assessment of each snippet by two reviewers. Through the analysis of Inter-Rater Reliability (IRR) [22], we observed a remarkable overall agreement of 93.3%. Any discrepancies among the reviewers were addressed and resolved through consensus, with reviewers engaging in discussions to clarify their rationales, aiming to reach a mutually agreeable label that aligns with the guidelines.

This collaborative process ultimately resulted in a unanimous agreement of 100%. Following this round, we made refinements to the annotation guidelines by further clarifying the definitions of each property.

In the subsequent round of annotation, we conducted double reviews for 430 notes, which comprised 10% of the total snippets. The IRR analysis for this round exhibited an even higher agreement level of 97.4%. Similar to the initial round, any differences in opinion among the annotators were addressed through discussions and consensus, once again achieving a 100% agreement rate.

### Proposed models

During our snippet review process, we observed that certain snippets could be classified under multiple defined key properties. To ensure that all pertinent information from the snippets was captured, we assigned labels based on the criteria met by each snippet. Consequently, some snippets ended up with more than one label, leading us to adopt a multi-label classification approach. This approach facilitates a more comprehensive and accurate representation of the diverse properties exhibited by the snippets.

Multi-label classification (MLC) is a classification technique in machine learning that finds numerous applications in a variety of real-world scenarios. MLC enables the assignment of multiple non-mutually exclusive labels to each sample. One commonly used approach within MLC is the Binary Relevance (BR), first introduced by Gonçalves and Quaresma in 2003 [23]. BR operates by training a multi-label classifier using a loss function that treats each class independently. The simplicity and widespread acceptance of BR within the field largely contribute to its popularity [24].

### Classification

In the context of ADRD, we undertook the task of classifying instances of suicide ideation and attempts. To accomplish this, we utilized the annotated data snippets to train or otherwise fine-tune and evaluate a diverse range of machine learning (ML) and deep learning (DL) based classification models. Our objective was to develop automated methodologies capable of accurately identifying and classifying instances of suicide ideation and attempts in individuals with ADRD. By harnessing ML and DL techniques, our aim was to enhance the efficiency and reliability of detecting and documenting suicidal tendencies within the ADRD population.

### Conventional Machine Learning (ML)

To investigate suicide ideation and attempts in individuals diagnosed with ADRD, we utilized a variety of conventional machine learning (ML) classification models. Our analysis incorporated logistic regression (LR) with L1 and L2 regularization [25] and support vector machines (SVM) with different kernels, such as linear and radial basis function (RBF) [26]. We also included decision tree and random forest algorithms in our study on suicide ideation and attempts among individuals with ADRD.

We leveraged the Python-based pysbd library for sentence boundary detection [27], segmentation, lemmatization, and part-of-speech (POS) tagging. These processes helped enhance our classification by providing valuable linguistic information through assigning grammatical labels to words. Such linguistic insights can be used as features in machine learning models, capturing syntactic patterns and relevant linguistic context. For instance, in sentiment analysis, discerning whether a word functions as an adjective or an adverb can offer significant clues about its conveyed sentiment.

Combining POS tags with other textual features like word frequency or n-grams allows for the creation of more informative representations, leading to improved accuracy and robustness of the classification process, especially in tasks where the linguistic structure of the text holds vital importance [28]. Moreover, we employed the TF-IDF technique with a minimum document frequency of 1 and n-gram ranges from 1 to 5 to capture important features [29].

During the preprocessing stage, we removed all the stop words from the notes, following the list provided by the Natural Language Toolkit (NLTK) [30,31]. We extracted word n-gram sequences using the scikit-learn library in Python. The resulting word n-gram sequences were weighted using the TF-IDF approach, which measures term importance based on their frequency and scarcity across documents. Additionally, L2 regularization was applied to logistic regression to prevent overfitting by penalizing large coefficients.

In our analysis, we benchmarked multiple machine learning techniques and preprocessing methods to classify instances of suicide ideation and attempts in individuals with ADRD. We leveraged the n-gram approach to capture the sequential information in the text by considering word sequences of different lengths. To enhance the quality of our data, we performed part-of-speech tagging to assign grammatical labels to words and utilized lemmatization to reduce words to their base form.

Furthermore, we removed stop words as they lack significant contextual information. By combining these techniques with logistic regression and SVM algorithms using specific kernels, we were able to effectively classify and investigate suicide ideation and attempts in individuals with ADRD.

In our logistic regression model, we employed both L1 and L2 regularization techniques to control overfitting and improve generalization. We will report the better result obtained from either L1 or L2 regularization, based on their impact on the model’s accuracy and stability. L1 regularization encourages sparsity by shrinking some coefficients to zero, while L2 regularization imposes a penalty on the squared magnitude of coefficients. By comparing the outcomes of both regularization methods, we can determine which one yields superior results in terms of model performance [32].

Linear SVMs were used to identify the hyperplane that best separates training observations with a maximum margin [26,33].

### Deep Models

Transformer-based pre-trained models have significantly advanced text classification tasks, particularly in cases where labeled data is scarce. These language models, extensively trained on vast quantities of information, offer a deep contextual understanding. By fine-tuning them with a limited amount of problem-specific data, they can produce impressive results [34]. For our classification task, we utilized BERT-Base uncased [35], RoBERTa [36], and ClinicalBERT [37].

BERT-Base uncased, RoBERTa, and ClinicalBERT are variants of the BERT model, a potent language representation model. BERT-Base uncased is trained on a large corpus of uncased text data and can be applied to various natural language processing tasks like text classification and named entity recognition. RoBERTa enhances the BERT architecture and training methods, introducing modifications such as larger batch sizes and more data for training, thereby improving performance on downstream NLP tasks. Conversely, ClinicalBERT is specifically trained on clinical text data like electronic health records and medical literature. This enables it to capture the nuances and specialized terminology of medical text, making it especially valuable for healthcare-related NLP tasks, such as clinical text classification and information extraction [35–37].

To maintain consistency across all transformer models, we employed HuggingFace’s transformers library [38] in conjunction with a GPU-enabled PyTorch [39] framework. To ensure identical train and test sets for each model, we randomly split the dataset. Each model was loaded and utilized directly, with 80% of the data serving as the training set and the remaining 20% as the test set.

Accommodating different transformer models may require variations in tokenization encodings and attention mask utilization. Nevertheless, the transformers library provided by HuggingFace streamlines the implementation process for each model. During training, we reserved 10% of our training inputs as a validation set to monitor our classifier’s performance. We used the “stratify” parameter to ensure that the validation set did not contain any unseen labels. If any labels appeared only once in the dataset, we included them in the training set for appropriate stratification. We also created PyTorch data loaders to load the data for training and prediction. We retained the initial layers of the models as they learn general features, focusing on fine-tuning only the final layers. The input training data was tokenized and fed into the models for fine-tuning. The fine-tuned models were then used to classify the test set. For training the models, we used a mini-batch size of 32 and a learning rate of 2e-5. The optimization process involved the AdamW optimizer, with the loss parameter set to Binary Cross Entropy with Logits [40].

To evaluate the training performance over 30 epochs, we employed micro F1 accuracy for validation. This method includes extracting the logits from the model’s output, passing them through a sigmoid function to generate outputs between 0 and 1, and setting a threshold of 0.50 to create predictions. We compared these predictions to the true labels to calculate accuracy.

## Results

From a randomly selected sample of 1647 notes pertaining to 260 patients diagnosed with ADRD and suicide based on the ICD code search in the MIMIC dataset, we annotated 4364 clinical notes for the presence of suicide ideation and attempts. To maximize the usefulness of the data and train a more comprehensive model, we categorized the clinical notes as non-suicidal, self-injurious behaviors (also known as non-suicidal self-injury (NSSI)), overdose, and various mental disorders including anxiety, depression, and bipolar disorder. Out of the annotated snippets, 2225 were deemed irrelevant, leaving us with 2139 snippets annotated with pertinent labels. The frequency and percentage of occurrence for each label are detailed in Table 2.

**Table 2:**
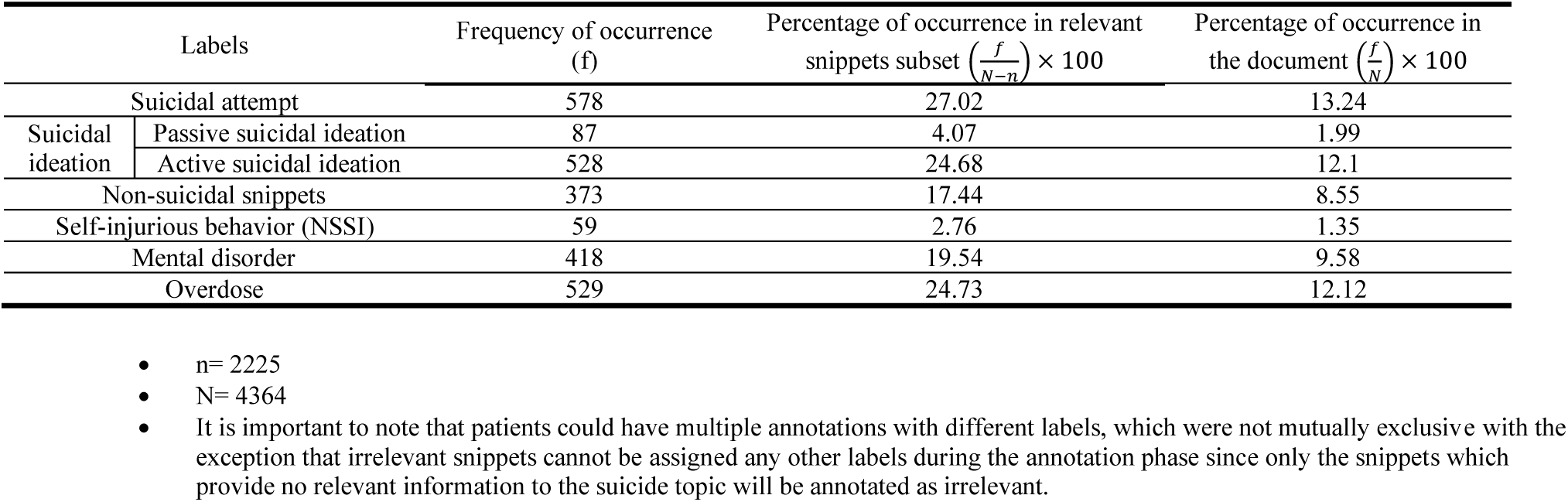
The frequency and percentage of occurrence of each label in relevant subset and the document

### Analysis and findings of models’ performance

Deep learning classifiers consistently outperformed conventional machine learning classifiers, achieving notably higher accuracy and precision. Among the deep learning models, Clinical BERT exhibited the best performance, securing a precision, recall, and F-1 score of 98% as a micro average, and a precision of 98%, recall (R) of 96%, and an F1-score (F) of 97% as a macro average. Comparatively, among conventional machine learning classifiers, Linear Support Vector Machine (L-SVM) offered the best overall performance with a precision of 79%, recall of 78%, and an F1-score of 79% in its micro average. In its macro average, it achieved a precision of 75%, recall of 61%, and an F1-score of 66%. Among the tested SVM kernels, the linear kernel exhibited superior performance for our specific classification task. The performances of Logistic Regression (LR) and Decision Tree (DT) models were marginally lower.

Table 3 outlines the precision, recall, and F1-score for both conventional machine learning and deep learning models across various labels. It provides a comprehensive overview of each method’s performance metrics, enabling a direct comparison of their effectiveness. The micro average considers the total count of true positives, false positives, and false negatives across all labels, providing a holistic view of the model’s classification ability, treating all labels equally. Conversely, the macro average calculates the average performance of the models on a per-label basis, disregarding class imbalance. This measure facilitates a balanced evaluation, assigning equal importance to each label, and emphasizing the models’ ability to generalize across different categories. The weighted average accounts for class distribution and offers a performance measure that accommodates label imbalance. It ascribes weights to each label based on its prevalence in the dataset, ensuring the evaluation reflects the model’s performance in a more representative manner.

**Table 3:**
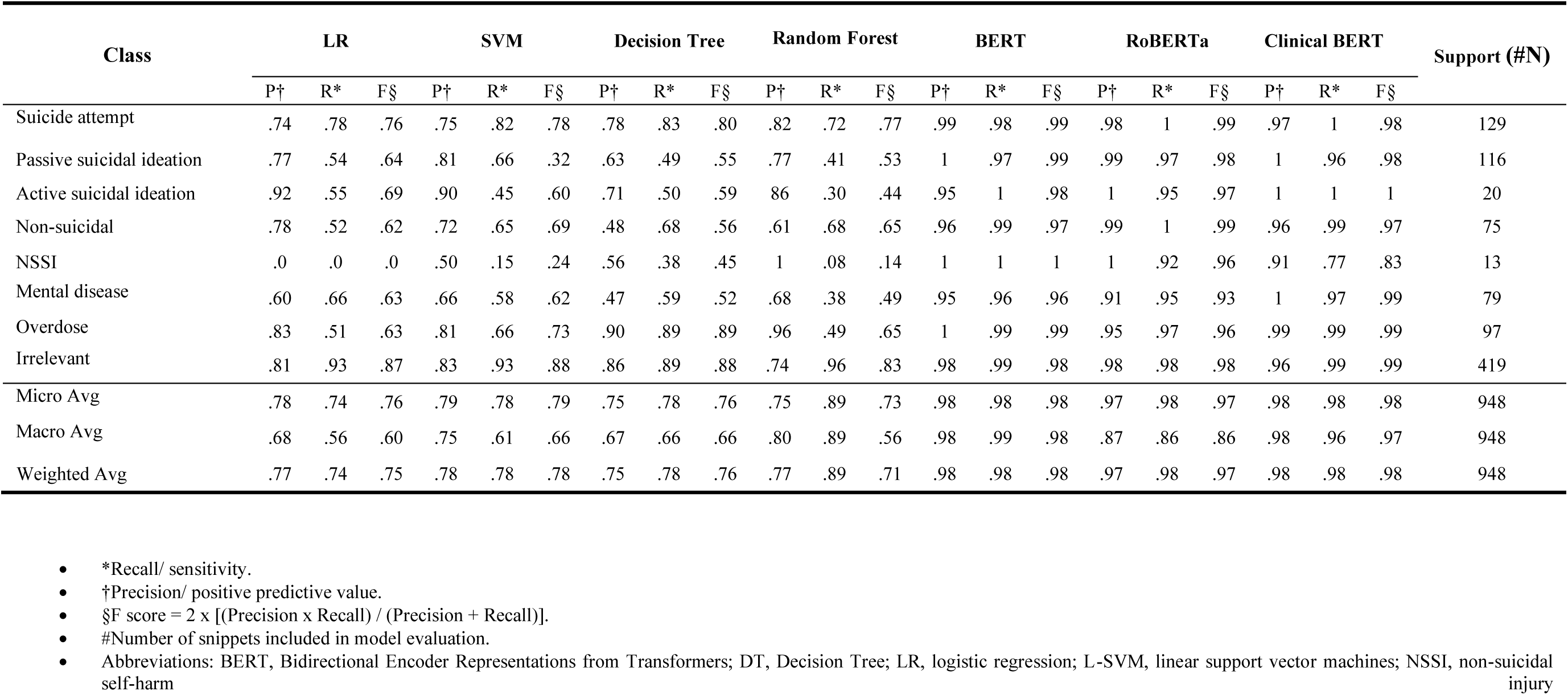
Performance of machine learning, and deep learning classifiers of ADRD patient population extracted from MIMIC dataset documented in unstructured narrative clinical notes.

## Discussion

In this study, we focused on utilizing the MIMIC dataset to extract a cohort of patients with Alzheimer’s Disease and Related Dementias (ADRD) who exhibited suicidal behavior based on specific ICD codes related to suicide. We employed three deep models: BERT, RoBERTa, and Clinical BERT, along with four conventional machine learning algorithms: logistic regression, SVM, random forest, and decision tree. Among the deep learning methods, ClinicalBERT demonstrated the best results, while among the conventional models we explored, the support vector machine exhibited promising results, albeit with slightly lower accuracy compared to the deep learning models. This underlines the potential of conventional machine learning techniques in classifying clinical text data related to suicidal behavior, especially when deploying deep learning models may not be feasible due to resource constraints or specific application requirements.

The use of deep learning models, particularly ClinicalBERT, allowed us to effectively classify the clinical notes into the given categories. It is not surprising that these deep learning models outperformed conventional machine learning, given their capacity to leverage pretraining on extensive textual data, even if from different domains. The weights obtained during pretraining enable these models to capture and understand complex patterns in the text, contributing to their high accuracy in classifying the data. Overall, the excellent performance of the multilabel text classification models in identifying suicide ideation and attempt in ADRD patients based on clinical text data underscores the potential of natural language processing techniques for early detection and intervention in mental health conditions among this population.

A notable limitation of this study was the lack of consideration for the temporal aspect of suicidal behavior. We acknowledge that some patients may have had suicidal ideation in the past but were not suicidal at the time of note collection. Future research should consider the temporal context and incorporate period assertion to enhance the accuracy of identifying individuals with current suicidal ideation or behavior.

In the annotation phase, we classified all “overdose” events under the “overdose” category. We acknowledge the limitations in determining the intent behind these occurrences based solely on the snippet content. This approach ensures that our classification models are trained and evaluated based on the presence of an overdose event, avoiding potentially inaccurate or subjective determinations of suicidal intent or unintentionality.

The tool developed in this study has valuable applications in clinical practice for assessing and managing ADRD patients with suicidal behavior. By automatically classifying clinical notes based on suicidal behavior categories, it identifies high-risk patients, expediting early interventions and saving time for clinicians and caregivers. Additionally, it provides insights into the complexity of suicidal behavior, informing treatment decisions and personalized care plans. However, this tool should be used to complement clinical judgment, not replace it. Integrating this tool could enhance suicide risk assessment in ADRD patients, leveraging natural language processing and machine learning for targeted interventions and improved patient care.

Further research and validation using larger and more diverse datasets are necessary to enhance the generalizability and applicability of these models in clinical practice.

## Conclusion

In conclusion, this study utilized the MIMIC dataset to extract a cohort of patients with Alzheimer’s Disease and Related Dementias (ADRD) who exhibited suicidal behavior based on specific ICD codes related to suicide. The clinical notes of these patients were annotated with eight labels, and various models were explored to infer these labels in a multi-label setting. The deep learning models, particularly ClinicalBERT, demonstrated superior performance in accurately classifying the clinical notes based on the given categories. The use of deep learning models, including BERT, RoBERTa, and Clinical BERT, proved effective in capturing contextual information and semantic relationships within the text data. Additionally, conventional machine learning techniques, such as the support vector machine, showed promise in classifying clinical text data related to suicidal behavior.

Overall, the findings underscore the potential of natural language processing techniques, particularly deep learning models, for early detection and intervention in mental health conditions among ADRD patients. They also highlight the value of conventional machine learning approaches in scenarios where resources may be limited. Future research should focus on integrating temporal information, validating the models on larger and diverse datasets, and examining their applicability in clinical practice to enhance generalizability and practical implementation.

## Data Availability

The data used in this article cannot be publicly shared due to restrictions imposed by the MIMIC dataset's policy. Although the dataset is publicly available, access to it requires the completion of specific training and obtaining a certificate. In compliance with the MIMIC dataset policy, we are unable to share the data utilized in this research.

## ACKNOWLEDGMENTS

Research reported in this publication was supported by the University of Florida Department of Pharmaceutical Outcomes & Policy, AI in the Health Sciences Initiative, College of Pharmacy. The content is solely the responsibility of the authors and does not necessarily represent the official views of the National Institutes of Health.

## DATA AVAILABILITY STATEMENT

The data used in this article cannot be publicly shared due to restrictions imposed by the MIMIC dataset’s policy. Although the dataset is publicly available, access to it requires completion of specific trainings and obtaining a certificate. In compliance with the MIMIC dataset policy, we are unable to share the data utilized in this research.

