## Appendix A for "A Natural Language Processing Algorithm for Classifying Suicidal Behaviors in Alzheimer’s Disease and Related Dementia Patients: Development and Validation Using Electronic Health Records Data"

| RelevantDiagnosis | Description | DiagnosisCohort2 | disease1 | | Textbox19 | Diagnosis | ICDDescription |
| --- | --- | --- | --- | --- | --- | --- | --- |
| Suicide-related event | Has the patient had a suicide-related event? | Suicide | Suicide | ICD10 | | R45.851 | Suicidal ideations |
| Suicide-related event | Has the patient had a suicide-related event? | Suicide | Suicide | ICD10 | | T14.91 | Suicide Attempt |
| Suicide-related event | Has the patient had a suicide-related event? | Suicide | Suicide | ICD10 | | T14.91XA | Suicide attempt, initial encounter |
| Suicide-related event | Has the patient had a suicide-related event? | Suicide | Suicide | ICD10 | | T14.91XD | Suicide attempt, subsequent encounter |
| Suicide-related event | Has the patient had a suicide-related event? | Suicide | Suicide | ICD10 | | T14.91XS | Suicide attempt, sequela |
| Suicide-related event | Has the patient had a suicide-related event? | Suicide | Suicide | ICD10 | | T36.0X2A | Poisoning by penicillins, intentional self-harm, initial encounter |
| Suicide-related event | Has the patient had a suicide-related event? | Suicide | Suicide | ICD10 | | T36.0X2D | Poisoning by penicillins, intentional self-harm, subsequent encounter |
| Suicide-related event | Has the patient had a suicide-related event? | Suicide | Suicide | ICD10 | | T36.0X2S | Poisoning by penicillins, intentional self-harm, sequela |
| Suicide-related event | Has the patient had a suicide-related event? | Suicide | Suicide | ICD10 | | T36.1X2A | Poisoning by cephalosporins and other beta-lactam antibiotics, intentional self-harm, initial encounter |
| Suicide-related event | Has the patient had a suicide-related event? | Suicide | Suicide | ICD10 | | T36.1X2D | Poisoning by cephalosporins and other beta-lactam antibiotics, intentional self-harm, subsequent encounter |
| Suicide-related event | Has the patient had a suicide-related event? | Suicide | Suicide | ICD10 | | T36.1X2S | Poisoning by cephalosporins and other beta-lactam antibiotics, intentional self-harm, sequela |
| Suicide-related event | Has the patient had a suicide-related event? | Suicide | Suicide | ICD10 | | T36.2X2A | Poisoning by chloramphenicol group, intentional self-harm, initial encounter |
| Suicide-related event | Has the patient had a suicide-related event? | Suicide | Suicide | ICD10 | | T36.2X2D | Poisoning by chloramphenicol group, intentional self-harm, subsequent encounter |
| Suicide-related event | Has the patient had a suicide-related event? | Suicide | Suicide | ICD10 | | T36.2X2S | Poisoning by chloramphenicol group, intentional self-harm, sequela |
| Suicide-related event | Has the patient had a suicide-related event? | Suicide | Suicide | ICD10 | | T36.3X2A | Poisoning by macrolides, intentional self-harm, initial encounter |
| Suicide-related event | Has the patient had a suicide-related event? | Suicide | Suicide | ICD10 | | T36.3X2D | Poisoning by macrolides, intentional self-harm, subsequent encounter |
| Suicide-related event | Has the patient had a suicide-related event? | Suicide | Suicide | ICD10 | | T36.3X2S | Poisoning by macrolides, intentional self-harm, sequela |
| Suicide-related event | Has the patient had a suicide-related event? | Suicide | Suicide | ICD10 | | T36.4X2A | Poisoning by tetracyclines, intentional self-harm, initial encounter |
| Suicide-related event | Has the patient had a suicide-related event? | Suicide | Suicide | ICD10 | | T36.4X2D | Poisoning by tetracyclines, intentional self-harm, subsequent encounter |
| Suicide-related event | Has the patient had a suicide-related event? | Suicide | Suicide | ICD10 | | T36.4X2S | Poisoning by tetracyclines, intentional self-harm, sequela |
| Suicide-related event | Has the patient had a suicide-related event? | Suicide | Suicide | ICD10 | | T36.5X2A | Poisoning by aminoglycosides, intentional self-harm, initial encounter |
| Suicide-related event | Has the patient had a suicide-related event? | Suicide | Suicide | ICD10 | | T36.5X2D | Poisoning by aminoglycosides, intentional self-harm, subsequent encounter |
| Suicide-related event | Has the patient had a suicide-related event? | Suicide | Suicide | ICD10 | | T36.5X2S | Poisoning by aminoglycosides, intentional self-harm, sequela |
| Suicide-related event | Has the patient had a suicide-related event? | Suicide | Suicide | ICD10 | | T36.6X2A | Poisoning by rifampicins, intentional self-harm, initial encounter |
| Suicide-related event | Has the patient had a suicide-related event? | Suicide | Suicide | ICD10 | | T36.6X2D | Poisoning by rifampicins, intentional self-harm, subsequent encounter |
| Suicide-related event | Has the patient had a suicide-related event? | Suicide | Suicide | ICD10 | | T36.6X2S | Poisoning by rifampicins, intentional self-harm, sequela |
| Suicide-related event | Has the patient had a suicide-related event? | Suicide | Suicide | ICD10 | | T36.7X2A | Poisoning by antifungal antibiotics, systemically used, intentional self-harm, initial encounter |
| Suicide-related event | Has the patient had a suicide-related event? | Suicide | Suicide | ICD10 | | T36.7X2D | Poisoning by antifungal antibiotics, systemically used, intentional self-harm, subsequent encounter |
| Suicide-related event | Has the patient had a suicide-related event? | Suicide | Suicide | ICD10 | | T36.7X2S | Poisoning by antifungal antibiotics, systemically used, intentional self-harm, sequela |
| Suicide-related event | Has the patient had a suicide-related event? | Suicide | Suicide | ICD10 | | T36.8X2A | Poisoning by other systemic antibiotics, intentional self-harm, initial encounter |
| Suicide-related event | Has the patient had a suicide-related event? | Suicide | Suicide | ICD10 | | T36.8X2D | Poisoning by other systemic antibiotics, intentional self-harm, subsequent encounter |
| Suicide-related event | Has the patient had a suicide-related event? | Suicide | Suicide | ICD10 | | T36.8X2S | Poisoning by other systemic antibiotics, intentional self-harm, sequela |
| Suicide-related event | Has the patient had a suicide-related event? | Suicide | Suicide | ICD10 | | T36.92XA | Poisoning by unspecified systemic antibiotic, intentional self-harm, initial encounter |
| Suicide-related event | Has the patient had a suicide-related event? | Suicide | Suicide | ICD10 | | T36.92XD | Poisoning by unspecified systemic antibiotic, intentional self-harm, subsequent encounter |
| Suicide-related event | Has the patient had a suicide-related event? | Suicide | Suicide | ICD10 | | T36.92XS | Poisoning by unspecified systemic antibiotic, intentional self-harm, sequela |
| Suicide-related event | Has the patient had a suicide-related event? | Suicide | Suicide | ICD10 | | T37.0X2A | Poisoning by sulfonamides, intentional self-harm, initial encounter |
| Suicide-related event | Has the patient had a suicide-related event? | Suicide | Suicide | ICD10 | | T37.0X2D | Poisoning by sulfonamides, intentional self-harm, subsequent encounter |
| Suicide-related event | Has the patient had a suicide-related event? | Suicide | Suicide | ICD10 | | T37.0X2S | Poisoning by sulfonamides, intentional self-harm, sequela |
| Suicide-related event | Has the patient had a suicide-related event? | Suicide | Suicide | ICD10 | | T37.1X2A | Poisoning by antimycobacterial drugs, intentional self-harm, initial encounter |
| Suicide-related event | Has the patient had a suicide-related event? | Suicide | Suicide | ICD10 | | T37.1X2D | Poisoning by antimycobacterial drugs, intentional self-harm, subsequent encounter |
| Suicide-related event | Has the patient had a suicide-related event? | Suicide | Suicide | ICD10 | | T37.1X2S | Poisoning by antimycobacterial drugs, intentional self-harm, sequela |
| Suicide-related event | Has the patient had a suicide-related event? | Suicide | Suicide | ICD10 | | T37.2X2A | Poisoning by antimalarials and drugs acting on other blood protozoa, intentional self-harm, initial encounter |
| Suicide-related event | Has the patient had a suicide-related event? | Suicide | Suicide | ICD10 | | T37.2X2D | Poisoning by antimalarials and drugs acting on other blood protozoa, intentional self-harm, subsequent encounter |
| Suicide-related event | Has the patient had a suicide-related event? | Suicide | Suicide | ICD10 | | T37.2X2S | Poisoning by antimalarials and drugs acting on other blood protozoa, intentional self-harm, sequela |
| Suicide-related event | Has the patient had a suicide-related event? | Suicide | Suicide | ICD10 | | T37.3X2A | Poisoning by other antiprotozoal drugs, intentional self-harm, initial encounter |
| Suicide-related event | Has the patient had a suicide-related event? | Suicide | Suicide | ICD10 | | T37.3X2D | Poisoning by other antiprotozoal drugs, intentional self-harm, subsequent encounter |
| Suicide-related event | Has the patient had a suicide-related event? | Suicide | Suicide | ICD10 | | T37.3X2S | Poisoning by other antiprotozoal drugs, intentional self-harm, sequela |
| Suicide-related event | Has the patient had a suicide-related event? | Suicide | Suicide | ICD10 | | T37.4X2A | Poisoning by anthelminthics, intentional self-harm, initial encounter |
| Suicide-related event | Has the patient had a suicide-related event? | Suicide | Suicide | ICD10 | | T37.4X2D | Poisoning by anthelminthics, intentional self-harm, subsequent encounter |
| Suicide-related event | Has the patient had a suicide-related event? | Suicide | Suicide | ICD10 | | T37.4X2S | Poisoning by anthelminthics, intentional self-harm, sequela |
| Suicide-related event | Has the patient had a suicide-related event? | Suicide | Suicide | ICD10 | | T37.5X2A | Poisoning by antiviral drugs, intentional self-harm, initial encounter |
| Suicide-related event | Has the patient had a suicide-related event? | Suicide | Suicide | ICD10 | | T37.5X2D | Poisoning by antiviral drugs, intentional self-harm, subsequent encounter |
| Suicide-related event | Has the patient had a suicide-related event? | Suicide | Suicide | ICD10 | | T37.5X2S | Poisoning by antiviral drugs, intentional self-harm, sequela |
| Suicide-related event | Has the patient had a suicide-related event? | Suicide | Suicide | ICD10 | | T37.8X2A | Poisoning by other specified systemic anti-infectives and antiparasitics, intentional self-harm, initial encounter |
| Suicide-related event | Has the patient had a suicide-related event? | Suicide | Suicide | ICD10 | | T37.8X2D | Poisoning by other specified systemic anti-infectives and antiparasitics, intentional self-harm, subsequent encounter |
| Suicide-related event | Has the patient had a suicide-related event? | Suicide | Suicide | ICD10 | | T37.8X2S | Poisoning by other specified systemic anti-infectives and antiparasitics, intentional self-harm, sequela |
| Suicide-related event | Has the patient had a suicide-related event? | Suicide | Suicide | ICD10 | | T37.92XA | Poisoning by unspecified systemic anti-infective and antiparasitics, intentional self-harm, initial encounter |
| Suicide-related event | Has the patient had a suicide-related event? | Suicide | Suicide | ICD10 | | T37.92XD | Poisoning by unspecified systemic anti-infective and antiparasitics, intentional self-harm, subsequent encounter |
| Suicide-related event | Has the patient had a suicide-related event? | Suicide | Suicide | ICD10 | | T37.92XS | Poisoning by unspecified systemic anti-infective and antiparasitics, intentional self-harm, sequela |
| Suicide-related event | Has the patient had a suicide-related event? | Suicide | Suicide | ICD10 | | T38.0X2A | Poisoning by glucocorticoids and synthetic analogues, intentional self-harm, initial encounter |
| Suicide-related event | Has the patient had a suicide-related event? | Suicide | Suicide | ICD10 | | T38.0X2D | Poisoning by glucocorticoids and synthetic analogues, intentional self-harm, subsequent encounter |
| Suicide-related event | Has the patient had a suicide-related event? | Suicide | Suicide | ICD10 | | T38.0X2S | Poisoning by glucocorticoids and synthetic analogues, intentional self-harm, sequela |
| Suicide-related event | Has the patient had a suicide-related event? | Suicide | Suicide | ICD10 | | T38.1X2A | Poisoning by thyroid hormones and substitutes, intentional self-harm, initial encounter |
| Suicide-related event | Has the patient had a suicide-related event? | Suicide | Suicide | ICD10 | | T38.1X2D | Poisoning by thyroid hormones and substitutes, intentional self-harm, subsequent encounter |
| Suicide-related event | Has the patient had a suicide-related event? | Suicide | Suicide | ICD10 | | T38.1X2S | Poisoning by thyroid hormones and substitutes, intentional self-harm, sequela |
| Suicide-related event | Has the patient had a suicide-related event? | Suicide | Suicide | ICD10 | | T38.2X2A | Poisoning by antithyroid drugs, intentional self-harm, initial encounter |
| Suicide-related event | Has the patient had a suicide-related event? | Suicide | Suicide | ICD10 | | T38.2X2D | Poisoning by antithyroid drugs, intentional self-harm, subsequent encounter |
| Suicide-related event | Has the patient had a suicide-related event? | Suicide | Suicide | ICD10 | | T38.2X2S | Poisoning by antithyroid drugs, intentional self-harm, sequela |
| Suicide-related event | Has the patient had a suicide-related event? | Suicide | Suicide | ICD10 | | T38.3X2A | Poisoning by insulin and oral hypoglycemic [antidiabetic] drugs, intentional self-harm, initial encounter |
| Suicide-related event | Has the patient had a suicide-related event? | Suicide | Suicide | ICD10 | | T38.3X2D | Poisoning by insulin and oral hypoglycemic [antidiabetic] drugs, intentional self-harm, subsequent encounter |
| Suicide-related event | Has the patient had a suicide-related event? | Suicide | Suicide | ICD10 | | T38.3X2S | Poisoning by insulin and oral hypoglycemic [antidiabetic] drugs, intentional self-harm, sequela |
| Suicide-related event | Has the patient had a suicide-related event? | Suicide | Suicide | ICD10 | | T38.4X2A | Poisoning by oral contraceptives, intentional self-harm, initial encounter |
| Suicide-related event | Has the patient had a suicide-related event? | Suicide | Suicide | ICD10 | | T38.4X2D | Poisoning by oral contraceptives, intentional self-harm, subsequent encounter |
| Suicide-related event | Has the patient had a suicide-related event? | Suicide | Suicide | ICD10 | | T38.4X2S | Poisoning by oral contraceptives, intentional self-harm, sequela |
| Suicide-related event | Has the patient had a suicide-related event? | Suicide | Suicide | ICD10 | | T38.5X2A | Poisoning by other estrogens and progestogens, intentional self-harm, initial encounter |
| Suicide-related event | Has the patient had a suicide-related event? | Suicide | Suicide | ICD10 | | T38.5X2D | Poisoning by other estrogens and progestogens, intentional self-harm, subsequent encounter |
| Suicide-related event | Has the patient had a suicide-related event? | Suicide | Suicide | ICD10 | | T38.5X2S | Poisoning by other estrogens and progestogens, intentional self-harm, sequela |
| Suicide-related event | Has the patient had a suicide-related event? | Suicide | Suicide | ICD10 | | T38.6X2A | Poisoning by antigonadotrophins, antiestrogens, antiandrogens, not elsewhere classified, intentional self-harm, initial encounter |
| Suicide-related event | Has the patient had a suicide-related event? | Suicide | Suicide | ICD10 | | T38.6X2D | Poisoning by antigonadotrophins, antiestrogens, antiandrogens, not elsewhere classified, intentional self-harm, subsequent encounter |
| Suicide-related event | Has the patient had a suicide-related event? | Suicide | Suicide | ICD10 | | T38.6X2S | Poisoning by antigonadotrophins, antiestrogens, antiandrogens, not elsewhere classified, intentional self-harm, sequela |
| Suicide-related event | Has the patient had a suicide-related event? | Suicide | Suicide | ICD10 | | T38.7X2A | Poisoning by androgens and anabolic congeners, intentional self-harm, initial encounter |
| Suicide-related event | Has the patient had a suicide-related event? | Suicide | Suicide | ICD10 | | T38.7X2D | Poisoning by androgens and anabolic congeners, intentional self-harm, subsequent encounter |
| Suicide-related event | Has the patient had a suicide-related event? | Suicide | Suicide | ICD10 | | T38.7X2S | Poisoning by androgens and anabolic congeners, intentional self-harm, sequela |
| Suicide-related event | Has the patient had a suicide-related event? | Suicide | Suicide | ICD10 | | T38.802A | Poisoning by unspecified hormones and synthetic substitutes, intentional self-harm, initial encounter |
| Suicide-related event | Has the patient had a suicide-related event? | Suicide | Suicide | ICD10 | | T38.802D | Poisoning by unspecified hormones and synthetic substitutes, intentional self-harm, subsequent encounter |
| Suicide-related event | Has the patient had a suicide-related event? | Suicide | Suicide | ICD10 | | T38.802S | Poisoning by unspecified hormones and synthetic substitutes, intentional self-harm, sequela |
| Suicide-related event | Has the patient had a suicide-related event? | Suicide | Suicide | ICD10 | | T38.812A | Poisoning by anterior pituitary [adenohypophyseal] hormones, intentional self-harm, initial encounter |
| Suicide-related event | Has the patient had a suicide-related event? | Suicide | Suicide | ICD10 | | T38.812D | Poisoning by anterior pituitary [adenohypophyseal] hormones, intentional self-harm, subsequent encounter |
| Suicide-related event | Has the patient had a suicide-related event? | Suicide | Suicide | ICD10 | | T38.812S | Poisoning by anterior pituitary [adenohypophyseal] hormones, intentional self-harm, sequela |
| Suicide-related event | Has the patient had a suicide-related event? | Suicide | Suicide | ICD10 | | T38.892A | Poisoning by other hormones and synthetic substitutes, intentional self-harm, initial encounter |
| Suicide-related event | Has the patient had a suicide-related event? | Suicide | Suicide | ICD10 | | T38.892D | Poisoning by other hormones and synthetic substitutes, intentional self-harm, subsequent encounter |
| Suicide-related event | Has the patient had a suicide-related event? | Suicide | Suicide | ICD10 | | T38.892S | Poisoning by other hormones and synthetic substitutes, intentional self-harm, sequela |
| Suicide-related event | Has the patient had a suicide-related event? | Suicide | Suicide | ICD10 | | T38.902A | Poisoning by unspecified hormone antagonists, intentional self-harm, initial encounter |
| Suicide-related event | Has the patient had a suicide-related event? | Suicide | Suicide | ICD10 | | T38.902D | Poisoning by unspecified hormone antagonists, intentional self-harm, subsequent encounter |
| Suicide-related event | Has the patient had a suicide-related event? | Suicide | Suicide | ICD10 | | T38.902S | Poisoning by unspecified hormone antagonists, intentional self-harm, sequela |
| Suicide-related event | Has the patient had a suicide-related event? | Suicide | Suicide | ICD10 | | T38.992A | Poisoning by other hormone antagonists, intentional self-harm, initial encounter |
| Suicide-related event | Has the patient had a suicide-related event? | Suicide | Suicide | ICD10 | | T38.992D | Poisoning by other hormone antagonists, intentional self-harm, subsequent encounter |
| Suicide-related event | Has the patient had a suicide-related event? | Suicide | Suicide | ICD10 | | T38.992S | Poisoning by other hormone antagonists, intentional self-harm, sequela |
| Suicide-related event | Has the patient had a suicide-related event? | Suicide | Suicide | ICD10 | | T39.012A | Poisoning by aspirin, intentional self-harm, initial encounter |
| Suicide-related event | Has the patient had a suicide-related event? | Suicide | Suicide | ICD10 | | T39.012D | Poisoning by aspirin, intentional self-harm, subsequent encounter |
| Suicide-related event | Has the patient had a suicide-related event? | Suicide | Suicide | ICD10 | | T39.012S | Poisoning by aspirin, intentional self-harm, sequela |
| Suicide-related event | Has the patient had a suicide-related event? | Suicide | Suicide | ICD10 | | T39.092A | Poisoning by salicylates, intentional self-harm, initial encounter |
| Suicide-related event | Has the patient had a suicide-related event? | Suicide | Suicide | ICD10 | | T39.092D | Poisoning by salicylates, intentional self-harm, subsequent encounter |
| Suicide-related event | Has the patient had a suicide-related event? | Suicide | Suicide | ICD10 | | T39.092S | Poisoning by salicylates, intentional self-harm, sequela |
| Suicide-related event | Has the patient had a suicide-related event? | Suicide | Suicide | ICD10 | | T39.1X2A | Poisoning by 4-Aminophenol derivatives, intentional self-harm, initial encounter |
| Suicide-related event | Has the patient had a suicide-related event? | Suicide | Suicide | ICD10 | | T39.1X2D | Poisoning by 4-Aminophenol derivatives, intentional self-harm, subsequent encounter |
| Suicide-related event | Has the patient had a suicide-related event? | Suicide | Suicide | ICD10 | | T39.1X2S | Poisoning by 4-Aminophenol derivatives, intentional self-harm, sequela |
| Suicide-related event | Has the patient had a suicide-related event? | Suicide | Suicide | ICD10 | | T39.2X2A | Poisoning by pyrazolone derivatives, intentional self-harm, initial encounter |
| Suicide-related event | Has the patient had a suicide-related event? | Suicide | Suicide | ICD10 | | T39.2X2D | Poisoning by pyrazolone derivatives, intentional self-harm, subsequent encounter |
| Suicide-related event | Has the patient had a suicide-related event? | Suicide | Suicide | ICD10 | | T39.2X2S | Poisoning by pyrazolone derivatives, intentional self-harm, sequela |
| Suicide-related event | Has the patient had a suicide-related event? | Suicide | Suicide | ICD10 | | T39.312A | Poisoning by propionic acid derivatives, intentional self-harm, initial encounter |
| Suicide-related event | Has the patient had a suicide-related event? | Suicide | Suicide | ICD10 | | T39.312D | Poisoning by propionic acid derivatives, intentional self-harm, subsequent encounter |
| Suicide-related event | Has the patient had a suicide-related event? | Suicide | Suicide | ICD10 | | T39.312S | Poisoning by propionic acid derivatives, intentional self-harm, sequela |
| Suicide-related event | Has the patient had a suicide-related event? | Suicide | Suicide | ICD10 | | T39.392A | Poisoning by other nonsteroidal anti-inflammatory drugs [NSAID], intentional self-harm, initial encounter |
| Suicide-related event | Has the patient had a suicide-related event? | Suicide | Suicide | ICD10 | | T39.392D | Poisoning by other nonsteroidal anti-inflammatory drugs [NSAID], intentional self-harm, subsequent encounter |
| Suicide-related event | Has the patient had a suicide-related event? | Suicide | Suicide | ICD10 | | T39.392S | Poisoning by other nonsteroidal anti-inflammatory drugs [NSAID], intentional self-harm, sequela |
| Suicide-related event | Has the patient had a suicide-related event? | Suicide | Suicide | ICD10 | | T39.4X2A | Poisoning by antirheumatics, not elsewhere classified, intentional self-harm, initial encounter |
| Suicide-related event | Has the patient had a suicide-related event? | Suicide | Suicide | ICD10 | | T39.4X2D | Poisoning by antirheumatics, not elsewhere classified, intentional self-harm, subsequent encounter |
| Suicide-related event | Has the patient had a suicide-related event? | Suicide | Suicide | ICD10 | | T39.4X2S | Poisoning by antirheumatics, not elsewhere classified, intentional self-harm, sequela |
| Suicide-related event | Has the patient had a suicide-related event? | Suicide | Suicide | ICD10 | | T39.8X2A | Poisoning by other nonopioid analgesics and antipyretics, not elsewhere classified, intentional self-harm, initial encounter |
| Suicide-related event | Has the patient had a suicide-related event? | Suicide | Suicide | ICD10 | | T39.8X2D | Poisoning by other nonopioid analgesics and antipyretics, not elsewhere classified, intentional self-harm, subsequent encounter |
| Suicide-related event | Has the patient had a suicide-related event? | Suicide | Suicide | ICD10 | | T39.8X2S | Poisoning by other nonopioid analgesics and antipyretics, not elsewhere classified, intentional self-harm, sequela |
| Suicide-related event | Has the patient had a suicide-related event? | Suicide | Suicide | ICD10 | | T39.92XA | Poisoning by unspecified nonopioid analgesic, antipyretic and antirheumatic, intentional self-harm, initial encounter |
| Suicide-related event | Has the patient had a suicide-related event? | Suicide | Suicide | ICD10 | | T39.92XD | Poisoning by unspecified nonopioid analgesic, antipyretic and antirheumatic, intentional self-harm, subsequent encounter |
| Suicide-related event | Has the patient had a suicide-related event? | Suicide | Suicide | ICD10 | | T39.92XS | Poisoning by unspecified nonopioid analgesic, antipyretic and antirheumatic, intentional self-harm, sequela |
| Suicide-related event | Has the patient had a suicide-related event? | Suicide | Suicide | ICD10 | | T40.0X2A | Poisoning by opium, intentional self-harm, initial encounter |
| Suicide-related event | Has the patient had a suicide-related event? | Suicide | Suicide | ICD10 | | T40.0X2D | Poisoning by opium, intentional self-harm, subsequent encounter |
| Suicide-related event | Has the patient had a suicide-related event? | Suicide | Suicide | ICD10 | | T40.0X2S | Poisoning by opium, intentional self-harm, sequela |
| Suicide-related event | Has the patient had a suicide-related event? | Suicide | Suicide | ICD10 | | T40.1X2A | Poisoning by heroin, intentional self-harm, initial encounter |
| Suicide-related event | Has the patient had a suicide-related event? | Suicide | Suicide | ICD10 | | T40.1X2D | Poisoning by heroin, intentional self-harm, subsequent encounter |
| Suicide-related event | Has the patient had a suicide-related event? | Suicide | Suicide | ICD10 | | T40.1X2S | Poisoning by heroin, intentional self-harm, sequela |
| Suicide-related event | Has the patient had a suicide-related event? | Suicide | Suicide | ICD10 | | T40.2X2A | Poisoning by other opioids, intentional self-harm, initial encounter |
| Suicide-related event | Has the patient had a suicide-related event? | Suicide | Suicide | ICD10 | | T40.2X2D | Poisoning by other opioids, intentional self-harm, subsequent encounter |
| Suicide-related event | Has the patient had a suicide-related event? | Suicide | Suicide | ICD10 | | T40.2X2S | Poisoning by other opioids, intentional self-harm, sequela |
| Suicide-related event | Has the patient had a suicide-related event? | Suicide | Suicide | ICD10 | | T40.3X2A | Poisoning by methadone, intentional self-harm, initial encounter |
| Suicide-related event | Has the patient had a suicide-related event? | Suicide | Suicide | ICD10 | | T40.3X2D | Poisoning by methadone, intentional self-harm, subsequent encounter |
| Suicide-related event | Has the patient had a suicide-related event? | Suicide | Suicide | ICD10 | | T40.3X2S | Poisoning by methadone, intentional self-harm, sequela |
| Suicide-related event | Has the patient had a suicide-related event? | Suicide | Suicide | ICD10 | | T40.4X2A | Poisoning by other synthetic narcotics, intentional self-harm, initial encounter |
| Suicide-related event | Has the patient had a suicide-related event? | Suicide | Suicide | ICD10 | | T40.4X2D | Poisoning by other synthetic narcotics, intentional self-harm, subsequent encounter |
| Suicide-related event | Has the patient had a suicide-related event? | Suicide | Suicide | ICD10 | | T40.4X2S | Poisoning by other synthetic narcotics, intentional self-harm, sequela |
| Suicide-related event | Has the patient had a suicide-related event? | Suicide | Suicide | ICD10 | | T40.5X2A | Poisoning by cocaine, intentional self-harm, initial encounter |
| Suicide-related event | Has the patient had a suicide-related event? | Suicide | Suicide | ICD10 | | T40.5X2D | Poisoning by cocaine, intentional self-harm, subsequent encounter |
| Suicide-related event | Has the patient had a suicide-related event? | Suicide | Suicide | ICD10 | | T40.5X2S | Poisoning by cocaine, intentional self-harm, sequela |
| Suicide-related event | Has the patient had a suicide-related event? | Suicide | Suicide | ICD10 | | T40.602A | Poisoning by unspecified narcotics, intentional self-harm, initial encounter |
| Suicide-related event | Has the patient had a suicide-related event? | Suicide | Suicide | ICD10 | | T40.602D | Poisoning by unspecified narcotics, intentional self-harm, subsequent encounter |
| Suicide-related event | Has the patient had a suicide-related event? | Suicide | Suicide | ICD10 | | T40.602S | Poisoning by unspecified narcotics, intentional self-harm, sequela |
| Suicide-related event | Has the patient had a suicide-related event? | Suicide | Suicide | ICD10 | | T40.692A | Poisoning by other narcotics, intentional self-harm, initial encounter |
| Suicide-related event | Has the patient had a suicide-related event? | Suicide | Suicide | ICD10 | | T40.692D | Poisoning by other narcotics, intentional self-harm, subsequent encounter |
| Suicide-related event | Has the patient had a suicide-related event? | Suicide | Suicide | ICD10 | | T40.692S | Poisoning by other narcotics, intentional self-harm, sequela |
| Suicide-related event | Has the patient had a suicide-related event? | Suicide | Suicide | ICD10 | | T40.7X2A | Poisoning by cannabis (derivatives), intentional self-harm, initial encounter |
| Suicide-related event | Has the patient had a suicide-related event? | Suicide | Suicide | ICD10 | | T40.7X2D | Poisoning by cannabis (derivatives), intentional self-harm, subsequent encounter |
| Suicide-related event | Has the patient had a suicide-related event? | Suicide | Suicide | ICD10 | | T40.7X2S | Poisoning by cannabis (derivatives), intentional self-harm, sequela |
| Suicide-related event | Has the patient had a suicide-related event? | Suicide | Suicide | ICD10 | | T40.8X2A | Poisoning by lysergide [LSD], intentional self-harm, initial encounter |
| Suicide-related event | Has the patient had a suicide-related event? | Suicide | Suicide | ICD10 | | T40.8X2D | Poisoning by lysergide [LSD], intentional self-harm, subsequent encounter |
| Suicide-related event | Has the patient had a suicide-related event? | Suicide | Suicide | ICD10 | | T40.8X2S | Poisoning by lysergide [LSD], intentional self-harm, sequela |
| Suicide-related event | Has the patient had a suicide-related event? | Suicide | Suicide | ICD10 | | T40.902A | Poisoning by unspecified psychodysleptics [hallucinogens], intentional self-harm, initial encounter |
| Suicide-related event | Has the patient had a suicide-related event? | Suicide | Suicide | ICD10 | | T40.902D | Poisoning by unspecified psychodysleptics [hallucinogens], intentional self-harm, subsequent encounter |
| Suicide-related event | Has the patient had a suicide-related event? | Suicide | Suicide | ICD10 | | T40.902S | Poisoning by unspecified psychodysleptics [hallucinogens], intentional self-harm, sequela |
| Suicide-related event | Has the patient had a suicide-related event? | Suicide | Suicide | ICD10 | | T40.992A | Poisoning by other psychodysleptics [hallucinogens], intentional self-harm, initial encounter |
| Suicide-related event | Has the patient had a suicide-related event? | Suicide | Suicide | ICD10 | | T40.992D | Poisoning by other psychodysleptics [hallucinogens], intentional self-harm, subsequent encounter |
| Suicide-related event | Has the patient had a suicide-related event? | Suicide | Suicide | ICD10 | | T40.992S | Poisoning by other psychodysleptics [hallucinogens], intentional self-harm, sequela |
| Suicide-related event | Has the patient had a suicide-related event? | Suicide | Suicide | ICD10 | | T41.0X2A | Poisoning by inhaled anesthetics, intentional self-harm, initial encounter |
| Suicide-related event | Has the patient had a suicide-related event? | Suicide | Suicide | ICD10 | | T41.0X2D | Poisoning by inhaled anesthetics, intentional self-harm, subsequent encounter |
| Suicide-related event | Has the patient had a suicide-related event? | Suicide | Suicide | ICD10 | | T41.0X2S | Poisoning by inhaled anesthetics, intentional self-harm, sequela |
| Suicide-related event | Has the patient had a suicide-related event? | Suicide | Suicide | ICD10 | | T41.1X2A | Poisoning by intravenous anesthetics, intentional self-harm, initial encounter |
| Suicide-related event | Has the patient had a suicide-related event? | Suicide | Suicide | ICD10 | | T41.1X2D | Poisoning by intravenous anesthetics, intentional self-harm, subsequent encounter |
| Suicide-related event | Has the patient had a suicide-related event? | Suicide | Suicide | ICD10 | | T41.1X2S | Poisoning by intravenous anesthetics, intentional self-harm, sequela |
| Suicide-related event | Has the patient had a suicide-related event? | Suicide | Suicide | ICD10 | | T41.202A | Poisoning by unspecified general anesthetics, intentional self-harm, initial encounter |
| Suicide-related event | Has the patient had a suicide-related event? | Suicide | Suicide | ICD10 | | T41.202D | Poisoning by unspecified general anesthetics, intentional self-harm, subsequent encounter |
| Suicide-related event | Has the patient had a suicide-related event? | Suicide | Suicide | ICD10 | | T41.202S | Poisoning by unspecified general anesthetics, intentional self-harm, sequela |
| Suicide-related event | Has the patient had a suicide-related event? | Suicide | Suicide | ICD10 | | T41.292A | Poisoning by other general anesthetics, intentional self-harm, initial encounter |
| Suicide-related event | Has the patient had a suicide-related event? | Suicide | Suicide | ICD10 | | T41.292D | Poisoning by other general anesthetics, intentional self-harm, subsequent encounter |
| Suicide-related event | Has the patient had a suicide-related event? | Suicide | Suicide | ICD10 | | T41.292S | Poisoning by other general anesthetics, intentional self-harm, sequela |
| Suicide-related event | Has the patient had a suicide-related event? | Suicide | Suicide | ICD10 | | T41.3X2A | Poisoning by local anesthetics, intentional self-harm, initial encounter |
| Suicide-related event | Has the patient had a suicide-related event? | Suicide | Suicide | ICD10 | | T41.3X2D | Poisoning by local anesthetics, intentional self-harm, subsequent encounter |
| Suicide-related event | Has the patient had a suicide-related event? | Suicide | Suicide | ICD10 | | T41.3X2S | Poisoning by local anesthetics, intentional self-harm, sequela |
| Suicide-related event | Has the patient had a suicide-related event? | Suicide | Suicide | ICD10 | | T41.42XA | Poisoning by unspecified anesthetic, intentional self-harm, initial encounter |
| Suicide-related event | Has the patient had a suicide-related event? | Suicide | Suicide | ICD10 | | T41.42XD | Poisoning by unspecified anesthetic, intentional self-harm, subsequent encounter |
| Suicide-related event | Has the patient had a suicide-related event? | Suicide | Suicide | ICD10 | | T41.42XS | Poisoning by unspecified anesthetic, intentional self-harm, sequela |
| Suicide-related event | Has the patient had a suicide-related event? | Suicide | Suicide | ICD10 | | T41.5X2A | Poisoning by therapeutic gases, intentional self-harm, initial encounter |
| Suicide-related event | Has the patient had a suicide-related event? | Suicide | Suicide | ICD10 | | T41.5X2D | Poisoning by therapeutic gases, intentional self-harm, subsequent encounter |
| Suicide-related event | Has the patient had a suicide-related event? | Suicide | Suicide | ICD10 | | T41.5X2S | Poisoning by therapeutic gases, intentional self-harm, sequela |
| Suicide-related event | Has the patient had a suicide-related event? | Suicide | Suicide | ICD10 | | T42.0X2A | Poisoning by hydantoin derivatives, intentional self-harm, initial encounter |
| Suicide-related event | Has the patient had a suicide-related event? | Suicide | Suicide | ICD10 | | T42.0X2D | Poisoning by hydantoin derivatives, intentional self-harm, subsequent encounter |
| Suicide-related event | Has the patient had a suicide-related event? | Suicide | Suicide | ICD10 | | T42.0X2S | Poisoning by hydantoin derivatives, intentional self-harm, sequela |
| Suicide-related event | Has the patient had a suicide-related event? | Suicide | Suicide | ICD10 | | T42.1X2A | Poisoning by iminostilbenes, intentional self-harm, initial encounter |
| Suicide-related event | Has the patient had a suicide-related event? | Suicide | Suicide | ICD10 | | T42.1X2D | Poisoning by iminostilbenes, intentional self-harm, subsequent encounter |
| Suicide-related event | Has the patient had a suicide-related event? | Suicide | Suicide | ICD10 | | T42.1X2S | Poisoning by iminostilbenes, intentional self-harm, sequela |
| Suicide-related event | Has the patient had a suicide-related event? | Suicide | Suicide | ICD10 | | T42.2X2A | Poisoning by succinimides and oxazolidinediones, intentional self-harm, initial encounter |
| Suicide-related event | Has the patient had a suicide-related event? | Suicide | Suicide | ICD10 | | T42.2X2D | Poisoning by succinimides and oxazolidinediones, intentional self-harm, subsequent encounter |
| Suicide-related event | Has the patient had a suicide-related event? | Suicide | Suicide | ICD10 | | T42.2X2S | Poisoning by succinimides and oxazolidinediones, intentional self-harm, sequela |
| Suicide-related event | Has the patient had a suicide-related event? | Suicide | Suicide | ICD10 | | T42.3X2A | Poisoning by barbiturates, intentional self-harm, initial encounter |
| Suicide-related event | Has the patient had a suicide-related event? | Suicide | Suicide | ICD10 | | T42.3X2D | Poisoning by barbiturates, intentional self-harm, subsequent encounter |
| Suicide-related event | Has the patient had a suicide-related event? | Suicide | Suicide | ICD10 | | T42.3X2S | Poisoning by barbiturates, intentional self-harm, sequela |
| Suicide-related event | Has the patient had a suicide-related event? | Suicide | Suicide | ICD10 | | T42.4X2A | Poisoning by benzodiazepines, intentional self-harm, initial encounter |
| Suicide-related event | Has the patient had a suicide-related event? | Suicide | Suicide | ICD10 | | T42.4X2D | Poisoning by benzodiazepines, intentional self-harm, subsequent encounter |
| Suicide-related event | Has the patient had a suicide-related event? | Suicide | Suicide | ICD10 | | T42.4X2S | Poisoning by benzodiazepines, intentional self-harm, sequela |
| Suicide-related event | Has the patient had a suicide-related event? | Suicide | Suicide | ICD10 | | T42.5X2A | Poisoning by mixed antiepileptics, intentional self-harm, initial encounter |
| Suicide-related event | Has the patient had a suicide-related event? | Suicide | Suicide | ICD10 | | T42.5X2D | Poisoning by mixed antiepileptics, intentional self-harm, subsequent encounter |
| Suicide-related event | Has the patient had a suicide-related event? | Suicide | Suicide | ICD10 | | T42.5X2S | Poisoning by mixed antiepileptics, intentional self-harm, sequela |
| Suicide-related event | Has the patient had a suicide-related event? | Suicide | Suicide | ICD10 | | T42.6X2A | Poisoning by other antiepileptic and sedative-hypnotic drugs, intentional self-harm, initial encounter |
| Suicide-related event | Has the patient had a suicide-related event? | Suicide | Suicide | ICD10 | | T42.6X2D | Poisoning by other antiepileptic and sedative-hypnotic drugs, intentional self-harm, subsequent encounter |
| Suicide-related event | Has the patient had a suicide-related event? | Suicide | Suicide | ICD10 | | T42.6X2S | Poisoning by other antiepileptic and sedative-hypnotic drugs, intentional self-harm, sequela |
| Suicide-related event | Has the patient had a suicide-related event? | Suicide | Suicide | ICD10 | | T42.72XA | Poisoning by unspecified antiepileptic and sedative-hypnotic drugs, intentional self-harm, initial encounter |
| Suicide-related event | Has the patient had a suicide-related event? | Suicide | Suicide | ICD10 | | T42.72XD | Poisoning by unspecified antiepileptic and sedative-hypnotic drugs, intentional self-harm, subsequent encounter |
| Suicide-related event | Has the patient had a suicide-related event? | Suicide | Suicide | ICD10 | | T42.72XS | Poisoning by unspecified antiepileptic and sedative-hypnotic drugs, intentional self-harm, sequela |
| Suicide-related event | Has the patient had a suicide-related event? | Suicide | Suicide | ICD10 | | T42.8X2A | Poisoning by antiparkinsonism drugs and other central muscle-tone depressants, intentional self-harm, initial encounter |
| Suicide-related event | Has the patient had a suicide-related event? | Suicide | Suicide | ICD10 | | T42.8X2D | Poisoning by antiparkinsonism drugs and other central muscle-tone depressants, intentional self-harm, subsequent encounter |
| Suicide-related event | Has the patient had a suicide-related event? | Suicide | Suicide | ICD10 | | T42.8X2S | Poisoning by antiparkinsonism drugs and other central muscle-tone depressants, intentional self-harm, sequela |
| Suicide-related event | Has the patient had a suicide-related event? | Suicide | Suicide | ICD10 | | T43.012A | Poisoning by tricyclic antidepressants, intentional self-harm, initial encounter |
| Suicide-related event | Has the patient had a suicide-related event? | Suicide | Suicide | ICD10 | | T43.012D | Poisoning by tricyclic antidepressants, intentional self-harm, subsequent encounter |
| Suicide-related event | Has the patient had a suicide-related event? | Suicide | Suicide | ICD10 | | T43.012S | Poisoning by tricyclic antidepressants, intentional self-harm, sequela |
| Suicide-related event | Has the patient had a suicide-related event? | Suicide | Suicide | ICD10 | | T43.022A | Poisoning by tetracyclic antidepressants, intentional self-harm, initial encounter |
| Suicide-related event | Has the patient had a suicide-related event? | Suicide | Suicide | ICD10 | | T43.022D | Poisoning by tetracyclic antidepressants, intentional self-harm, subsequent encounter |
| Suicide-related event | Has the patient had a suicide-related event? | Suicide | Suicide | ICD10 | | T43.022S | Poisoning by tetracyclic antidepressants, intentional self-harm, sequela |
| Suicide-related event | Has the patient had a suicide-related event? | Suicide | Suicide | ICD10 | | T43.1X2A | Poisoning by monoamine-oxidase-inhibitor antidepressants, intentional self-harm, initial encounter |
| Suicide-related event | Has the patient had a suicide-related event? | Suicide | Suicide | ICD10 | | T43.1X2D | Poisoning by monoamine-oxidase-inhibitor antidepressants, intentional self-harm, subsequent encounter |
| Suicide-related event | Has the patient had a suicide-related event? | Suicide | Suicide | ICD10 | | T43.1X2S | Poisoning by monoamine-oxidase-inhibitor antidepressants, intentional self-harm, sequela |
| Suicide-related event | Has the patient had a suicide-related event? | Suicide | Suicide | ICD10 | | T43.202A | Poisoning by unspecified antidepressants, intentional self-harm, initial encounter |
| Suicide-related event | Has the patient had a suicide-related event? | Suicide | Suicide | ICD10 | | T43.202D | Poisoning by unspecified antidepressants, intentional self-harm, subsequent encounter |
| Suicide-related event | Has the patient had a suicide-related event? | Suicide | Suicide | ICD10 | | T43.202S | Poisoning by unspecified antidepressants, intentional self-harm, sequela |
| Suicide-related event | Has the patient had a suicide-related event? | Suicide | Suicide | ICD10 | | T43.212A | Poisoning by selective serotonin and norepinephrine reuptake inhibitors, intentional self-harm, initial encounter |
| Suicide-related event | Has the patient had a suicide-related event? | Suicide | Suicide | ICD10 | | T43.212D | Poisoning by selective serotonin and norepinephrine reuptake inhibitors, intentional self-harm, subsequent encounter |
| Suicide-related event | Has the patient had a suicide-related event? | Suicide | Suicide | ICD10 | | T43.212S | Poisoning by selective serotonin and norepinephrine reuptake inhibitors, intentional self-harm, sequela |
| Suicide-related event | Has the patient had a suicide-related event? | Suicide | Suicide | ICD10 | | T43.222A | Poisoning by selective serotonin reuptake inhibitors, intentional self-harm, initial encounter |
| Suicide-related event | Has the patient had a suicide-related event? | Suicide | Suicide | ICD10 | | T43.222D | Poisoning by selective serotonin reuptake inhibitors, intentional self-harm, subsequent encounter |
| Suicide-related event | Has the patient had a suicide-related event? | Suicide | Suicide | ICD10 | | T43.222S | Poisoning by selective serotonin reuptake inhibitors, intentional self-harm, sequela |
| Suicide-related event | Has the patient had a suicide-related event? | Suicide | Suicide | ICD10 | | T43.292A | Poisoning by other antidepressants, intentional self-harm, initial encounter |
| Suicide-related event | Has the patient had a suicide-related event? | Suicide | Suicide | ICD10 | | T43.292D | Poisoning by other antidepressants, intentional self-harm, subsequent encounter |
| Suicide-related event | Has the patient had a suicide-related event? | Suicide | Suicide | ICD10 | | T43.292S | Poisoning by other antidepressants, intentional self-harm, sequela |
| Suicide-related event | Has the patient had a suicide-related event? | Suicide | Suicide | ICD10 | | T43.3X2A | Poisoning by phenothiazine antipsychotics and neuroleptics, intentional self-harm, initial encounter |
| Suicide-related event | Has the patient had a suicide-related event? | Suicide | Suicide | ICD10 | | T43.3X2D | Poisoning by phenothiazine antipsychotics and neuroleptics, intentional self-harm, subsequent encounter |
| Suicide-related event | Has the patient had a suicide-related event? | Suicide | Suicide | ICD10 | | T43.3X2S | Poisoning by phenothiazine antipsychotics and neuroleptics, intentional self-harm, sequela |
| Suicide-related event | Has the patient had a suicide-related event? | Suicide | Suicide | ICD10 | | T43.4X2A | Poisoning by butyrophenone and thiothixene neuroleptics, intentional self-harm, initial encounter |
| Suicide-related event | Has the patient had a suicide-related event? | Suicide | Suicide | ICD10 | | T43.4X2D | Poisoning by butyrophenone and thiothixene neuroleptics, intentional self-harm, subsequent encounter |
| Suicide-related event | Has the patient had a suicide-related event? | Suicide | Suicide | ICD10 | | T43.4X2S | Poisoning by butyrophenone and thiothixene neuroleptics, intentional self-harm, sequela |
| Suicide-related event | Has the patient had a suicide-related event? | Suicide | Suicide | ICD10 | | T43.502A | Poisoning by unspecified antipsychotics and neuroleptics, intentional self-harm, initial encounter |
| Suicide-related event | Has the patient had a suicide-related event? | Suicide | Suicide | ICD10 | | T43.502D | Poisoning by unspecified antipsychotics and neuroleptics, intentional self-harm, subsequent encounter |
| Suicide-related event | Has the patient had a suicide-related event? | Suicide | Suicide | ICD10 | | T43.502S | Poisoning by unspecified antipsychotics and neuroleptics, intentional self-harm, sequela |
| Suicide-related event | Has the patient had a suicide-related event? | Suicide | Suicide | ICD10 | | T43.592A | Poisoning by other antipsychotics and neuroleptics, intentional self-harm, initial encounter |
| Suicide-related event | Has the patient had a suicide-related event? | Suicide | Suicide | ICD10 | | T43.592D | Poisoning by other antipsychotics and neuroleptics, intentional self-harm, subsequent encounter |
| Suicide-related event | Has the patient had a suicide-related event? | Suicide | Suicide | ICD10 | | T43.592S | Poisoning by other antipsychotics and neuroleptics, intentional self-harm, sequela |
| Suicide-related event | Has the patient had a suicide-related event? | Suicide | Suicide | ICD10 | | T43.602A | Poisoning by unspecified psychostimulants, intentional self-harm, initial encounter |
| Suicide-related event | Has the patient had a suicide-related event? | Suicide | Suicide | ICD10 | | T43.602D | Poisoning by unspecified psychostimulants, intentional self-harm, subsequent encounter |
| Suicide-related event | Has the patient had a suicide-related event? | Suicide | Suicide | ICD10 | | T43.602S | Poisoning by unspecified psychostimulants, intentional self-harm, sequela |
| Suicide-related event | Has the patient had a suicide-related event? | Suicide | Suicide | ICD10 | | T43.612A | Poisoning by caffeine, intentional self-harm, initial encounter |
| Suicide-related event | Has the patient had a suicide-related event? | Suicide | Suicide | ICD10 | | T43.612D | Poisoning by caffeine, intentional self-harm, subsequent encounter |
| Suicide-related event | Has the patient had a suicide-related event? | Suicide | Suicide | ICD10 | | T43.612S | Poisoning by caffeine, intentional self-harm, sequela |
| Suicide-related event | Has the patient had a suicide-related event? | Suicide | Suicide | ICD10 | | T43.622A | Poisoning by amphetamines, intentional self-harm, initial encounter |
| Suicide-related event | Has the patient had a suicide-related event? | Suicide | Suicide | ICD10 | | T43.622D | Poisoning by amphetamines, intentional self-harm, subsequent encounter |
| Suicide-related event | Has the patient had a suicide-related event? | Suicide | Suicide | ICD10 | | T43.622S | Poisoning by amphetamines, intentional self-harm, sequela |
| Suicide-related event | Has the patient had a suicide-related event? | Suicide | Suicide | ICD10 | | T43.632A | Poisoning by methylphenidate, intentional self-harm, initial encounter |
| Suicide-related event | Has the patient had a suicide-related event? | Suicide | Suicide | ICD10 | | T43.632D | Poisoning by methylphenidate, intentional self-harm, subsequent encounter |
| Suicide-related event | Has the patient had a suicide-related event? | Suicide | Suicide | ICD10 | | T43.632S | Poisoning by methylphenidate, intentional self-harm, sequela |
| Suicide-related event | Has the patient had a suicide-related event? | Suicide | Suicide | ICD10 | | T43.642A | Poisoning by ecstasy, intentional self-harm, initial encounter |
| Suicide-related event | Has the patient had a suicide-related event? | Suicide | Suicide | ICD10 | | T43.642D | Poisoning by ecstasy, intentional self-harm, subsequent encounter |
| Suicide-related event | Has the patient had a suicide-related event? | Suicide | Suicide | ICD10 | | T43.642S | Poisoning by ecstasy, intentional self-harm, sequela |
| Suicide-related event | Has the patient had a suicide-related event? | Suicide | Suicide | ICD10 | | T43.692A | Poisoning by other psychostimulants, intentional self-harm, initial encounter |
| Suicide-related event | Has the patient had a suicide-related event? | Suicide | Suicide | ICD10 | | T43.692D | Poisoning by other psychostimulants, intentional self-harm, subsequent encounter |
| Suicide-related event | Has the patient had a suicide-related event? | Suicide | Suicide | ICD10 | | T43.692S | Poisoning by other psychostimulants, intentional self-harm, sequela |
| Suicide-related event | Has the patient had a suicide-related event? | Suicide | Suicide | ICD10 | | T43.8X2A | Poisoning by other psychotropic drugs, intentional self-harm, initial encounter |
| Suicide-related event | Has the patient had a suicide-related event? | Suicide | Suicide | ICD10 | | T43.8X2D | Poisoning by other psychotropic drugs, intentional self-harm, subsequent encounter |
| Suicide-related event | Has the patient had a suicide-related event? | Suicide | Suicide | ICD10 | | T43.8X2S | Poisoning by other psychotropic drugs, intentional self-harm, sequela |
| Suicide-related event | Has the patient had a suicide-related event? | Suicide | Suicide | ICD10 | | T43.92XA | Poisoning by unspecified psychotropic drug, intentional self-harm, initial encounter |
| Suicide-related event | Has the patient had a suicide-related event? | Suicide | Suicide | ICD10 | | T43.92XD | Poisoning by unspecified psychotropic drug, intentional self-harm, subsequent encounter |
| Suicide-related event | Has the patient had a suicide-related event? | Suicide | Suicide | ICD10 | | T43.92XS | Poisoning by unspecified psychotropic drug, intentional self-harm, sequela |
| Suicide-related event | Has the patient had a suicide-related event? | Suicide | Suicide | ICD10 | | T44.0X2A | Poisoning by anticholinesterase agents, intentional self-harm, initial encounter |
| Suicide-related event | Has the patient had a suicide-related event? | Suicide | Suicide | ICD10 | | T44.0X2D | Poisoning by anticholinesterase agents, intentional self-harm, subsequent encounter |
| Suicide-related event | Has the patient had a suicide-related event? | Suicide | Suicide | ICD10 | | T44.0X2S | Poisoning by anticholinesterase agents, intentional self-harm, sequela |
| Suicide-related event | Has the patient had a suicide-related event? | Suicide | Suicide | ICD10 | | T44.1X2A | Poisoning by other parasympathomimetics [cholinergics], intentional self-harm, initial encounter |
| Suicide-related event | Has the patient had a suicide-related event? | Suicide | Suicide | ICD10 | | T44.1X2D | Poisoning by other parasympathomimetics [cholinergics], intentional self-harm, subsequent encounter |
| Suicide-related event | Has the patient had a suicide-related event? | Suicide | Suicide | ICD10 | | T44.1X2S | Poisoning by other parasympathomimetics [cholinergics], intentional self-harm, sequela |
| Suicide-related event | Has the patient had a suicide-related event? | Suicide | Suicide | ICD10 | | T44.2X2A | Poisoning by ganglionic blocking drugs, intentional self-harm, initial encounter |
| Suicide-related event | Has the patient had a suicide-related event? | Suicide | Suicide | ICD10 | | T44.2X2D | Poisoning by ganglionic blocking drugs, intentional self-harm, subsequent encounter |
| Suicide-related event | Has the patient had a suicide-related event? | Suicide | Suicide | ICD10 | | T44.2X2S | Poisoning by ganglionic blocking drugs, intentional self-harm, sequela |
| Suicide-related event | Has the patient had a suicide-related event? | Suicide | Suicide | ICD10 | | T44.3X2A | Poisoning by other parasympatholytics [anticholinergics and antimuscarinics] and spasmolytics, intentional self-harm, initial encounter |
| Suicide-related event | Has the patient had a suicide-related event? | Suicide | Suicide | ICD10 | | T44.3X2D | Poisoning by other parasympatholytics [anticholinergics and antimuscarinics] and spasmolytics, intentional self-harm, subsequent encounter |
| Suicide-related event | Has the patient had a suicide-related event? | Suicide | Suicide | ICD10 | | T44.3X2S | Poisoning by other parasympatholytics [anticholinergics and antimuscarinics] and spasmolytics, intentional self-harm, sequela |
| Suicide-related event | Has the patient had a suicide-related event? | Suicide | Suicide | ICD10 | | T44.4X2A | Poisoning by predominantly alpha-adrenoreceptor agonists, intentional self-harm, initial encounter |
| Suicide-related event | Has the patient had a suicide-related event? | Suicide | Suicide | ICD10 | | T44.4X2D | Poisoning by predominantly alpha-adrenoreceptor agonists, intentional self-harm, subsequent encounter |
| Suicide-related event | Has the patient had a suicide-related event? | Suicide | Suicide | ICD10 | | T44.4X2S | Poisoning by predominantly alpha-adrenoreceptor agonists, intentional self-harm, sequela |
| Suicide-related event | Has the patient had a suicide-related event? | Suicide | Suicide | ICD10 | | T44.5X2A | Poisoning by predominantly beta-adrenoreceptor agonists, intentional self-harm, initial encounter |
| Suicide-related event | Has the patient had a suicide-related event? | Suicide | Suicide | ICD10 | | T44.5X2D | Poisoning by predominantly beta-adrenoreceptor agonists, intentional self-harm, subsequent encounter |
| Suicide-related event | Has the patient had a suicide-related event? | Suicide | Suicide | ICD10 | | T44.5X2S | Poisoning by predominantly beta-adrenoreceptor agonists, intentional self-harm, sequela |
| Suicide-related event | Has the patient had a suicide-related event? | Suicide | Suicide | ICD10 | | T44.6X2A | Poisoning by alpha-adrenoreceptor antagonists, intentional self-harm, initial encounter |
| Suicide-related event | Has the patient had a suicide-related event? | Suicide | Suicide | ICD10 | | T44.6X2D | Poisoning by alpha-adrenoreceptor antagonists, intentional self-harm, subsequent encounter |
| Suicide-related event | Has the patient had a suicide-related event? | Suicide | Suicide | ICD10 | | T44.6X2S | Poisoning by alpha-adrenoreceptor antagonists, intentional self-harm, sequela |
| Suicide-related event | Has the patient had a suicide-related event? | Suicide | Suicide | ICD10 | | T44.7X2A | Poisoning by beta-adrenoreceptor antagonists, intentional self-harm, initial encounter |
| Suicide-related event | Has the patient had a suicide-related event? | Suicide | Suicide | ICD10 | | T44.7X2D | Poisoning by beta-adrenoreceptor antagonists, intentional self-harm, subsequent encounter |
| Suicide-related event | Has the patient had a suicide-related event? | Suicide | Suicide | ICD10 | | T44.7X2S | Poisoning by beta-adrenoreceptor antagonists, intentional self-harm, sequela |
| Suicide-related event | Has the patient had a suicide-related event? | Suicide | Suicide | ICD10 | | T44.8X2A | Poisoning by centrally-acting and adrenergic-neuron-blocking agents, intentional self-harm, initial encounter |
| Suicide-related event | Has the patient had a suicide-related event? | Suicide | Suicide | ICD10 | | T44.8X2D | Poisoning by centrally-acting and adrenergic-neuron-blocking agents, intentional self-harm, subsequent encounter |
| Suicide-related event | Has the patient had a suicide-related event? | Suicide | Suicide | ICD10 | | T44.8X2S | Poisoning by centrally-acting and adrenergic-neuron-blocking agents, intentional self-harm, sequela |
| Suicide-related event | Has the patient had a suicide-related event? | Suicide | Suicide | ICD10 | | T44.902A | Poisoning by unspecified drugs primarily affecting the autonomic nervous system, intentional self-harm, initial encounter |
| Suicide-related event | Has the patient had a suicide-related event? | Suicide | Suicide | ICD10 | | T44.902D | Poisoning by unspecified drugs primarily affecting the autonomic nervous system, intentional self-harm, subsequent encounter |
| Suicide-related event | Has the patient had a suicide-related event? | Suicide | Suicide | ICD10 | | T44.902S | Poisoning by unspecified drugs primarily affecting the autonomic nervous system, intentional self-harm, sequela |
| Suicide-related event | Has the patient had a suicide-related event? | Suicide | Suicide | ICD10 | | T44.992A | Poisoning by other drug primarily affecting the autonomic nervous system, intentional self-harm, initial encounter |
| Suicide-related event | Has the patient had a suicide-related event? | Suicide | Suicide | ICD10 | | T44.992D | Poisoning by other drug primarily affecting the autonomic nervous system, intentional self-harm, subsequent encounter |
| Suicide-related event | Has the patient had a suicide-related event? | Suicide | Suicide | ICD10 | | T44.992S | Poisoning by other drug primarily affecting the autonomic nervous system, intentional self-harm, sequela |
| Suicide-related event | Has the patient had a suicide-related event? | Suicide | Suicide | ICD10 | | T45.0X2A | Poisoning by antiallergic and antiemetic drugs, intentional self-harm, initial encounter |
| Suicide-related event | Has the patient had a suicide-related event? | Suicide | Suicide | ICD10 | | T45.0X2D | Poisoning by antiallergic and antiemetic drugs, intentional self-harm, subsequent encounter |
| Suicide-related event | Has the patient had a suicide-related event? | Suicide | Suicide | ICD10 | | T45.0X2S | Poisoning by antiallergic and antiemetic drugs, intentional self-harm, sequela |
| Suicide-related event | Has the patient had a suicide-related event? | Suicide | Suicide | ICD10 | | T45.1X2A | Poisoning by antineoplastic and immunosuppressive drugs, intentional self-harm, initial encounter |
| Suicide-related event | Has the patient had a suicide-related event? | Suicide | Suicide | ICD10 | | T45.1X2D | Poisoning by antineoplastic and immunosuppressive drugs, intentional self-harm, subsequent encounter |
| Suicide-related event | Has the patient had a suicide-related event? | Suicide | Suicide | ICD10 | | T45.1X2S | Poisoning by antineoplastic and immunosuppressive drugs, intentional self-harm, sequela |
| Suicide-related event | Has the patient had a suicide-related event? | Suicide | Suicide | ICD10 | | T45.2X2A | Poisoning by vitamins, intentional self-harm, initial encounter |
| Suicide-related event | Has the patient had a suicide-related event? | Suicide | Suicide | ICD10 | | T45.2X2D | Poisoning by vitamins, intentional self-harm, subsequent encounter |
| Suicide-related event | Has the patient had a suicide-related event? | Suicide | Suicide | ICD10 | | T45.2X2S | Poisoning by vitamins, intentional self-harm, sequela |
| Suicide-related event | Has the patient had a suicide-related event? | Suicide | Suicide | ICD10 | | T45.3X2A | Poisoning by enzymes, intentional self-harm, initial encounter |
| Suicide-related event | Has the patient had a suicide-related event? | Suicide | Suicide | ICD10 | | T45.3X2D | Poisoning by enzymes, intentional self-harm, subsequent encounter |
| Suicide-related event | Has the patient had a suicide-related event? | Suicide | Suicide | ICD10 | | T45.3X2S | Poisoning by enzymes, intentional self-harm, sequela |
| Suicide-related event | Has the patient had a suicide-related event? | Suicide | Suicide | ICD10 | | T45.4X2A | Poisoning by iron and its compounds, intentional self-harm, initial encounter |
| Suicide-related event | Has the patient had a suicide-related event? | Suicide | Suicide | ICD10 | | T45.4X2D | Poisoning by iron and its compounds, intentional self-harm, subsequent encounter |
| Suicide-related event | Has the patient had a suicide-related event? | Suicide | Suicide | ICD10 | | T45.4X2S | Poisoning by iron and its compounds, intentional self-harm, sequela |
| Suicide-related event | Has the patient had a suicide-related event? | Suicide | Suicide | ICD10 | | T45.512A | Poisoning by anticoagulants, intentional self-harm, initial encounter |
| Suicide-related event | Has the patient had a suicide-related event? | Suicide | Suicide | ICD10 | | T45.512D | Poisoning by anticoagulants, intentional self-harm, subsequent encounter |
| Suicide-related event | Has the patient had a suicide-related event? | Suicide | Suicide | ICD10 | | T45.512S | Poisoning by anticoagulants, intentional self-harm, sequela |
| Suicide-related event | Has the patient had a suicide-related event? | Suicide | Suicide | ICD10 | | T45.522A | Poisoning by antithrombotic drugs, intentional self-harm, initial encounter |
| Suicide-related event | Has the patient had a suicide-related event? | Suicide | Suicide | ICD10 | | T45.522D | Poisoning by antithrombotic drugs, intentional self-harm, subsequent encounter |
| Suicide-related event | Has the patient had a suicide-related event? | Suicide | Suicide | ICD10 | | T45.522S | Poisoning by antithrombotic drugs, intentional self-harm, sequela |
| Suicide-related event | Has the patient had a suicide-related event? | Suicide | Suicide | ICD10 | | T45.602A | Poisoning by unspecified fibrinolysis-affecting drugs, intentional self-harm, initial encounter |
| Suicide-related event | Has the patient had a suicide-related event? | Suicide | Suicide | ICD10 | | T45.602D | Poisoning by unspecified fibrinolysis-affecting drugs, intentional self-harm, subsequent encounter |
| Suicide-related event | Has the patient had a suicide-related event? | Suicide | Suicide | ICD10 | | T45.602S | Poisoning by unspecified fibrinolysis-affecting drugs, intentional self-harm, sequela |
| Suicide-related event | Has the patient had a suicide-related event? | Suicide | Suicide | ICD10 | | T45.612A | Poisoning by thrombolytic drug, intentional self-harm, initial encounter |
| Suicide-related event | Has the patient had a suicide-related event? | Suicide | Suicide | ICD10 | | T45.612D | Poisoning by thrombolytic drug, intentional self-harm, subsequent encounter |
| Suicide-related event | Has the patient had a suicide-related event? | Suicide | Suicide | ICD10 | | T45.612S | Poisoning by thrombolytic drug, intentional self-harm, sequela |
| Suicide-related event | Has the patient had a suicide-related event? | Suicide | Suicide | ICD10 | | T45.622A | Poisoning by hemostatic drug, intentional self-harm, initial encounter |
| Suicide-related event | Has the patient had a suicide-related event? | Suicide | Suicide | ICD10 | | T45.622D | Poisoning by hemostatic drug, intentional self-harm, subsequent encounter |
| Suicide-related event | Has the patient had a suicide-related event? | Suicide | Suicide | ICD10 | | T45.622S | Poisoning by hemostatic drug, intentional self-harm, sequela |
| Suicide-related event | Has the patient had a suicide-related event? | Suicide | Suicide | ICD10 | | T45.692A | Poisoning by other fibrinolysis-affecting drugs, intentional self-harm, initial encounter |
| Suicide-related event | Has the patient had a suicide-related event? | Suicide | Suicide | ICD10 | | T45.692D | Poisoning by other fibrinolysis-affecting drugs, intentional self-harm, subsequent encounter |
| Suicide-related event | Has the patient had a suicide-related event? | Suicide | Suicide | ICD10 | | T45.692S | Poisoning by other fibrinolysis-affecting drugs, intentional self-harm, sequela |
| Suicide-related event | Has the patient had a suicide-related event? | Suicide | Suicide | ICD10 | | T45.7X2A | Poisoning by anticoagulant antagonists, vitamin K and other coagulants, intentional self-harm, initial encounter |
| Suicide-related event | Has the patient had a suicide-related event? | Suicide | Suicide | ICD10 | | T45.7X2D | Poisoning by anticoagulant antagonists, vitamin K and other coagulants, intentional self-harm, subsequent encounter |
| Suicide-related event | Has the patient had a suicide-related event? | Suicide | Suicide | ICD10 | | T45.7X2S | Poisoning by anticoagulant antagonists, vitamin K and other coagulants, intentional self-harm, sequela |
| Suicide-related event | Has the patient had a suicide-related event? | Suicide | Suicide | ICD10 | | T45.8X2A | Poisoning by other primarily systemic and hematological agents, intentional self-harm, initial encounter |
| Suicide-related event | Has the patient had a suicide-related event? | Suicide | Suicide | ICD10 | | T45.8X2D | Poisoning by other primarily systemic and hematological agents, intentional self-harm, subsequent encounter |
| Suicide-related event | Has the patient had a suicide-related event? | Suicide | Suicide | ICD10 | | T45.8X2S | Poisoning by other primarily systemic and hematological agents, intentional self-harm, sequela |
| Suicide-related event | Has the patient had a suicide-related event? | Suicide | Suicide | ICD10 | | T45.92XA | Poisoning by unspecified primarily systemic and hematological agent, intentional self-harm, initial encounter |
| Suicide-related event | Has the patient had a suicide-related event? | Suicide | Suicide | ICD10 | | T45.92XD | Poisoning by unspecified primarily systemic and hematological agent, intentional self-harm, subsequent encounter |
| Suicide-related event | Has the patient had a suicide-related event? | Suicide | Suicide | ICD10 | | T45.92XS | Poisoning by unspecified primarily systemic and hematological agent, intentional self-harm, sequela |
| Suicide-related event | Has the patient had a suicide-related event? | Suicide | Suicide | ICD10 | | T46.0X2A | Poisoning by cardiac-stimulant glycosides and drugs of similar action, intentional self-harm, initial encounter |
| Suicide-related event | Has the patient had a suicide-related event? | Suicide | Suicide | ICD10 | | T46.0X2D | Poisoning by cardiac-stimulant glycosides and drugs of similar action, intentional self-harm, subsequent encounter |
| Suicide-related event | Has the patient had a suicide-related event? | Suicide | Suicide | ICD10 | | T46.0X2S | Poisoning by cardiac-stimulant glycosides and drugs of similar action, intentional self-harm, sequela |
| Suicide-related event | Has the patient had a suicide-related event? | Suicide | Suicide | ICD10 | | T46.1X2A | Poisoning by calcium-channel blockers, intentional self-harm, initial encounter |
| Suicide-related event | Has the patient had a suicide-related event? | Suicide | Suicide | ICD10 | | T46.1X2D | Poisoning by calcium-channel blockers, intentional self-harm, subsequent encounter |
| Suicide-related event | Has the patient had a suicide-related event? | Suicide | Suicide | ICD10 | | T46.1X2S | Poisoning by calcium-channel blockers, intentional self-harm, sequela |
| Suicide-related event | Has the patient had a suicide-related event? | Suicide | Suicide | ICD10 | | T46.2X2A | Poisoning by other antidysrhythmic drugs, intentional self-harm, initial encounter |
| Suicide-related event | Has the patient had a suicide-related event? | Suicide | Suicide | ICD10 | | T46.2X2D | Poisoning by other antidysrhythmic drugs, intentional self-harm, subsequent encounter |
| Suicide-related event | Has the patient had a suicide-related event? | Suicide | Suicide | ICD10 | | T46.2X2S | Poisoning by other antidysrhythmic drugs, intentional self-harm, sequela |
| Suicide-related event | Has the patient had a suicide-related event? | Suicide | Suicide | ICD10 | | T46.3X2A | Poisoning by coronary vasodilators, intentional self-harm, initial encounter |
| Suicide-related event | Has the patient had a suicide-related event? | Suicide | Suicide | ICD10 | | T46.3X2D | Poisoning by coronary vasodilators, intentional self-harm, subsequent encounter |
| Suicide-related event | Has the patient had a suicide-related event? | Suicide | Suicide | ICD10 | | T46.3X2S | Poisoning by coronary vasodilators, intentional self-harm, sequela |
| Suicide-related event | Has the patient had a suicide-related event? | Suicide | Suicide | ICD10 | | T46.4X2A | Poisoning by angiotensin-converting-enzyme inhibitors, intentional self-harm, initial encounter |
| Suicide-related event | Has the patient had a suicide-related event? | Suicide | Suicide | ICD10 | | T46.4X2D | Poisoning by angiotensin-converting-enzyme inhibitors, intentional self-harm, subsequent encounter |
| Suicide-related event | Has the patient had a suicide-related event? | Suicide | Suicide | ICD10 | | T46.4X2S | Poisoning by angiotensin-converting-enzyme inhibitors, intentional self-harm, sequela |
| Suicide-related event | Has the patient had a suicide-related event? | Suicide | Suicide | ICD10 | | T46.5X2A | Poisoning by other antihypertensive drugs, intentional self-harm, initial encounter |
| Suicide-related event | Has the patient had a suicide-related event? | Suicide | Suicide | ICD10 | | T46.5X2D | Poisoning by other antihypertensive drugs, intentional self-harm, subsequent encounter |
| Suicide-related event | Has the patient had a suicide-related event? | Suicide | Suicide | ICD10 | | T46.5X2S | Poisoning by other antihypertensive drugs, intentional self-harm, sequela |
| Suicide-related event | Has the patient had a suicide-related event? | Suicide | Suicide | ICD10 | | T46.6X2A | Poisoning by antihyperlipidemic and antiarteriosclerotic drugs, intentional self-harm, initial encounter |
| Suicide-related event | Has the patient had a suicide-related event? | Suicide | Suicide | ICD10 | | T46.6X2D | Poisoning by antihyperlipidemic and antiarteriosclerotic drugs, intentional self-harm, subsequent encounter |
| Suicide-related event | Has the patient had a suicide-related event? | Suicide | Suicide | ICD10 | | T46.6X2S | Poisoning by antihyperlipidemic and antiarteriosclerotic drugs, intentional self-harm, sequela |
| Suicide-related event | Has the patient had a suicide-related event? | Suicide | Suicide | ICD10 | | T46.7X2A | Poisoning by peripheral vasodilators, intentional self-harm, initial encounter |
| Suicide-related event | Has the patient had a suicide-related event? | Suicide | Suicide | ICD10 | | T46.7X2D | Poisoning by peripheral vasodilators, intentional self-harm, subsequent encounter |
| Suicide-related event | Has the patient had a suicide-related event? | Suicide | Suicide | ICD10 | | T46.7X2S | Poisoning by peripheral vasodilators, intentional self-harm, sequela |
| Suicide-related event | Has the patient had a suicide-related event? | Suicide | Suicide | ICD10 | | T46.8X2A | Poisoning by antivaricose drugs, including sclerosing agents, intentional self-harm, initial encounter |
| Suicide-related event | Has the patient had a suicide-related event? | Suicide | Suicide | ICD10 | | T46.8X2D | Poisoning by antivaricose drugs, including sclerosing agents, intentional self-harm, subsequent encounter |
| Suicide-related event | Has the patient had a suicide-related event? | Suicide | Suicide | ICD10 | | T46.8X2S | Poisoning by antivaricose drugs, including sclerosing agents, intentional self-harm, sequela |
| Suicide-related event | Has the patient had a suicide-related event? | Suicide | Suicide | ICD10 | | T46.902A | Poisoning by unspecified agents primarily affecting the cardiovascular system, intentional self-harm, initial encounter |
| Suicide-related event | Has the patient had a suicide-related event? | Suicide | Suicide | ICD10 | | T46.902D | Poisoning by unspecified agents primarily affecting the cardiovascular system, intentional self-harm, subsequent encounter |
| Suicide-related event | Has the patient had a suicide-related event? | Suicide | Suicide | ICD10 | | T46.902S | Poisoning by unspecified agents primarily affecting the cardiovascular system, intentional self-harm, sequela |
| Suicide-related event | Has the patient had a suicide-related event? | Suicide | Suicide | ICD10 | | T46.992A | Poisoning by other agents primarily affecting the cardiovascular system, intentional self-harm, initial encounter |
| Suicide-related event | Has the patient had a suicide-related event? | Suicide | Suicide | ICD10 | | T46.992D | Poisoning by other agents primarily affecting the cardiovascular system, intentional self-harm, subsequent encounter |
| Suicide-related event | Has the patient had a suicide-related event? | Suicide | Suicide | ICD10 | | T46.992S | Poisoning by other agents primarily affecting the cardiovascular system, intentional self-harm, sequela |
| Suicide-related event | Has the patient had a suicide-related event? | Suicide | Suicide | ICD10 | | T47.0X2A | Poisoning by histamine H2-receptor blockers, intentional self-harm, initial encounter |
| Suicide-related event | Has the patient had a suicide-related event? | Suicide | Suicide | ICD10 | | T47.0X2D | Poisoning by histamine H2-receptor blockers, intentional self-harm, subsequent encounter |
| Suicide-related event | Has the patient had a suicide-related event? | Suicide | Suicide | ICD10 | | T47.0X2S | Poisoning by histamine H2-receptor blockers, intentional self-harm, sequela |
| Suicide-related event | Has the patient had a suicide-related event? | Suicide | Suicide | ICD10 | | T47.1X2A | Poisoning by other antacids and anti-gastric-secretion drugs, intentional self-harm, initial encounter |
| Suicide-related event | Has the patient had a suicide-related event? | Suicide | Suicide | ICD10 | | T47.1X2D | Poisoning by other antacids and anti-gastric-secretion drugs, intentional self-harm, subsequent encounter |
| Suicide-related event | Has the patient had a suicide-related event? | Suicide | Suicide | ICD10 | | T47.1X2S | Poisoning by other antacids and anti-gastric-secretion drugs, intentional self-harm, sequela |
| Suicide-related event | Has the patient had a suicide-related event? | Suicide | Suicide | ICD10 | | T47.2X2A | Poisoning by stimulant laxatives, intentional self-harm, initial encounter |
| Suicide-related event | Has the patient had a suicide-related event? | Suicide | Suicide | ICD10 | | T47.2X2D | Poisoning by stimulant laxatives, intentional self-harm, subsequent encounter |
| Suicide-related event | Has the patient had a suicide-related event? | Suicide | Suicide | ICD10 | | T47.2X2S | Poisoning by stimulant laxatives, intentional self-harm, sequela |
| Suicide-related event | Has the patient had a suicide-related event? | Suicide | Suicide | ICD10 | | T47.3X2A | Poisoning by saline and osmotic laxatives, intentional self-harm, initial encounter |
| Suicide-related event | Has the patient had a suicide-related event? | Suicide | Suicide | ICD10 | | T47.3X2D | Poisoning by saline and osmotic laxatives, intentional self-harm, subsequent encounter |
| Suicide-related event | Has the patient had a suicide-related event? | Suicide | Suicide | ICD10 | | T47.3X2S | Poisoning by saline and osmotic laxatives, intentional self-harm, sequela |
| Suicide-related event | Has the patient had a suicide-related event? | Suicide | Suicide | ICD10 | | T47.4X2A | Poisoning by other laxatives, intentional self-harm, initial encounter |
| Suicide-related event | Has the patient had a suicide-related event? | Suicide | Suicide | ICD10 | | T47.4X2D | Poisoning by other laxatives, intentional self-harm, subsequent encounter |
| Suicide-related event | Has the patient had a suicide-related event? | Suicide | Suicide | ICD10 | | T47.4X2S | Poisoning by other laxatives, intentional self-harm, sequela |
| Suicide-related event | Has the patient had a suicide-related event? | Suicide | Suicide | ICD10 | | T47.5X2A | Poisoning by digestants, intentional self-harm, initial encounter |
| Suicide-related event | Has the patient had a suicide-related event? | Suicide | Suicide | ICD10 | | T47.5X2D | Poisoning by digestants, intentional self-harm, subsequent encounter |
| Suicide-related event | Has the patient had a suicide-related event? | Suicide | Suicide | ICD10 | | T47.5X2S | Poisoning by digestants, intentional self-harm, sequela |
| Suicide-related event | Has the patient had a suicide-related event? | Suicide | Suicide | ICD10 | | T47.6X2A | Poisoning by antidiarrheal drugs, intentional self-harm, initial encounter |
| Suicide-related event | Has the patient had a suicide-related event? | Suicide | Suicide | ICD10 | | T47.6X2D | Poisoning by antidiarrheal drugs, intentional self-harm, subsequent encounter |
| Suicide-related event | Has the patient had a suicide-related event? | Suicide | Suicide | ICD10 | | T47.6X2S | Poisoning by antidiarrheal drugs, intentional self-harm, sequela |
| Suicide-related event | Has the patient had a suicide-related event? | Suicide | Suicide | ICD10 | | T47.7X2A | Poisoning by emetics, intentional self-harm, initial encounter |
| Suicide-related event | Has the patient had a suicide-related event? | Suicide | Suicide | ICD10 | | T47.7X2D | Poisoning by emetics, intentional self-harm, subsequent encounter |
| Suicide-related event | Has the patient had a suicide-related event? | Suicide | Suicide | ICD10 | | T47.7X2S | Poisoning by emetics, intentional self-harm, sequela |
| Suicide-related event | Has the patient had a suicide-related event? | Suicide | Suicide | ICD10 | | T47.8X2A | Poisoning by other agents primarily affecting gastrointestinal system, intentional self-harm, initial encounter |
| Suicide-related event | Has the patient had a suicide-related event? | Suicide | Suicide | ICD10 | | T47.8X2D | Poisoning by other agents primarily affecting gastrointestinal system, intentional self-harm, subsequent encounter |
| Suicide-related event | Has the patient had a suicide-related event? | Suicide | Suicide | ICD10 | | T47.8X2S | Poisoning by other agents primarily affecting gastrointestinal system, intentional self-harm, sequela |
| Suicide-related event | Has the patient had a suicide-related event? | Suicide | Suicide | ICD10 | | T47.92XA | Poisoning by unspecified agents primarily affecting the gastrointestinal system, intentional self-harm, initial encounter |
| Suicide-related event | Has the patient had a suicide-related event? | Suicide | Suicide | ICD10 | | T47.92XD | Poisoning by unspecified agents primarily affecting the gastrointestinal system, intentional self-harm, subsequent encounter |
| Suicide-related event | Has the patient had a suicide-related event? | Suicide | Suicide | ICD10 | | T47.92XS | Poisoning by unspecified agents primarily affecting the gastrointestinal system, intentional self-harm, sequela |
| Suicide-related event | Has the patient had a suicide-related event? | Suicide | Suicide | ICD10 | | T48.0X2A | Poisoning by oxytocic drugs, intentional self-harm, initial encounter |
| Suicide-related event | Has the patient had a suicide-related event? | Suicide | Suicide | ICD10 | | T48.0X2D | Poisoning by oxytocic drugs, intentional self-harm, subsequent encounter |
| Suicide-related event | Has the patient had a suicide-related event? | Suicide | Suicide | ICD10 | | T48.0X2S | Poisoning by oxytocic drugs, intentional self-harm, sequela |
| Suicide-related event | Has the patient had a suicide-related event? | Suicide | Suicide | ICD10 | | T48.1X2A | Poisoning by skeletal muscle relaxants [neuromuscular blocking agents], intentional self-harm, initial encounter |
| Suicide-related event | Has the patient had a suicide-related event? | Suicide | Suicide | ICD10 | | T48.1X2D | Poisoning by skeletal muscle relaxants [neuromuscular blocking agents], intentional self-harm, subsequent encounter |
| Suicide-related event | Has the patient had a suicide-related event? | Suicide | Suicide | ICD10 | | T48.1X2S | Poisoning by skeletal muscle relaxants [neuromuscular blocking agents], intentional self-harm, sequela |
| Suicide-related event | Has the patient had a suicide-related event? | Suicide | Suicide | ICD10 | | T48.202A | Poisoning by unspecified drugs acting on muscles, intentional self-harm, initial encounter |
| Suicide-related event | Has the patient had a suicide-related event? | Suicide | Suicide | ICD10 | | T48.202D | Poisoning by unspecified drugs acting on muscles, intentional self-harm, subsequent encounter |
| Suicide-related event | Has the patient had a suicide-related event? | Suicide | Suicide | ICD10 | | T48.202S | Poisoning by unspecified drugs acting on muscles, intentional self-harm, sequela |
| Suicide-related event | Has the patient had a suicide-related event? | Suicide | Suicide | ICD10 | | T48.292A | Poisoning by other drugs acting on muscles, intentional self-harm, initial encounter |
| Suicide-related event | Has the patient had a suicide-related event? | Suicide | Suicide | ICD10 | | T48.292D | Poisoning by other drugs acting on muscles, intentional self-harm, subsequent encounter |
| Suicide-related event | Has the patient had a suicide-related event? | Suicide | Suicide | ICD10 | | T48.292S | Poisoning by other drugs acting on muscles, intentional self-harm, sequela |
| Suicide-related event | Has the patient had a suicide-related event? | Suicide | Suicide | ICD10 | | T48.3X2A | Poisoning by antitussives, intentional self-harm, initial encounter |
| Suicide-related event | Has the patient had a suicide-related event? | Suicide | Suicide | ICD10 | | T48.3X2D | Poisoning by antitussives, intentional self-harm, subsequent encounter |
| Suicide-related event | Has the patient had a suicide-related event? | Suicide | Suicide | ICD10 | | T48.3X2S | Poisoning by antitussives, intentional self-harm, sequela |
| Suicide-related event | Has the patient had a suicide-related event? | Suicide | Suicide | ICD10 | | T48.4X2A | Poisoning by expectorants, intentional self-harm, initial encounter |
| Suicide-related event | Has the patient had a suicide-related event? | Suicide | Suicide | ICD10 | | T48.4X2D | Poisoning by expectorants, intentional self-harm, subsequent encounter |
| Suicide-related event | Has the patient had a suicide-related event? | Suicide | Suicide | ICD10 | | T48.4X2S | Poisoning by expectorants, intentional self-harm, sequela |
| Suicide-related event | Has the patient had a suicide-related event? | Suicide | Suicide | ICD10 | | T48.5X2A | Poisoning by other anti-common-cold drugs, intentional self-harm, initial encounter |
| Suicide-related event | Has the patient had a suicide-related event? | Suicide | Suicide | ICD10 | | T48.5X2D | Poisoning by other anti-common-cold drugs, intentional self-harm, subsequent encounter |
| Suicide-related event | Has the patient had a suicide-related event? | Suicide | Suicide | ICD10 | | T48.5X2S | Poisoning by other anti-common-cold drugs, intentional self-harm, sequela |
| Suicide-related event | Has the patient had a suicide-related event? | Suicide | Suicide | ICD10 | | T48.6X2A | Poisoning by antiasthmatics, intentional self-harm, initial encounter |
| Suicide-related event | Has the patient had a suicide-related event? | Suicide | Suicide | ICD10 | | T48.6X2D | Poisoning by antiasthmatics, intentional self-harm, subsequent encounter |
| Suicide-related event | Has the patient had a suicide-related event? | Suicide | Suicide | ICD10 | | T48.6X2S | Poisoning by antiasthmatics, intentional self-harm, sequela |
| Suicide-related event | Has the patient had a suicide-related event? | Suicide | Suicide | ICD10 | | T48.902A | Poisoning by unspecified agents primarily acting on the respiratory system, intentional self-harm, initial encounter |
| Suicide-related event | Has the patient had a suicide-related event? | Suicide | Suicide | ICD10 | | T48.902D | Poisoning by unspecified agents primarily acting on the respiratory system, intentional self-harm, subsequent encounter |
| Suicide-related event | Has the patient had a suicide-related event? | Suicide | Suicide | ICD10 | | T48.902S | Poisoning by unspecified agents primarily acting on the respiratory system, intentional self-harm, sequela |
| Suicide-related event | Has the patient had a suicide-related event? | Suicide | Suicide | ICD10 | | T48.992A | Poisoning by other agents primarily acting on the respiratory system, intentional self-harm, initial encounter |
| Suicide-related event | Has the patient had a suicide-related event? | Suicide | Suicide | ICD10 | | T48.992D | Poisoning by other agents primarily acting on the respiratory system, intentional self-harm, subsequent encounter |
| Suicide-related event | Has the patient had a suicide-related event? | Suicide | Suicide | ICD10 | | T48.992S | Poisoning by other agents primarily acting on the respiratory system, intentional self-harm, sequela |
| Suicide-related event | Has the patient had a suicide-related event? | Suicide | Suicide | ICD10 | | T49.0X2A | Poisoning by local antifungal, anti-infective and anti-inflammatory drugs, intentional self-harm, initial encounter |
| Suicide-related event | Has the patient had a suicide-related event? | Suicide | Suicide | ICD10 | | T49.0X2D | Poisoning by local antifungal, anti-infective and anti-inflammatory drugs, intentional self-harm, subsequent encounter |
| Suicide-related event | Has the patient had a suicide-related event? | Suicide | Suicide | ICD10 | | T49.0X2S | Poisoning by local antifungal, anti-infective and anti-inflammatory drugs, intentional self-harm, sequela |
| Suicide-related event | Has the patient had a suicide-related event? | Suicide | Suicide | ICD10 | | T49.1X2A | Poisoning by antipruritics, intentional self-harm, initial encounter |
| Suicide-related event | Has the patient had a suicide-related event? | Suicide | Suicide | ICD10 | | T49.1X2D | Poisoning by antipruritics, intentional self-harm, subsequent encounter |
| Suicide-related event | Has the patient had a suicide-related event? | Suicide | Suicide | ICD10 | | T49.1X2S | Poisoning by antipruritics, intentional self-harm, sequela |
| Suicide-related event | Has the patient had a suicide-related event? | Suicide | Suicide | ICD10 | | T49.2X2A | Poisoning by local astringents and local detergents, intentional self-harm, initial encounter |
| Suicide-related event | Has the patient had a suicide-related event? | Suicide | Suicide | ICD10 | | T49.2X2D | Poisoning by local astringents and local detergents, intentional self-harm, subsequent encounter |
| Suicide-related event | Has the patient had a suicide-related event? | Suicide | Suicide | ICD10 | | T49.2X2S | Poisoning by local astringents and local detergents, intentional self-harm, sequela |
| Suicide-related event | Has the patient had a suicide-related event? | Suicide | Suicide | ICD10 | | T49.3X2A | Poisoning by emollients, demulcents and protectants, intentional self-harm, initial encounter |
| Suicide-related event | Has the patient had a suicide-related event? | Suicide | Suicide | ICD10 | | T49.3X2D | Poisoning by emollients, demulcents and protectants, intentional self-harm, subsequent encounter |
| Suicide-related event | Has the patient had a suicide-related event? | Suicide | Suicide | ICD10 | | T49.3X2S | Poisoning by emollients, demulcents and protectants, intentional self-harm, sequela |
| Suicide-related event | Has the patient had a suicide-related event? | Suicide | Suicide | ICD10 | | T49.4X2A | Poisoning by keratolytics, keratoplastics, and other hair treatment drugs and preparations, intentional self-harm, initial encounter |
| Suicide-related event | Has the patient had a suicide-related event? | Suicide | Suicide | ICD10 | | T49.4X2D | Poisoning by keratolytics, keratoplastics, and other hair treatment drugs and preparations, intentional self-harm, subsequent encounter |
| Suicide-related event | Has the patient had a suicide-related event? | Suicide | Suicide | ICD10 | | T49.4X2S | Poisoning by keratolytics, keratoplastics, and other hair treatment drugs and preparations, intentional self-harm, sequela |
| Suicide-related event | Has the patient had a suicide-related event? | Suicide | Suicide | ICD10 | | T49.5X2A | Poisoning by ophthalmological drugs and preparations, intentional self-harm, initial encounter |
| Suicide-related event | Has the patient had a suicide-related event? | Suicide | Suicide | ICD10 | | T49.5X2D | Poisoning by ophthalmological drugs and preparations, intentional self-harm, subsequent encounter |
| Suicide-related event | Has the patient had a suicide-related event? | Suicide | Suicide | ICD10 | | T49.5X2S | Poisoning by ophthalmological drugs and preparations, intentional self-harm, sequela |
| Suicide-related event | Has the patient had a suicide-related event? | Suicide | Suicide | ICD10 | | T49.6X2A | Poisoning by otorhinolaryngological drugs and preparations, intentional self-harm, initial encounter |
| Suicide-related event | Has the patient had a suicide-related event? | Suicide | Suicide | ICD10 | | T49.6X2D | Poisoning by otorhinolaryngological drugs and preparations, intentional self-harm, subsequent encounter |
| Suicide-related event | Has the patient had a suicide-related event? | Suicide | Suicide | ICD10 | | T49.6X2S | Poisoning by otorhinolaryngological drugs and preparations, intentional self-harm, sequela |
| Suicide-related event | Has the patient had a suicide-related event? | Suicide | Suicide | ICD10 | | T49.7X2A | Poisoning by dental drugs, topically applied, intentional self-harm, initial encounter |
| Suicide-related event | Has the patient had a suicide-related event? | Suicide | Suicide | ICD10 | | T49.7X2D | Poisoning by dental drugs, topically applied, intentional self-harm, subsequent encounter |
| Suicide-related event | Has the patient had a suicide-related event? | Suicide | Suicide | ICD10 | | T49.7X2S | Poisoning by dental drugs, topically applied, intentional self-harm, sequela |
| Suicide-related event | Has the patient had a suicide-related event? | Suicide | Suicide | ICD10 | | T49.8X2A | Poisoning by other topical agents, intentional self-harm, initial encounter |
| Suicide-related event | Has the patient had a suicide-related event? | Suicide | Suicide | ICD10 | | T49.8X2D | Poisoning by other topical agents, intentional self-harm, subsequent encounter |
| Suicide-related event | Has the patient had a suicide-related event? | Suicide | Suicide | ICD10 | | T49.8X2S | Poisoning by other topical agents, intentional self-harm, sequela |
| Suicide-related event | Has the patient had a suicide-related event? | Suicide | Suicide | ICD10 | | T49.92XA | Poisoning by unspecified topical agent, intentional self-harm, initial encounter |
| Suicide-related event | Has the patient had a suicide-related event? | Suicide | Suicide | ICD10 | | T49.92XD | Poisoning by unspecified topical agent, intentional self-harm, subsequent encounter |
| Suicide-related event | Has the patient had a suicide-related event? | Suicide | Suicide | ICD10 | | T49.92XS | Poisoning by unspecified topical agent, intentional self-harm, sequela |
| Suicide-related event | Has the patient had a suicide-related event? | Suicide | Suicide | ICD10 | | T50.0X2A | Poisoning by mineralocorticoids and their antagonists, intentional self-harm, initial encounter |
| Suicide-related event | Has the patient had a suicide-related event? | Suicide | Suicide | ICD10 | | T50.0X2D | Poisoning by mineralocorticoids and their antagonists, intentional self-harm, subsequent encounter |
| Suicide-related event | Has the patient had a suicide-related event? | Suicide | Suicide | ICD10 | | T50.0X2S | Poisoning by mineralocorticoids and their antagonists, intentional self-harm, sequela |
| Suicide-related event | Has the patient had a suicide-related event? | Suicide | Suicide | ICD10 | | T50.1X2A | Poisoning by loop [high-ceiling] diuretics, intentional self-harm, initial encounter |
| Suicide-related event | Has the patient had a suicide-related event? | Suicide | Suicide | ICD10 | | T50.1X2D | Poisoning by loop [high-ceiling] diuretics, intentional self-harm, subsequent encounter |
| Suicide-related event | Has the patient had a suicide-related event? | Suicide | Suicide | ICD10 | | T50.1X2S | Poisoning by loop [high-ceiling] diuretics, intentional self-harm, sequela |
| Suicide-related event | Has the patient had a suicide-related event? | Suicide | Suicide | ICD10 | | T50.2X2A | Poisoning by carbonic-anhydrase inhibitors, benzothiadiazides and other diuretics, intentional self-harm, initial encounter |
| Suicide-related event | Has the patient had a suicide-related event? | Suicide | Suicide | ICD10 | | T50.2X2D | Poisoning by carbonic-anhydrase inhibitors, benzothiadiazides and other diuretics, intentional self-harm, subsequent encounter |
| Suicide-related event | Has the patient had a suicide-related event? | Suicide | Suicide | ICD10 | | T50.2X2S | Poisoning by carbonic-anhydrase inhibitors, benzothiadiazides and other diuretics, intentional self-harm, sequela |
| Suicide-related event | Has the patient had a suicide-related event? | Suicide | Suicide | ICD10 | | T50.3X2A | Poisoning by electrolytic, caloric and water-balance agents, intentional self-harm, initial encounter |
| Suicide-related event | Has the patient had a suicide-related event? | Suicide | Suicide | ICD10 | | T50.3X2D | Poisoning by electrolytic, caloric and water-balance agents, intentional self-harm, subsequent encounter |
| Suicide-related event | Has the patient had a suicide-related event? | Suicide | Suicide | ICD10 | | T50.3X2S | Poisoning by electrolytic, caloric and water-balance agents, intentional self-harm, sequela |
| Suicide-related event | Has the patient had a suicide-related event? | Suicide | Suicide | ICD10 | | T50.4X2A | Poisoning by drugs affecting uric acid metabolism, intentional self-harm, initial encounter |
| Suicide-related event | Has the patient had a suicide-related event? | Suicide | Suicide | ICD10 | | T50.4X2D | Poisoning by drugs affecting uric acid metabolism, intentional self-harm, subsequent encounter |
| Suicide-related event | Has the patient had a suicide-related event? | Suicide | Suicide | ICD10 | | T50.4X2S | Poisoning by drugs affecting uric acid metabolism, intentional self-harm, sequela |
| Suicide-related event | Has the patient had a suicide-related event? | Suicide | Suicide | ICD10 | | T50.5X2A | Poisoning by appetite depressants, intentional self-harm, initial encounter |
| Suicide-related event | Has the patient had a suicide-related event? | Suicide | Suicide | ICD10 | | T50.5X2D | Poisoning by appetite depressants, intentional self-harm, subsequent encounter |
| Suicide-related event | Has the patient had a suicide-related event? | Suicide | Suicide | ICD10 | | T50.5X2S | Poisoning by appetite depressants, intentional self-harm, sequela |
| Suicide-related event | Has the patient had a suicide-related event? | Suicide | Suicide | ICD10 | | T50.6X2A | Poisoning by antidotes and chelating agents, intentional self-harm, initial encounter |
| Suicide-related event | Has the patient had a suicide-related event? | Suicide | Suicide | ICD10 | | T50.6X2D | Poisoning by antidotes and chelating agents, intentional self-harm, subsequent encounter |
| Suicide-related event | Has the patient had a suicide-related event? | Suicide | Suicide | ICD10 | | T50.6X2S | Poisoning by antidotes and chelating agents, intentional self-harm, sequela |
| Suicide-related event | Has the patient had a suicide-related event? | Suicide | Suicide | ICD10 | | T50.7X2A | Poisoning by analeptics and opioid receptor antagonists, intentional self-harm, initial encounter |
| Suicide-related event | Has the patient had a suicide-related event? | Suicide | Suicide | ICD10 | | T50.7X2D | Poisoning by analeptics and opioid receptor antagonists, intentional self-harm, subsequent encounter |
| Suicide-related event | Has the patient had a suicide-related event? | Suicide | Suicide | ICD10 | | T50.7X2S | Poisoning by analeptics and opioid receptor antagonists, intentional self-harm, sequela |
| Suicide-related event | Has the patient had a suicide-related event? | Suicide | Suicide | ICD10 | | T50.8X2A | Poisoning by diagnostic agents, intentional self-harm, initial encounter |
| Suicide-related event | Has the patient had a suicide-related event? | Suicide | Suicide | ICD10 | | T50.8X2D | Poisoning by diagnostic agents, intentional self-harm, subsequent encounter |
| Suicide-related event | Has the patient had a suicide-related event? | Suicide | Suicide | ICD10 | | T50.8X2S | Poisoning by diagnostic agents, intentional self-harm, sequela |
| Suicide-related event | Has the patient had a suicide-related event? | Suicide | Suicide | ICD10 | | T50.902A | Poisoning by unspecified drugs, medicaments and biological substances, intentional self-harm, initial encounter |
| Suicide-related event | Has the patient had a suicide-related event? | Suicide | Suicide | ICD10 | | T50.902D | Poisoning by unspecified drugs, medicaments and biological substances, intentional self-harm, subsequent encounter |
| Suicide-related event | Has the patient had a suicide-related event? | Suicide | Suicide | ICD10 | | T50.902S | Poisoning by unspecified drugs, medicaments and biological substances, intentional self-harm, sequela |
| Suicide-related event | Has the patient had a suicide-related event? | Suicide | Suicide | ICD10 | | T50.992A | Poisoning by other drugs, medicaments and biological substances, intentional self-harm, initial encounter |
| Suicide-related event | Has the patient had a suicide-related event? | Suicide | Suicide | ICD10 | | T50.992D | Poisoning by other drugs, medicaments and biological substances, intentional self-harm, subsequent encounter |
| Suicide-related event | Has the patient had a suicide-related event? | Suicide | Suicide | ICD10 | | T50.992S | Poisoning by other drugs, medicaments and biological substances, intentional self-harm, sequela |
| Suicide-related event | Has the patient had a suicide-related event? | Suicide | Suicide | ICD10 | | T50.A12A | Poisoning by pertussis vaccine, including combinations with a pertussis component, intentional self-harm, initial encounter |
| Suicide-related event | Has the patient had a suicide-related event? | Suicide | Suicide | ICD10 | | T50.A12D | Poisoning by pertussis vaccine, including combinations with a pertussis component, intentional self-harm, subsequent encounter |
| Suicide-related event | Has the patient had a suicide-related event? | Suicide | Suicide | ICD10 | | T50.A12S | Poisoning by pertussis vaccine, including combinations with a pertussis component, intentional self-harm, sequela |
| Suicide-related event | Has the patient had a suicide-related event? | Suicide | Suicide | ICD10 | | T50.A22A | Poisoning by mixed bacterial vaccines without a pertussis component, intentional self-harm, initial encounter |
| Suicide-related event | Has the patient had a suicide-related event? | Suicide | Suicide | ICD10 | | T50.A22D | Poisoning by mixed bacterial vaccines without a pertussis component, intentional self-harm, subsequent encounter |
| Suicide-related event | Has the patient had a suicide-related event? | Suicide | Suicide | ICD10 | | T50.A22S | Poisoning by mixed bacterial vaccines without a pertussis component, intentional self-harm, sequela |
| Suicide-related event | Has the patient had a suicide-related event? | Suicide | Suicide | ICD10 | | T50.A92A | Poisoning by other bacterial vaccines, intentional self-harm, initial encounter |
| Suicide-related event | Has the patient had a suicide-related event? | Suicide | Suicide | ICD10 | | T50.A92D | Poisoning by other bacterial vaccines, intentional self-harm, subsequent encounter |
| Suicide-related event | Has the patient had a suicide-related event? | Suicide | Suicide | ICD10 | | T50.A92S | Poisoning by other bacterial vaccines, intentional self-harm, sequela |
| Suicide-related event | Has the patient had a suicide-related event? | Suicide | Suicide | ICD10 | | T50.B12A | Poisoning by smallpox vaccines, intentional self-harm, initial encounter |
| Suicide-related event | Has the patient had a suicide-related event? | Suicide | Suicide | ICD10 | | T50.B12D | Poisoning by smallpox vaccines, intentional self-harm, subsequent encounter |
| Suicide-related event | Has the patient had a suicide-related event? | Suicide | Suicide | ICD10 | | T50.B12S | Poisoning by smallpox vaccines, intentional self-harm, sequela |
| Suicide-related event | Has the patient had a suicide-related event? | Suicide | Suicide | ICD10 | | T50.B92A | Poisoning by other viral vaccines, intentional self-harm, initial encounter |
| Suicide-related event | Has the patient had a suicide-related event? | Suicide | Suicide | ICD10 | | T50.B92D | Poisoning by other viral vaccines, intentional self-harm, subsequent encounter |
| Suicide-related event | Has the patient had a suicide-related event? | Suicide | Suicide | ICD10 | | T50.B92S | Poisoning by other viral vaccines, intentional self-harm, sequela |
| Suicide-related event | Has the patient had a suicide-related event? | Suicide | Suicide | ICD10 | | T50.Z12A | Poisoning by immunoglobulin, intentional self-harm, initial encounter |
| Suicide-related event | Has the patient had a suicide-related event? | Suicide | Suicide | ICD10 | | T50.Z12D | Poisoning by immunoglobulin, intentional self-harm, subsequent encounter |
| Suicide-related event | Has the patient had a suicide-related event? | Suicide | Suicide | ICD10 | | T50.Z12S | Poisoning by immunoglobulin, intentional self-harm, sequela |
| Suicide-related event | Has the patient had a suicide-related event? | Suicide | Suicide | ICD10 | | T50.Z92A | Poisoning by other vaccines and biological substances, intentional self-harm, initial encounter |
| Suicide-related event | Has the patient had a suicide-related event? | Suicide | Suicide | ICD10 | | T50.Z92D | Poisoning by other vaccines and biological substances, intentional self-harm, subsequent encounter |
| Suicide-related event | Has the patient had a suicide-related event? | Suicide | Suicide | ICD10 | | T50.Z92S | Poisoning by other vaccines and biological substances, intentional self-harm, sequela |
| Suicide-related event | Has the patient had a suicide-related event? | Suicide | Suicide | ICD10 | | T51.0X2A | Toxic effect of ethanol, intentional self-harm, initial encounter |
| Suicide-related event | Has the patient had a suicide-related event? | Suicide | Suicide | ICD10 | | T51.0X2D | Toxic effect of ethanol, intentional self-harm, subsequent encounter |
| Suicide-related event | Has the patient had a suicide-related event? | Suicide | Suicide | ICD10 | | T51.0X2S | Toxic effect of ethanol, intentional self-harm, sequela |
| Suicide-related event | Has the patient had a suicide-related event? | Suicide | Suicide | ICD10 | | T51.1X2A | Toxic effect of methanol, intentional self-harm, initial encounter |
| Suicide-related event | Has the patient had a suicide-related event? | Suicide | Suicide | ICD10 | | T51.1X2D | Toxic effect of methanol, intentional self-harm, subsequent encounter |
| Suicide-related event | Has the patient had a suicide-related event? | Suicide | Suicide | ICD10 | | T51.1X2S | Toxic effect of methanol, intentional self-harm, sequela |
| Suicide-related event | Has the patient had a suicide-related event? | Suicide | Suicide | ICD10 | | T51.2X2A | Toxic effect of 2-Propanol, intentional self-harm, initial encounter |
| Suicide-related event | Has the patient had a suicide-related event? | Suicide | Suicide | ICD10 | | T51.2X2D | Toxic effect of 2-Propanol, intentional self-harm, subsequent encounter |
| Suicide-related event | Has the patient had a suicide-related event? | Suicide | Suicide | ICD10 | | T51.2X2S | Toxic effect of 2-Propanol, intentional self-harm, sequela |
| Suicide-related event | Has the patient had a suicide-related event? | Suicide | Suicide | ICD10 | | T51.3X2A | Toxic effect of fusel oil, intentional self-harm, initial encounter |
| Suicide-related event | Has the patient had a suicide-related event? | Suicide | Suicide | ICD10 | | T51.3X2D | Toxic effect of fusel oil, intentional self-harm, subsequent encounter |
| Suicide-related event | Has the patient had a suicide-related event? | Suicide | Suicide | ICD10 | | T51.3X2S | Toxic effect of fusel oil, intentional self-harm, sequela |
| Suicide-related event | Has the patient had a suicide-related event? | Suicide | Suicide | ICD10 | | T51.8X2A | Toxic effect of other alcohols, intentional self-harm, initial encounter |
| Suicide-related event | Has the patient had a suicide-related event? | Suicide | Suicide | ICD10 | | T51.8X2D | Toxic effect of other alcohols, intentional self-harm, subsequent encounter |
| Suicide-related event | Has the patient had a suicide-related event? | Suicide | Suicide | ICD10 | | T51.8X2S | Toxic effect of other alcohols, intentional self-harm, sequela |
| Suicide-related event | Has the patient had a suicide-related event? | Suicide | Suicide | ICD10 | | T51.92XA | Toxic effect of unspecified alcohol, intentional self-harm, initial encounter |
| Suicide-related event | Has the patient had a suicide-related event? | Suicide | Suicide | ICD10 | | T51.92XD | Toxic effect of unspecified alcohol, intentional self-harm, subsequent encounter |
| Suicide-related event | Has the patient had a suicide-related event? | Suicide | Suicide | ICD10 | | T51.92XS | Toxic effect of unspecified alcohol, intentional self-harm, sequela |
| Suicide-related event | Has the patient had a suicide-related event? | Suicide | Suicide | ICD10 | | T52.0X2A | Toxic effect of petroleum products, intentional self-harm, initial encounter |
| Suicide-related event | Has the patient had a suicide-related event? | Suicide | Suicide | ICD10 | | T52.0X2D | Toxic effect of petroleum products, intentional self-harm, subsequent encounter |
| Suicide-related event | Has the patient had a suicide-related event? | Suicide | Suicide | ICD10 | | T52.0X2S | Toxic effect of petroleum products, intentional self-harm, sequela |
| Suicide-related event | Has the patient had a suicide-related event? | Suicide | Suicide | ICD10 | | T52.1X2A | Toxic effect of benzene, intentional self-harm, initial encounter |
| Suicide-related event | Has the patient had a suicide-related event? | Suicide | Suicide | ICD10 | | T52.1X2D | Toxic effect of benzene, intentional self-harm, subsequent encounter |
| Suicide-related event | Has the patient had a suicide-related event? | Suicide | Suicide | ICD10 | | T52.1X2S | Toxic effect of benzene, intentional self-harm, sequela |
| Suicide-related event | Has the patient had a suicide-related event? | Suicide | Suicide | ICD10 | | T52.2X2A | Toxic effect of homologues of benzene, intentional self-harm, initial encounter |
| Suicide-related event | Has the patient had a suicide-related event? | Suicide | Suicide | ICD10 | | T52.2X2D | Toxic effect of homologues of benzene, intentional self-harm, subsequent encounter |
| Suicide-related event | Has the patient had a suicide-related event? | Suicide | Suicide | ICD10 | | T52.2X2S | Toxic effect of homologues of benzene, intentional self-harm, sequela |
| Suicide-related event | Has the patient had a suicide-related event? | Suicide | Suicide | ICD10 | | T52.3X2A | Toxic effect of glycols, intentional self-harm, initial encounter |
| Suicide-related event | Has the patient had a suicide-related event? | Suicide | Suicide | ICD10 | | T52.3X2D | Toxic effect of glycols, intentional self-harm, subsequent encounter |
| Suicide-related event | Has the patient had a suicide-related event? | Suicide | Suicide | ICD10 | | T52.3X2S | Toxic effect of glycols, intentional self-harm, sequela |
| Suicide-related event | Has the patient had a suicide-related event? | Suicide | Suicide | ICD10 | | T52.4X2A | Toxic effect of ketones, intentional self-harm, initial encounter |
| Suicide-related event | Has the patient had a suicide-related event? | Suicide | Suicide | ICD10 | | T52.4X2D | Toxic effect of ketones, intentional self-harm, subsequent encounter |
| Suicide-related event | Has the patient had a suicide-related event? | Suicide | Suicide | ICD10 | | T52.4X2S | Toxic effect of ketones, intentional self-harm, sequela |
| Suicide-related event | Has the patient had a suicide-related event? | Suicide | Suicide | ICD10 | | T52.8X2A | Toxic effect of other organic solvents, intentional self-harm, initial encounter |
| Suicide-related event | Has the patient had a suicide-related event? | Suicide | Suicide | ICD10 | | T52.8X2D | Toxic effect of other organic solvents, intentional self-harm, subsequent encounter |
| Suicide-related event | Has the patient had a suicide-related event? | Suicide | Suicide | ICD10 | | T52.8X2S | Toxic effect of other organic solvents, intentional self-harm, sequela |
| Suicide-related event | Has the patient had a suicide-related event? | Suicide | Suicide | ICD10 | | T52.92XA | Toxic effect of unspecified organic solvent, intentional self-harm, initial encounter |
| Suicide-related event | Has the patient had a suicide-related event? | Suicide | Suicide | ICD10 | | T52.92XD | Toxic effect of unspecified organic solvent, intentional self-harm, subsequent encounter |
| Suicide-related event | Has the patient had a suicide-related event? | Suicide | Suicide | ICD10 | | T52.92XS | Toxic effect of unspecified organic solvent, intentional self-harm, sequela |
| Suicide-related event | Has the patient had a suicide-related event? | Suicide | Suicide | ICD10 | | T53.0X2A | Toxic effect of carbon tetrachloride, intentional self-harm, initial encounter |
| Suicide-related event | Has the patient had a suicide-related event? | Suicide | Suicide | ICD10 | | T53.0X2D | Toxic effect of carbon tetrachloride, intentional self-harm, subsequent encounter |
| Suicide-related event | Has the patient had a suicide-related event? | Suicide | Suicide | ICD10 | | T53.0X2S | Toxic effect of carbon tetrachloride, intentional self-harm, sequela |
| Suicide-related event | Has the patient had a suicide-related event? | Suicide | Suicide | ICD10 | | T53.1X2A | Toxic effect of chloroform, intentional self-harm, initial encounter |
| Suicide-related event | Has the patient had a suicide-related event? | Suicide | Suicide | ICD10 | | T53.1X2D | Toxic effect of chloroform, intentional self-harm, subsequent encounter |
| Suicide-related event | Has the patient had a suicide-related event? | Suicide | Suicide | ICD10 | | T53.1X2S | Toxic effect of chloroform, intentional self-harm, sequela |
| Suicide-related event | Has the patient had a suicide-related event? | Suicide | Suicide | ICD10 | | T53.2X2A | Toxic effect of trichloroethylene, intentional self-harm, initial encounter |
| Suicide-related event | Has the patient had a suicide-related event? | Suicide | Suicide | ICD10 | | T53.2X2D | Toxic effect of trichloroethylene, intentional self-harm, subsequent encounter |
| Suicide-related event | Has the patient had a suicide-related event? | Suicide | Suicide | ICD10 | | T53.2X2S | Toxic effect of trichloroethylene, intentional self-harm, sequela |
| Suicide-related event | Has the patient had a suicide-related event? | Suicide | Suicide | ICD10 | | T53.3X2A | Toxic effect of tetrachloroethylene, intentional self-harm, initial encounter |
| Suicide-related event | Has the patient had a suicide-related event? | Suicide | Suicide | ICD10 | | T53.3X2D | Toxic effect of tetrachloroethylene, intentional self-harm, subsequent encounter |
| Suicide-related event | Has the patient had a suicide-related event? | Suicide | Suicide | ICD10 | | T53.3X2S | Toxic effect of tetrachloroethylene, intentional self-harm, sequela |
| Suicide-related event | Has the patient had a suicide-related event? | Suicide | Suicide | ICD10 | | T53.4X2A | Toxic effect of dichloromethane, intentional self-harm, initial encounter |
| Suicide-related event | Has the patient had a suicide-related event? | Suicide | Suicide | ICD10 | | T53.4X2D | Toxic effect of dichloromethane, intentional self-harm, subsequent encounter |
| Suicide-related event | Has the patient had a suicide-related event? | Suicide | Suicide | ICD10 | | T53.4X2S | Toxic effect of dichloromethane, intentional self-harm, sequela |
| Suicide-related event | Has the patient had a suicide-related event? | Suicide | Suicide | ICD10 | | T53.5X2A | Toxic effect of chlorofluorocarbons, intentional self-harm, initial encounter |
| Suicide-related event | Has the patient had a suicide-related event? | Suicide | Suicide | ICD10 | | T53.5X2D | Toxic effect of chlorofluorocarbons, intentional self-harm, subsequent encounter |
| Suicide-related event | Has the patient had a suicide-related event? | Suicide | Suicide | ICD10 | | T53.5X2S | Toxic effect of chlorofluorocarbons, intentional self-harm, sequela |
| Suicide-related event | Has the patient had a suicide-related event? | Suicide | Suicide | ICD10 | | T53.6X2A | Toxic effect of other halogen derivatives of aliphatic hydrocarbons, intentional self-harm, initial encounter |
| Suicide-related event | Has the patient had a suicide-related event? | Suicide | Suicide | ICD10 | | T53.6X2D | Toxic effect of other halogen derivatives of aliphatic hydrocarbons, intentional self-harm, subsequent encounter |
| Suicide-related event | Has the patient had a suicide-related event? | Suicide | Suicide | ICD10 | | T53.6X2S | Toxic effect of other halogen derivatives of aliphatic hydrocarbons, intentional self-harm, sequela |
| Suicide-related event | Has the patient had a suicide-related event? | Suicide | Suicide | ICD10 | | T53.7X2A | Toxic effect of other halogen derivatives of aromatic hydrocarbons, intentional self-harm, initial encounter |
| Suicide-related event | Has the patient had a suicide-related event? | Suicide | Suicide | ICD10 | | T53.7X2D | Toxic effect of other halogen derivatives of aromatic hydrocarbons, intentional self-harm, subsequent encounter |
| Suicide-related event | Has the patient had a suicide-related event? | Suicide | Suicide | ICD10 | | T53.7X2S | Toxic effect of other halogen derivatives of aromatic hydrocarbons, intentional self-harm, sequela |
| Suicide-related event | Has the patient had a suicide-related event? | Suicide | Suicide | ICD10 | | T53.92XA | Toxic effect of unspecified halogen derivatives of aliphatic and aromatic hydrocarbons, intentional self-harm, initial encounter |
| Suicide-related event | Has the patient had a suicide-related event? | Suicide | Suicide | ICD10 | | T53.92XD | Toxic effect of unspecified halogen derivatives of aliphatic and aromatic hydrocarbons, intentional self-harm, subsequent encounter |
| Suicide-related event | Has the patient had a suicide-related event? | Suicide | Suicide | ICD10 | | T53.92XS | Toxic effect of unspecified halogen derivatives of aliphatic and aromatic hydrocarbons, intentional self-harm, sequela |
| Suicide-related event | Has the patient had a suicide-related event? | Suicide | Suicide | ICD10 | | T54.0X2A | Toxic effect of phenol and phenol homologues, intentional self-harm, initial encounter |
| Suicide-related event | Has the patient had a suicide-related event? | Suicide | Suicide | ICD10 | | T54.0X2D | Toxic effect of phenol and phenol homologues, intentional self-harm, subsequent encounter |
| Suicide-related event | Has the patient had a suicide-related event? | Suicide | Suicide | ICD10 | | T54.0X2S | Toxic effect of phenol and phenol homologues, intentional self-harm, sequela |
| Suicide-related event | Has the patient had a suicide-related event? | Suicide | Suicide | ICD10 | | T54.1X2A | Toxic effect of other corrosive organic compounds, intentional self-harm, initial encounter |
| Suicide-related event | Has the patient had a suicide-related event? | Suicide | Suicide | ICD10 | | T54.1X2D | Toxic effect of other corrosive organic compounds, intentional self-harm, subsequent encounter |
| Suicide-related event | Has the patient had a suicide-related event? | Suicide | Suicide | ICD10 | | T54.1X2S | Toxic effect of other corrosive organic compounds, intentional self-harm, sequela |
| Suicide-related event | Has the patient had a suicide-related event? | Suicide | Suicide | ICD10 | | T54.2X2A | Toxic effect of corrosive acids and acid-like substances, intentional self-harm, initial encounter |
| Suicide-related event | Has the patient had a suicide-related event? | Suicide | Suicide | ICD10 | | T54.2X2D | Toxic effect of corrosive acids and acid-like substances, intentional self-harm, subsequent encounter |
| Suicide-related event | Has the patient had a suicide-related event? | Suicide | Suicide | ICD10 | | T54.2X2S | Toxic effect of corrosive acids and acid-like substances, intentional self-harm, sequela |
| Suicide-related event | Has the patient had a suicide-related event? | Suicide | Suicide | ICD10 | | T54.3X2A | Toxic effect of corrosive alkalis and alkali-like substances, intentional self-harm, initial encounter |
| Suicide-related event | Has the patient had a suicide-related event? | Suicide | Suicide | ICD10 | | T54.3X2D | Toxic effect of corrosive alkalis and alkali-like substances, intentional self-harm, subsequent encounter |
| Suicide-related event | Has the patient had a suicide-related event? | Suicide | Suicide | ICD10 | | T54.3X2S | Toxic effect of corrosive alkalis and alkali-like substances, intentional self-harm, sequela |
| Suicide-related event | Has the patient had a suicide-related event? | Suicide | Suicide | ICD10 | | T54.92XA | Toxic effect of unspecified corrosive substance, intentional self-harm, initial encounter |
| Suicide-related event | Has the patient had a suicide-related event? | Suicide | Suicide | ICD10 | | T54.92XD | Toxic effect of unspecified corrosive substance, intentional self-harm, subsequent encounter |
| Suicide-related event | Has the patient had a suicide-related event? | Suicide | Suicide | ICD10 | | T54.92XS | Toxic effect of unspecified corrosive substance, intentional self-harm, sequela |
| Suicide-related event | Has the patient had a suicide-related event? | Suicide | Suicide | ICD10 | | T55.0X2A | Toxic effect of soaps, intentional self-harm, initial encounter |
| Suicide-related event | Has the patient had a suicide-related event? | Suicide | Suicide | ICD10 | | T55.0X2D | Toxic effect of soaps, intentional self-harm, subsequent encounter |
| Suicide-related event | Has the patient had a suicide-related event? | Suicide | Suicide | ICD10 | | T55.0X2S | Toxic effect of soaps, intentional self-harm, sequela |
| Suicide-related event | Has the patient had a suicide-related event? | Suicide | Suicide | ICD10 | | T55.1X2A | Toxic effect of detergents, intentional self-harm, initial encounter |
| Suicide-related event | Has the patient had a suicide-related event? | Suicide | Suicide | ICD10 | | T55.1X2D | Toxic effect of detergents, intentional self-harm, subsequent encounter |
| Suicide-related event | Has the patient had a suicide-related event? | Suicide | Suicide | ICD10 | | T55.1X2S | Toxic effect of detergents, intentional self-harm, sequela |
| Suicide-related event | Has the patient had a suicide-related event? | Suicide | Suicide | ICD10 | | T56.0X2A | Toxic effect of lead and its compounds, intentional self-harm, initial encounter |
| Suicide-related event | Has the patient had a suicide-related event? | Suicide | Suicide | ICD10 | | T56.0X2D | Toxic effect of lead and its compounds, intentional self-harm, subsequent encounter |
| Suicide-related event | Has the patient had a suicide-related event? | Suicide | Suicide | ICD10 | | T56.0X2S | Toxic effect of lead and its compounds, intentional self-harm, sequela |
| Suicide-related event | Has the patient had a suicide-related event? | Suicide | Suicide | ICD10 | | T56.1X2A | Toxic effect of mercury and its compounds, intentional self-harm, initial encounter |
| Suicide-related event | Has the patient had a suicide-related event? | Suicide | Suicide | ICD10 | | T56.1X2D | Toxic effect of mercury and its compounds, intentional self-harm, subsequent encounter |
| Suicide-related event | Has the patient had a suicide-related event? | Suicide | Suicide | ICD10 | | T56.1X2S | Toxic effect of mercury and its compounds, intentional self-harm, sequela |
| Suicide-related event | Has the patient had a suicide-related event? | Suicide | Suicide | ICD10 | | T56.2X2A | Toxic effect of chromium and its compounds, intentional self-harm, initial encounter |
| Suicide-related event | Has the patient had a suicide-related event? | Suicide | Suicide | ICD10 | | T56.2X2D | Toxic effect of chromium and its compounds, intentional self-harm, subsequent encounter |
| Suicide-related event | Has the patient had a suicide-related event? | Suicide | Suicide | ICD10 | | T56.2X2S | Toxic effect of chromium and its compounds, intentional self-harm, sequela |
| Suicide-related event | Has the patient had a suicide-related event? | Suicide | Suicide | ICD10 | | T56.3X2A | Toxic effect of cadmium and its compounds, intentional self-harm, initial encounter |
| Suicide-related event | Has the patient had a suicide-related event? | Suicide | Suicide | ICD10 | | T56.3X2D | Toxic effect of cadmium and its compounds, intentional self-harm, subsequent encounter |
| Suicide-related event | Has the patient had a suicide-related event? | Suicide | Suicide | ICD10 | | T56.3X2S | Toxic effect of cadmium and its compounds, intentional self-harm, sequela |
| Suicide-related event | Has the patient had a suicide-related event? | Suicide | Suicide | ICD10 | | T56.4X2A | Toxic effect of copper and its compounds, intentional self-harm, initial encounter |
| Suicide-related event | Has the patient had a suicide-related event? | Suicide | Suicide | ICD10 | | T56.4X2D | Toxic effect of copper and its compounds, intentional self-harm, subsequent encounter |
| Suicide-related event | Has the patient had a suicide-related event? | Suicide | Suicide | ICD10 | | T56.4X2S | Toxic effect of copper and its compounds, intentional self-harm, sequela |
| Suicide-related event | Has the patient had a suicide-related event? | Suicide | Suicide | ICD10 | | T56.5X2A | Toxic effect of zinc and its compounds, intentional self-harm, initial encounter |
| Suicide-related event | Has the patient had a suicide-related event? | Suicide | Suicide | ICD10 | | T56.5X2D | Toxic effect of zinc and its compounds, intentional self-harm, subsequent encounter |
| Suicide-related event | Has the patient had a suicide-related event? | Suicide | Suicide | ICD10 | | T56.5X2S | Toxic effect of zinc and its compounds, intentional self-harm, sequela |
| Suicide-related event | Has the patient had a suicide-related event? | Suicide | Suicide | ICD10 | | T56.6X2A | Toxic effect of tin and its compounds, intentional self-harm, initial encounter |
| Suicide-related event | Has the patient had a suicide-related event? | Suicide | Suicide | ICD10 | | T56.6X2D | Toxic effect of tin and its compounds, intentional self-harm, subsequent encounter |
| Suicide-related event | Has the patient had a suicide-related event? | Suicide | Suicide | ICD10 | | T56.6X2S | Toxic effect of tin and its compounds, intentional self-harm, sequela |
| Suicide-related event | Has the patient had a suicide-related event? | Suicide | Suicide | ICD10 | | T56.7X2A | Toxic effect of beryllium and its compounds, intentional self-harm, initial encounter |
| Suicide-related event | Has the patient had a suicide-related event? | Suicide | Suicide | ICD10 | | T56.7X2D | Toxic effect of beryllium and its compounds, intentional self-harm, subsequent encounter |
| Suicide-related event | Has the patient had a suicide-related event? | Suicide | Suicide | ICD10 | | T56.7X2S | Toxic effect of beryllium and its compounds, intentional self-harm, sequela |
| Suicide-related event | Has the patient had a suicide-related event? | Suicide | Suicide | ICD10 | | T56.812A | Toxic effect of thallium, intentional self-harm, initial encounter |
| Suicide-related event | Has the patient had a suicide-related event? | Suicide | Suicide | ICD10 | | T56.812D | Toxic effect of thallium, intentional self-harm, subsequent encounter |
| Suicide-related event | Has the patient had a suicide-related event? | Suicide | Suicide | ICD10 | | T56.812S | Toxic effect of thallium, intentional self-harm, sequela |
| Suicide-related event | Has the patient had a suicide-related event? | Suicide | Suicide | ICD10 | | T56.892A | Toxic effect of other metals, intentional self-harm, initial encounter |
| Suicide-related event | Has the patient had a suicide-related event? | Suicide | Suicide | ICD10 | | T56.892D | Toxic effect of other metals, intentional self-harm, subsequent encounter |
| Suicide-related event | Has the patient had a suicide-related event? | Suicide | Suicide | ICD10 | | T56.892S | Toxic effect of other metals, intentional self-harm, sequela |
| Suicide-related event | Has the patient had a suicide-related event? | Suicide | Suicide | ICD10 | | T56.92XA | Toxic effect of unspecified metal, intentional self-harm, initial encounter |
| Suicide-related event | Has the patient had a suicide-related event? | Suicide | Suicide | ICD10 | | T56.92XD | Toxic effect of unspecified metal, intentional self-harm, subsequent encounter |
| Suicide-related event | Has the patient had a suicide-related event? | Suicide | Suicide | ICD10 | | T56.92XS | Toxic effect of unspecified metal, intentional self-harm, sequela |
| Suicide-related event | Has the patient had a suicide-related event? | Suicide | Suicide | ICD10 | | T57.0X2A | Toxic effect of arsenic and its compounds, intentional self-harm, initial encounter |
| Suicide-related event | Has the patient had a suicide-related event? | Suicide | Suicide | ICD10 | | T57.0X2D | Toxic effect of arsenic and its compounds, intentional self-harm, subsequent encounter |
| Suicide-related event | Has the patient had a suicide-related event? | Suicide | Suicide | ICD10 | | T57.0X2S | Toxic effect of arsenic and its compounds, intentional self-harm, sequela |
| Suicide-related event | Has the patient had a suicide-related event? | Suicide | Suicide | ICD10 | | T57.1X2A | Toxic effect of phosphorus and its compounds, intentional self-harm, initial encounter |
| Suicide-related event | Has the patient had a suicide-related event? | Suicide | Suicide | ICD10 | | T57.1X2D | Toxic effect of phosphorus and its compounds, intentional self-harm, subsequent encounter |
| Suicide-related event | Has the patient had a suicide-related event? | Suicide | Suicide | ICD10 | | T57.1X2S | Toxic effect of phosphorus and its compounds, intentional self-harm, sequela |
| Suicide-related event | Has the patient had a suicide-related event? | Suicide | Suicide | ICD10 | | T57.2X2A | Toxic effect of manganese and its compounds, intentional self-harm, initial encounter |
| Suicide-related event | Has the patient had a suicide-related event? | Suicide | Suicide | ICD10 | | T57.2X2D | Toxic effect of manganese and its compounds, intentional self-harm, subsequent encounter |
| Suicide-related event | Has the patient had a suicide-related event? | Suicide | Suicide | ICD10 | | T57.2X2S | Toxic effect of manganese and its compounds, intentional self-harm, sequela |
| Suicide-related event | Has the patient had a suicide-related event? | Suicide | Suicide | ICD10 | | T57.3X2A | Toxic effect of hydrogen cyanide, intentional self-harm, initial encounter |
| Suicide-related event | Has the patient had a suicide-related event? | Suicide | Suicide | ICD10 | | T57.3X2D | Toxic effect of hydrogen cyanide, intentional self-harm, subsequent encounter |
| Suicide-related event | Has the patient had a suicide-related event? | Suicide | Suicide | ICD10 | | T57.3X2S | Toxic effect of hydrogen cyanide, intentional self-harm, sequela |
| Suicide-related event | Has the patient had a suicide-related event? | Suicide | Suicide | ICD10 | | T57.8X2A | Toxic effect of other specified inorganic substances, intentional self-harm, initial encounter |
| Suicide-related event | Has the patient had a suicide-related event? | Suicide | Suicide | ICD10 | | T57.8X2D | Toxic effect of other specified inorganic substances, intentional self-harm, subsequent encounter |
| Suicide-related event | Has the patient had a suicide-related event? | Suicide | Suicide | ICD10 | | T57.8X2S | Toxic effect of other specified inorganic substances, intentional self-harm, sequela |
| Suicide-related event | Has the patient had a suicide-related event? | Suicide | Suicide | ICD10 | | T57.92XA | Toxic effect of unspecified inorganic substance, intentional self-harm, initial encounter |
| Suicide-related event | Has the patient had a suicide-related event? | Suicide | Suicide | ICD10 | | T57.92XD | Toxic effect of unspecified inorganic substance, intentional self-harm, subsequent encounter |
| Suicide-related event | Has the patient had a suicide-related event? | Suicide | Suicide | ICD10 | | T57.92XS | Toxic effect of unspecified inorganic substance, intentional self-harm, sequela |
| Suicide-related event | Has the patient had a suicide-related event? | Suicide | Suicide | ICD10 | | T58.02XA | Toxic effect of carbon monoxide from motor vehicle exhaust, intentional self-harm, initial encounter |
| Suicide-related event | Has the patient had a suicide-related event? | Suicide | Suicide | ICD10 | | T58.02XD | Toxic effect of carbon monoxide from motor vehicle exhaust, intentional self-harm, subsequent encounter |
| Suicide-related event | Has the patient had a suicide-related event? | Suicide | Suicide | ICD10 | | T58.02XS | Toxic effect of carbon monoxide from motor vehicle exhaust, intentional self-harm, sequela |
| Suicide-related event | Has the patient had a suicide-related event? | Suicide | Suicide | ICD10 | | T58.12XA | Toxic effect of carbon monoxide from utility gas, intentional self-harm, initial encounter |
| Suicide-related event | Has the patient had a suicide-related event? | Suicide | Suicide | ICD10 | | T58.12XD | Toxic effect of carbon monoxide from utility gas, intentional self-harm, subsequent encounter |
| Suicide-related event | Has the patient had a suicide-related event? | Suicide | Suicide | ICD10 | | T58.12XS | Toxic effect of carbon monoxide from utility gas, intentional self-harm, sequela |
| Suicide-related event | Has the patient had a suicide-related event? | Suicide | Suicide | ICD10 | | T58.2X2A | Toxic effect of carbon monoxide from incomplete combustion of other domestic fuels, intentional self-harm, initial encounter |
| Suicide-related event | Has the patient had a suicide-related event? | Suicide | Suicide | ICD10 | | T58.2X2D | Toxic effect of carbon monoxide from incomplete combustion of other domestic fuels, intentional self-harm, subsequent encounter |
| Suicide-related event | Has the patient had a suicide-related event? | Suicide | Suicide | ICD10 | | T58.2X2S | Toxic effect of carbon monoxide from incomplete combustion of other domestic fuels, intentional self-harm, sequela |
| Suicide-related event | Has the patient had a suicide-related event? | Suicide | Suicide | ICD10 | | T58.8X2A | Toxic effect of carbon monoxide from other source, intentional self-harm, initial encounter |
| Suicide-related event | Has the patient had a suicide-related event? | Suicide | Suicide | ICD10 | | T58.8X2D | Toxic effect of carbon monoxide from other source, intentional self-harm, subsequent encounter |
| Suicide-related event | Has the patient had a suicide-related event? | Suicide | Suicide | ICD10 | | T58.8X2S | Toxic effect of carbon monoxide from other source, intentional self-harm, sequela |
| Suicide-related event | Has the patient had a suicide-related event? | Suicide | Suicide | ICD10 | | T58.92XA | Toxic effect of carbon monoxide from unspecified source, intentional self-harm, initial encounter |
| Suicide-related event | Has the patient had a suicide-related event? | Suicide | Suicide | ICD10 | | T58.92XD | Toxic effect of carbon monoxide from unspecified source, intentional self-harm, subsequent encounter |
| Suicide-related event | Has the patient had a suicide-related event? | Suicide | Suicide | ICD10 | | T58.92XS | Toxic effect of carbon monoxide from unspecified source, intentional self-harm, sequela |
| Suicide-related event | Has the patient had a suicide-related event? | Suicide | Suicide | ICD10 | | T59.0X2A | Toxic effect of nitrogen oxides, intentional self-harm, initial encounter |
| Suicide-related event | Has the patient had a suicide-related event? | Suicide | Suicide | ICD10 | | T59.0X2D | Toxic effect of nitrogen oxides, intentional self-harm, subsequent encounter |
| Suicide-related event | Has the patient had a suicide-related event? | Suicide | Suicide | ICD10 | | T59.0X2S | Toxic effect of nitrogen oxides, intentional self-harm, sequela |
| Suicide-related event | Has the patient had a suicide-related event? | Suicide | Suicide | ICD10 | | T59.1X2A | Toxic effect of sulfur dioxide, intentional self-harm, initial encounter |
| Suicide-related event | Has the patient had a suicide-related event? | Suicide | Suicide | ICD10 | | T59.1X2D | Toxic effect of sulfur dioxide, intentional self-harm, subsequent encounter |
| Suicide-related event | Has the patient had a suicide-related event? | Suicide | Suicide | ICD10 | | T59.1X2S | Toxic effect of sulfur dioxide, intentional self-harm, sequela |
| Suicide-related event | Has the patient had a suicide-related event? | Suicide | Suicide | ICD10 | | T59.2X2A | Toxic effect of formaldehyde, intentional self-harm, initial encounter |
| Suicide-related event | Has the patient had a suicide-related event? | Suicide | Suicide | ICD10 | | T59.2X2D | Toxic effect of formaldehyde, intentional self-harm, subsequent encounter |
| Suicide-related event | Has the patient had a suicide-related event? | Suicide | Suicide | ICD10 | | T59.2X2S | Toxic effect of formaldehyde, intentional self-harm, sequela |
| Suicide-related event | Has the patient had a suicide-related event? | Suicide | Suicide | ICD10 | | T59.3X2A | Toxic effect of lacrimogenic gas, intentional self-harm, initial encounter |
| Suicide-related event | Has the patient had a suicide-related event? | Suicide | Suicide | ICD10 | | T59.3X2D | Toxic effect of lacrimogenic gas, intentional self-harm, subsequent encounter |
| Suicide-related event | Has the patient had a suicide-related event? | Suicide | Suicide | ICD10 | | T59.3X2S | Toxic effect of lacrimogenic gas, intentional self-harm, sequela |
| Suicide-related event | Has the patient had a suicide-related event? | Suicide | Suicide | ICD10 | | T59.4X2A | Toxic effect of chlorine gas, intentional self-harm, initial encounter |
| Suicide-related event | Has the patient had a suicide-related event? | Suicide | Suicide | ICD10 | | T59.4X2D | Toxic effect of chlorine gas, intentional self-harm, subsequent encounter |
| Suicide-related event | Has the patient had a suicide-related event? | Suicide | Suicide | ICD10 | | T59.4X2S | Toxic effect of chlorine gas, intentional self-harm, sequela |
| Suicide-related event | Has the patient had a suicide-related event? | Suicide | Suicide | ICD10 | | T59.5X2A | Toxic effect of fluorine gas and hydrogen fluoride, intentional self-harm, initial encounter |
| Suicide-related event | Has the patient had a suicide-related event? | Suicide | Suicide | ICD10 | | T59.5X2D | Toxic effect of fluorine gas and hydrogen fluoride, intentional self-harm, subsequent encounter |
| Suicide-related event | Has the patient had a suicide-related event? | Suicide | Suicide | ICD10 | | T59.5X2S | Toxic effect of fluorine gas and hydrogen fluoride, intentional self-harm, sequela |
| Suicide-related event | Has the patient had a suicide-related event? | Suicide | Suicide | ICD10 | | T59.6X2A | Toxic effect of hydrogen sulfide, intentional self-harm, initial encounter |
| Suicide-related event | Has the patient had a suicide-related event? | Suicide | Suicide | ICD10 | | T59.6X2D | Toxic effect of hydrogen sulfide, intentional self-harm, subsequent encounter |
| Suicide-related event | Has the patient had a suicide-related event? | Suicide | Suicide | ICD10 | | T59.6X2S | Toxic effect of hydrogen sulfide, intentional self-harm, sequela |
| Suicide-related event | Has the patient had a suicide-related event? | Suicide | Suicide | ICD10 | | T59.7X2A | Toxic effect of carbon dioxide, intentional self-harm, initial encounter |
| Suicide-related event | Has the patient had a suicide-related event? | Suicide | Suicide | ICD10 | | T59.7X2D | Toxic effect of carbon dioxide, intentional self-harm, subsequent encounter |
| Suicide-related event | Has the patient had a suicide-related event? | Suicide | Suicide | ICD10 | | T59.7X2S | Toxic effect of carbon dioxide, intentional self-harm, sequela |
| Suicide-related event | Has the patient had a suicide-related event? | Suicide | Suicide | ICD10 | | T59.812A | Toxic effect of smoke, intentional self-harm, initial encounter |
| Suicide-related event | Has the patient had a suicide-related event? | Suicide | Suicide | ICD10 | | T59.812D | Toxic effect of smoke, intentional self-harm, subsequent encounter |
| Suicide-related event | Has the patient had a suicide-related event? | Suicide | Suicide | ICD10 | | T59.812S | Toxic effect of smoke, intentional self-harm, sequela |
| Suicide-related event | Has the patient had a suicide-related event? | Suicide | Suicide | ICD10 | | T59.892A | Toxic effect of other specified gases, fumes and vapors, intentional self-harm, initial encounter |
| Suicide-related event | Has the patient had a suicide-related event? | Suicide | Suicide | ICD10 | | T59.892D | Toxic effect of other specified gases, fumes and vapors, intentional self-harm, subsequent encounter |
| Suicide-related event | Has the patient had a suicide-related event? | Suicide | Suicide | ICD10 | | T59.892S | Toxic effect of other specified gases, fumes and vapors, intentional self-harm, sequela |
| Suicide-related event | Has the patient had a suicide-related event? | Suicide | Suicide | ICD10 | | T59.92XA | Toxic effect of unspecified gases, fumes and vapors, intentional self-harm, initial encounter |
| Suicide-related event | Has the patient had a suicide-related event? | Suicide | Suicide | ICD10 | | T59.92XD | Toxic effect of unspecified gases, fumes and vapors, intentional self-harm, subsequent encounter |
| Suicide-related event | Has the patient had a suicide-related event? | Suicide | Suicide | ICD10 | | T59.92XS | Toxic effect of unspecified gases, fumes and vapors, intentional self-harm, sequela |
| Suicide-related event | Has the patient had a suicide-related event? | Suicide | Suicide | ICD10 | | T60.0X2A | Toxic effect of organophosphate and carbamate insecticides, intentional self-harm, initial encounter |
| Suicide-related event | Has the patient had a suicide-related event? | Suicide | Suicide | ICD10 | | T60.0X2D | Toxic effect of organophosphate and carbamate insecticides, intentional self-harm, subsequent encounter |
| Suicide-related event | Has the patient had a suicide-related event? | Suicide | Suicide | ICD10 | | T60.0X2S | Toxic effect of organophosphate and carbamate insecticides, intentional self-harm, sequela |
| Suicide-related event | Has the patient had a suicide-related event? | Suicide | Suicide | ICD10 | | T60.1X2A | Toxic effect of halogenated insecticides, intentional self-harm, initial encounter |
| Suicide-related event | Has the patient had a suicide-related event? | Suicide | Suicide | ICD10 | | T60.1X2D | Toxic effect of halogenated insecticides, intentional self-harm, subsequent encounter |
| Suicide-related event | Has the patient had a suicide-related event? | Suicide | Suicide | ICD10 | | T60.1X2S | Toxic effect of halogenated insecticides, intentional self-harm, sequela |
| Suicide-related event | Has the patient had a suicide-related event? | Suicide | Suicide | ICD10 | | T60.2X2A | Toxic effect of other insecticides, intentional self-harm, initial encounter |
| Suicide-related event | Has the patient had a suicide-related event? | Suicide | Suicide | ICD10 | | T60.2X2D | Toxic effect of other insecticides, intentional self-harm, subsequent encounter |
| Suicide-related event | Has the patient had a suicide-related event? | Suicide | Suicide | ICD10 | | T60.2X2S | Toxic effect of other insecticides, intentional self-harm, sequela |
| Suicide-related event | Has the patient had a suicide-related event? | Suicide | Suicide | ICD10 | | T60.3X2A | Toxic effect of herbicides and fungicides, intentional self-harm, initial encounter |
| Suicide-related event | Has the patient had a suicide-related event? | Suicide | Suicide | ICD10 | | T60.3X2D | Toxic effect of herbicides and fungicides, intentional self-harm, subsequent encounter |
| Suicide-related event | Has the patient had a suicide-related event? | Suicide | Suicide | ICD10 | | T60.3X2S | Toxic effect of herbicides and fungicides, intentional self-harm, sequela |
| Suicide-related event | Has the patient had a suicide-related event? | Suicide | Suicide | ICD10 | | T60.4X2A | Toxic effect of rodenticides, intentional self-harm, initial encounter |
| Suicide-related event | Has the patient had a suicide-related event? | Suicide | Suicide | ICD10 | | T60.4X2D | Toxic effect of rodenticides, intentional self-harm, subsequent encounter |
| Suicide-related event | Has the patient had a suicide-related event? | Suicide | Suicide | ICD10 | | T60.4X2S | Toxic effect of rodenticides, intentional self-harm, sequela |
| Suicide-related event | Has the patient had a suicide-related event? | Suicide | Suicide | ICD10 | | T60.8X2A | Toxic effect of other pesticides, intentional self-harm, initial encounter |
| Suicide-related event | Has the patient had a suicide-related event? | Suicide | Suicide | ICD10 | | T60.8X2D | Toxic effect of other pesticides, intentional self-harm, subsequent encounter |
| Suicide-related event | Has the patient had a suicide-related event? | Suicide | Suicide | ICD10 | | T60.8X2S | Toxic effect of other pesticides, intentional self-harm, sequela |
| Suicide-related event | Has the patient had a suicide-related event? | Suicide | Suicide | ICD10 | | T60.92XA | Toxic effect of unspecified pesticide, intentional self-harm, initial encounter |
| Suicide-related event | Has the patient had a suicide-related event? | Suicide | Suicide | ICD10 | | T60.92XD | Toxic effect of unspecified pesticide, intentional self-harm, subsequent encounter |
| Suicide-related event | Has the patient had a suicide-related event? | Suicide | Suicide | ICD10 | | T60.92XS | Toxic effect of unspecified pesticide, intentional self-harm, sequela |
| Suicide-related event | Has the patient had a suicide-related event? | Suicide | Suicide | ICD10 | | T61.02XA | Ciguatera fish poisoning, intentional self-harm, initial encounter |
| Suicide-related event | Has the patient had a suicide-related event? | Suicide | Suicide | ICD10 | | T61.02XD | Ciguatera fish poisoning, intentional self-harm, subsequent encounter |
| Suicide-related event | Has the patient had a suicide-related event? | Suicide | Suicide | ICD10 | | T61.02XS | Ciguatera fish poisoning, intentional self-harm, sequela |
| Suicide-related event | Has the patient had a suicide-related event? | Suicide | Suicide | ICD10 | | T61.12XA | Scombroid fish poisoning, intentional self-harm, initial encounter |
| Suicide-related event | Has the patient had a suicide-related event? | Suicide | Suicide | ICD10 | | T61.12XD | Scombroid fish poisoning, intentional self-harm, subsequent encounter |
| Suicide-related event | Has the patient had a suicide-related event? | Suicide | Suicide | ICD10 | | T61.12XS | Scombroid fish poisoning, intentional self-harm, sequela |
| Suicide-related event | Has the patient had a suicide-related event? | Suicide | Suicide | ICD10 | | T61.772A | Other fish poisoning, intentional self-harm, initial encounter |
| Suicide-related event | Has the patient had a suicide-related event? | Suicide | Suicide | ICD10 | | T61.772D | Other fish poisoning, intentional self-harm, subsequent encounter |
| Suicide-related event | Has the patient had a suicide-related event? | Suicide | Suicide | ICD10 | | T61.772S | Other fish poisoning, intentional self-harm, sequela |
| Suicide-related event | Has the patient had a suicide-related event? | Suicide | Suicide | ICD10 | | T61.782A | Other shellfish poisoning, intentional self-harm, initial encounter |
| Suicide-related event | Has the patient had a suicide-related event? | Suicide | Suicide | ICD10 | | T61.782D | Other shellfish poisoning, intentional self-harm, subsequent encounter |
| Suicide-related event | Has the patient had a suicide-related event? | Suicide | Suicide | ICD10 | | T61.782S | Other shellfish poisoning, intentional self-harm, sequela |
| Suicide-related event | Has the patient had a suicide-related event? | Suicide | Suicide | ICD10 | | T61.8X2A | Toxic effect of other seafood, intentional self-harm, initial encounter |
| Suicide-related event | Has the patient had a suicide-related event? | Suicide | Suicide | ICD10 | | T61.8X2D | Toxic effect of other seafood, intentional self-harm, subsequent encounter |
| Suicide-related event | Has the patient had a suicide-related event? | Suicide | Suicide | ICD10 | | T61.8X2S | Toxic effect of other seafood, intentional self-harm, sequela |
| Suicide-related event | Has the patient had a suicide-related event? | Suicide | Suicide | ICD10 | | T61.92XA | Toxic effect of unspecified seafood, intentional self-harm, initial encounter |
| Suicide-related event | Has the patient had a suicide-related event? | Suicide | Suicide | ICD10 | | T61.92XD | Toxic effect of unspecified seafood, intentional self-harm, subsequent encounter |
| Suicide-related event | Has the patient had a suicide-related event? | Suicide | Suicide | ICD10 | | T61.92XS | Toxic effect of unspecified seafood, intentional self-harm, sequela |
| Suicide-related event | Has the patient had a suicide-related event? | Suicide | Suicide | ICD10 | | T62.0X2A | Toxic effect of ingested mushrooms, intentional self-harm, initial encounter |
| Suicide-related event | Has the patient had a suicide-related event? | Suicide | Suicide | ICD10 | | T62.0X2D | Toxic effect of ingested mushrooms, intentional self-harm, subsequent encounter |
| Suicide-related event | Has the patient had a suicide-related event? | Suicide | Suicide | ICD10 | | T62.0X2S | Toxic effect of ingested mushrooms, intentional self-harm, sequela |
| Suicide-related event | Has the patient had a suicide-related event? | Suicide | Suicide | ICD10 | | T62.1X2A | Toxic effect of ingested berries, intentional self-harm, initial encounter |
| Suicide-related event | Has the patient had a suicide-related event? | Suicide | Suicide | ICD10 | | T62.1X2D | Toxic effect of ingested berries, intentional self-harm, subsequent encounter |
| Suicide-related event | Has the patient had a suicide-related event? | Suicide | Suicide | ICD10 | | T62.1X2S | Toxic effect of ingested berries, intentional self-harm, sequela |
| Suicide-related event | Has the patient had a suicide-related event? | Suicide | Suicide | ICD10 | | T62.2X2A | Toxic effect of other ingested (parts of) plant(s), intentional self-harm, initial encounter |
| Suicide-related event | Has the patient had a suicide-related event? | Suicide | Suicide | ICD10 | | T62.2X2D | Toxic effect of other ingested (parts of) plant(s), intentional self-harm, subsequent encounter |
| Suicide-related event | Has the patient had a suicide-related event? | Suicide | Suicide | ICD10 | | T62.2X2S | Toxic effect of other ingested (parts of) plant(s), intentional self-harm, sequela |
| Suicide-related event | Has the patient had a suicide-related event? | Suicide | Suicide | ICD10 | | T62.8X2A | Toxic effect of other specified noxious substances eaten as food, intentional self-harm, initial encounter |
| Suicide-related event | Has the patient had a suicide-related event? | Suicide | Suicide | ICD10 | | T62.8X2D | Toxic effect of other specified noxious substances eaten as food, intentional self-harm, subsequent encounter |
| Suicide-related event | Has the patient had a suicide-related event? | Suicide | Suicide | ICD10 | | T62.8X2S | Toxic effect of other specified noxious substances eaten as food, intentional self-harm, sequela |
| Suicide-related event | Has the patient had a suicide-related event? | Suicide | Suicide | ICD10 | | T62.92XA | Toxic effect of unspecified noxious substance eaten as food, intentional self-harm, initial encounter |
| Suicide-related event | Has the patient had a suicide-related event? | Suicide | Suicide | ICD10 | | T62.92XD | Toxic effect of unspecified noxious substance eaten as food, intentional self-harm, subsequent encounter |
| Suicide-related event | Has the patient had a suicide-related event? | Suicide | Suicide | ICD10 | | T62.92XS | Toxic effect of unspecified noxious substance eaten as food, intentional self-harm, sequela |
| Suicide-related event | Has the patient had a suicide-related event? | Suicide | Suicide | ICD10 | | T63.002A | Toxic effect of unspecified snake venom, intentional self-harm, initial encounter |
| Suicide-related event | Has the patient had a suicide-related event? | Suicide | Suicide | ICD10 | | T63.002D | Toxic effect of unspecified snake venom, intentional self-harm, subsequent encounter |
| Suicide-related event | Has the patient had a suicide-related event? | Suicide | Suicide | ICD10 | | T63.002S | Toxic effect of unspecified snake venom, intentional self-harm, sequela |
| Suicide-related event | Has the patient had a suicide-related event? | Suicide | Suicide | ICD10 | | T63.012A | Toxic effect of rattlesnake venom, intentional self-harm, initial encounter |
| Suicide-related event | Has the patient had a suicide-related event? | Suicide | Suicide | ICD10 | | T63.012D | Toxic effect of rattlesnake venom, intentional self-harm, subsequent encounter |
| Suicide-related event | Has the patient had a suicide-related event? | Suicide | Suicide | ICD10 | | T63.012S | Toxic effect of rattlesnake venom, intentional self-harm, sequela |
| Suicide-related event | Has the patient had a suicide-related event? | Suicide | Suicide | ICD10 | | T63.022A | Toxic effect of coral snake venom, intentional self-harm, initial encounter |
| Suicide-related event | Has the patient had a suicide-related event? | Suicide | Suicide | ICD10 | | T63.022D | Toxic effect of coral snake venom, intentional self-harm, subsequent encounter |
| Suicide-related event | Has the patient had a suicide-related event? | Suicide | Suicide | ICD10 | | T63.022S | Toxic effect of coral snake venom, intentional self-harm, sequela |
| Suicide-related event | Has the patient had a suicide-related event? | Suicide | Suicide | ICD10 | | T63.032A | Toxic effect of taipan venom, intentional self-harm, initial encounter |
| Suicide-related event | Has the patient had a suicide-related event? | Suicide | Suicide | ICD10 | | T63.032D | Toxic effect of taipan venom, intentional self-harm, subsequent encounter |
| Suicide-related event | Has the patient had a suicide-related event? | Suicide | Suicide | ICD10 | | T63.032S | Toxic effect of taipan venom, intentional self-harm, sequela |
| Suicide-related event | Has the patient had a suicide-related event? | Suicide | Suicide | ICD10 | | T63.042A | Toxic effect of cobra venom, intentional self-harm, initial encounter |
| Suicide-related event | Has the patient had a suicide-related event? | Suicide | Suicide | ICD10 | | T63.042D | Toxic effect of cobra venom, intentional self-harm, subsequent encounter |
| Suicide-related event | Has the patient had a suicide-related event? | Suicide | Suicide | ICD10 | | T63.042S | Toxic effect of cobra venom, intentional self-harm, sequela |
| Suicide-related event | Has the patient had a suicide-related event? | Suicide | Suicide | ICD10 | | T63.062A | Toxic effect of venom of other North and South American snake, intentional self-harm, initial encounter |
| Suicide-related event | Has the patient had a suicide-related event? | Suicide | Suicide | ICD10 | | T63.062D | Toxic effect of venom of other North and South American snake, intentional self-harm, subsequent encounter |
| Suicide-related event | Has the patient had a suicide-related event? | Suicide | Suicide | ICD10 | | T63.062S | Toxic effect of venom of other North and South American snake, intentional self-harm, sequela |
| Suicide-related event | Has the patient had a suicide-related event? | Suicide | Suicide | ICD10 | | T63.072A | Toxic effect of venom of other Australian snake, intentional self-harm, initial encounter |
| Suicide-related event | Has the patient had a suicide-related event? | Suicide | Suicide | ICD10 | | T63.072D | Toxic effect of venom of other Australian snake, intentional self-harm, subsequent encounter |
| Suicide-related event | Has the patient had a suicide-related event? | Suicide | Suicide | ICD10 | | T63.072S | Toxic effect of venom of other Australian snake, intentional self-harm, sequela |
| Suicide-related event | Has the patient had a suicide-related event? | Suicide | Suicide | ICD10 | | T63.082A | Toxic effect of venom of other African and Asian snake, intentional self-harm, initial encounter |
| Suicide-related event | Has the patient had a suicide-related event? | Suicide | Suicide | ICD10 | | T63.082D | Toxic effect of venom of other African and Asian snake, intentional self-harm, subsequent encounter |
| Suicide-related event | Has the patient had a suicide-related event? | Suicide | Suicide | ICD10 | | T63.082S | Toxic effect of venom of other African and Asian snake, intentional self-harm, sequela |
| Suicide-related event | Has the patient had a suicide-related event? | Suicide | Suicide | ICD10 | | T63.092A | Toxic effect of venom of other snake, intentional self-harm, initial encounter |
| Suicide-related event | Has the patient had a suicide-related event? | Suicide | Suicide | ICD10 | | T63.092D | Toxic effect of venom of other snake, intentional self-harm, subsequent encounter |
| Suicide-related event | Has the patient had a suicide-related event? | Suicide | Suicide | ICD10 | | T63.092S | Toxic effect of venom of other snake, intentional self-harm, sequela |
| Suicide-related event | Has the patient had a suicide-related event? | Suicide | Suicide | ICD10 | | T63.112A | Toxic effect of venom of gila monster, intentional self-harm, initial encounter |
| Suicide-related event | Has the patient had a suicide-related event? | Suicide | Suicide | ICD10 | | T63.112D | Toxic effect of venom of gila monster, intentional self-harm, subsequent encounter |
| Suicide-related event | Has the patient had a suicide-related event? | Suicide | Suicide | ICD10 | | T63.112S | Toxic effect of venom of gila monster, intentional self-harm, sequela |
| Suicide-related event | Has the patient had a suicide-related event? | Suicide | Suicide | ICD10 | | T63.122A | Toxic effect of venom of other venomous lizard, intentional self-harm, initial encounter |
| Suicide-related event | Has the patient had a suicide-related event? | Suicide | Suicide | ICD10 | | T63.122D | Toxic effect of venom of other venomous lizard, intentional self-harm, subsequent encounter |
| Suicide-related event | Has the patient had a suicide-related event? | Suicide | Suicide | ICD10 | | T63.122S | Toxic effect of venom of other venomous lizard, intentional self-harm, sequela |
| Suicide-related event | Has the patient had a suicide-related event? | Suicide | Suicide | ICD10 | | T63.192A | Toxic effect of venom of other reptiles, intentional self-harm, initial encounter |
| Suicide-related event | Has the patient had a suicide-related event? | Suicide | Suicide | ICD10 | | T63.192D | Toxic effect of venom of other reptiles, intentional self-harm, subsequent encounter |
| Suicide-related event | Has the patient had a suicide-related event? | Suicide | Suicide | ICD10 | | T63.192S | Toxic effect of venom of other reptiles, intentional self-harm, sequela |
| Suicide-related event | Has the patient had a suicide-related event? | Suicide | Suicide | ICD10 | | T63.2X2A | Toxic effect of venom of scorpion, intentional self-harm, initial encounter |
| Suicide-related event | Has the patient had a suicide-related event? | Suicide | Suicide | ICD10 | | T63.2X2D | Toxic effect of venom of scorpion, intentional self-harm, subsequent encounter |
| Suicide-related event | Has the patient had a suicide-related event? | Suicide | Suicide | ICD10 | | T63.2X2S | Toxic effect of venom of scorpion, intentional self-harm, sequela |
| Suicide-related event | Has the patient had a suicide-related event? | Suicide | Suicide | ICD10 | | T63.302A | Toxic effect of unspecified spider venom, intentional self-harm, initial encounter |
| Suicide-related event | Has the patient had a suicide-related event? | Suicide | Suicide | ICD10 | | T63.302D | Toxic effect of unspecified spider venom, intentional self-harm, subsequent encounter |
| Suicide-related event | Has the patient had a suicide-related event? | Suicide | Suicide | ICD10 | | T63.302S | Toxic effect of unspecified spider venom, intentional self-harm, sequela |
| Suicide-related event | Has the patient had a suicide-related event? | Suicide | Suicide | ICD10 | | T63.312A | Toxic effect of venom of black widow spider, intentional self-harm, initial encounter |
| Suicide-related event | Has the patient had a suicide-related event? | Suicide | Suicide | ICD10 | | T63.312D | Toxic effect of venom of black widow spider, intentional self-harm, subsequent encounter |
| Suicide-related event | Has the patient had a suicide-related event? | Suicide | Suicide | ICD10 | | T63.312S | Toxic effect of venom of black widow spider, intentional self-harm, sequela |
| Suicide-related event | Has the patient had a suicide-related event? | Suicide | Suicide | ICD10 | | T63.322A | Toxic effect of venom of tarantula, intentional self-harm, initial encounter |
| Suicide-related event | Has the patient had a suicide-related event? | Suicide | Suicide | ICD10 | | T63.322D | Toxic effect of venom of tarantula, intentional self-harm, subsequent encounter |
| Suicide-related event | Has the patient had a suicide-related event? | Suicide | Suicide | ICD10 | | T63.322S | Toxic effect of venom of tarantula, intentional self-harm, sequela |
| Suicide-related event | Has the patient had a suicide-related event? | Suicide | Suicide | ICD10 | | T63.332A | Toxic effect of venom of brown recluse spider, intentional self-harm, initial encounter |
| Suicide-related event | Has the patient had a suicide-related event? | Suicide | Suicide | ICD10 | | T63.332D | Toxic effect of venom of brown recluse spider, intentional self-harm, subsequent encounter |
| Suicide-related event | Has the patient had a suicide-related event? | Suicide | Suicide | ICD10 | | T63.332S | Toxic effect of venom of brown recluse spider, intentional self-harm, sequela |
| Suicide-related event | Has the patient had a suicide-related event? | Suicide | Suicide | ICD10 | | T63.392A | Toxic effect of venom of other spider, intentional self-harm, initial encounter |
| Suicide-related event | Has the patient had a suicide-related event? | Suicide | Suicide | ICD10 | | T63.392D | Toxic effect of venom of other spider, intentional self-harm, subsequent encounter |
| Suicide-related event | Has the patient had a suicide-related event? | Suicide | Suicide | ICD10 | | T63.392S | Toxic effect of venom of other spider, intentional self-harm, sequela |
| Suicide-related event | Has the patient had a suicide-related event? | Suicide | Suicide | ICD10 | | T63.412A | Toxic effect of venom of centipedes and venomous millipedes, intentional self-harm, initial encounter |
| Suicide-related event | Has the patient had a suicide-related event? | Suicide | Suicide | ICD10 | | T63.412D | Toxic effect of venom of centipedes and venomous millipedes, intentional self-harm, subsequent encounter |
| Suicide-related event | Has the patient had a suicide-related event? | Suicide | Suicide | ICD10 | | T63.412S | Toxic effect of venom of centipedes and venomous millipedes, intentional self-harm, sequela |
| Suicide-related event | Has the patient had a suicide-related event? | Suicide | Suicide | ICD10 | | T63.422A | Toxic effect of venom of ants, intentional self-harm, initial encounter |
| Suicide-related event | Has the patient had a suicide-related event? | Suicide | Suicide | ICD10 | | T63.422D | Toxic effect of venom of ants, intentional self-harm, subsequent encounter |
| Suicide-related event | Has the patient had a suicide-related event? | Suicide | Suicide | ICD10 | | T63.422S | Toxic effect of venom of ants, intentional self-harm, sequela |
| Suicide-related event | Has the patient had a suicide-related event? | Suicide | Suicide | ICD10 | | T63.432A | Toxic effect of venom of caterpillars, intentional self-harm, initial encounter |
| Suicide-related event | Has the patient had a suicide-related event? | Suicide | Suicide | ICD10 | | T63.432D | Toxic effect of venom of caterpillars, intentional self-harm, subsequent encounter |
| Suicide-related event | Has the patient had a suicide-related event? | Suicide | Suicide | ICD10 | | T63.432S | Toxic effect of venom of caterpillars, intentional self-harm, sequela |
| Suicide-related event | Has the patient had a suicide-related event? | Suicide | Suicide | ICD10 | | T63.442A | Toxic effect of venom of bees, intentional self-harm, initial encounter |
| Suicide-related event | Has the patient had a suicide-related event? | Suicide | Suicide | ICD10 | | T63.442D | Toxic effect of venom of bees, intentional self-harm, subsequent encounter |
| Suicide-related event | Has the patient had a suicide-related event? | Suicide | Suicide | ICD10 | | T63.442S | Toxic effect of venom of bees, intentional self-harm, sequela |
| Suicide-related event | Has the patient had a suicide-related event? | Suicide | Suicide | ICD10 | | T63.452A | Toxic effect of venom of hornets, intentional self-harm, initial encounter |
| Suicide-related event | Has the patient had a suicide-related event? | Suicide | Suicide | ICD10 | | T63.452D | Toxic effect of venom of hornets, intentional self-harm, subsequent encounter |
| Suicide-related event | Has the patient had a suicide-related event? | Suicide | Suicide | ICD10 | | T63.452S | Toxic effect of venom of hornets, intentional self-harm, sequela |
| Suicide-related event | Has the patient had a suicide-related event? | Suicide | Suicide | ICD10 | | T63.462A | Toxic effect of venom of wasps, intentional self-harm, initial encounter |
| Suicide-related event | Has the patient had a suicide-related event? | Suicide | Suicide | ICD10 | | T63.462D | Toxic effect of venom of wasps, intentional self-harm, subsequent encounter |
| Suicide-related event | Has the patient had a suicide-related event? | Suicide | Suicide | ICD10 | | T63.462S | Toxic effect of venom of wasps, intentional self-harm, sequela |
| Suicide-related event | Has the patient had a suicide-related event? | Suicide | Suicide | ICD10 | | T63.482A | Toxic effect of venom of other arthropod, intentional self-harm, initial encounter |
| Suicide-related event | Has the patient had a suicide-related event? | Suicide | Suicide | ICD10 | | T63.482D | Toxic effect of venom of other arthropod, intentional self-harm, subsequent encounter |
| Suicide-related event | Has the patient had a suicide-related event? | Suicide | Suicide | ICD10 | | T63.482S | Toxic effect of venom of other arthropod, intentional self-harm, sequela |
| Suicide-related event | Has the patient had a suicide-related event? | Suicide | Suicide | ICD10 | | T63.512A | Toxic effect of contact with stingray, intentional self-harm, initial encounter |
| Suicide-related event | Has the patient had a suicide-related event? | Suicide | Suicide | ICD10 | | T63.512D | Toxic effect of contact with stingray, intentional self-harm, subsequent encounter |
| Suicide-related event | Has the patient had a suicide-related event? | Suicide | Suicide | ICD10 | | T63.512S | Toxic effect of contact with stingray, intentional self-harm, sequela |
| Suicide-related event | Has the patient had a suicide-related event? | Suicide | Suicide | ICD10 | | T63.592A | Toxic effect of contact with other venomous fish, intentional self-harm, initial encounter |
| Suicide-related event | Has the patient had a suicide-related event? | Suicide | Suicide | ICD10 | | T63.592D | Toxic effect of contact with other venomous fish, intentional self-harm, subsequent encounter |
| Suicide-related event | Has the patient had a suicide-related event? | Suicide | Suicide | ICD10 | | T63.592S | Toxic effect of contact with other venomous fish, intentional self-harm, sequela |
| Suicide-related event | Has the patient had a suicide-related event? | Suicide | Suicide | ICD10 | | T63.612A | Toxic effect of contact with Portugese Man-o-war, intentional self-harm, initial encounter |
| Suicide-related event | Has the patient had a suicide-related event? | Suicide | Suicide | ICD10 | | T63.612D | Toxic effect of contact with Portugese Man-o-war, intentional self-harm, subsequent encounter |
| Suicide-related event | Has the patient had a suicide-related event? | Suicide | Suicide | ICD10 | | T63.612S | Toxic effect of contact with Portugese Man-o-war, intentional self-harm, sequela |
| Suicide-related event | Has the patient had a suicide-related event? | Suicide | Suicide | ICD10 | | T63.622A | Toxic effect of contact with other jellyfish, intentional self-harm, initial encounter |
| Suicide-related event | Has the patient had a suicide-related event? | Suicide | Suicide | ICD10 | | T63.622D | Toxic effect of contact with other jellyfish, intentional self-harm, subsequent encounter |
| Suicide-related event | Has the patient had a suicide-related event? | Suicide | Suicide | ICD10 | | T63.622S | Toxic effect of contact with other jellyfish, intentional self-harm, sequela |
| Suicide-related event | Has the patient had a suicide-related event? | Suicide | Suicide | ICD10 | | T63.632A | Toxic effect of contact with sea anemone, intentional self-harm, initial encounter |
| Suicide-related event | Has the patient had a suicide-related event? | Suicide | Suicide | ICD10 | | T63.632D | Toxic effect of contact with sea anemone, intentional self-harm, subsequent encounter |
| Suicide-related event | Has the patient had a suicide-related event? | Suicide | Suicide | ICD10 | | T63.632S | Toxic effect of contact with sea anemone, intentional self-harm, sequela |
| Suicide-related event | Has the patient had a suicide-related event? | Suicide | Suicide | ICD10 | | T63.692A | Toxic effect of contact with other venomous marine animals, intentional self-harm, initial encounter |
| Suicide-related event | Has the patient had a suicide-related event? | Suicide | Suicide | ICD10 | | T63.692D | Toxic effect of contact with other venomous marine animals, intentional self-harm, subsequent encounter |
| Suicide-related event | Has the patient had a suicide-related event? | Suicide | Suicide | ICD10 | | T63.692S | Toxic effect of contact with other venomous marine animals, intentional self-harm, sequela |
| Suicide-related event | Has the patient had a suicide-related event? | Suicide | Suicide | ICD10 | | T63.712A | Toxic effect of contact with venomous marine plant, intentional self-harm, initial encounter |
| Suicide-related event | Has the patient had a suicide-related event? | Suicide | Suicide | ICD10 | | T63.712D | Toxic effect of contact with venomous marine plant, intentional self-harm, subsequent encounter |
| Suicide-related event | Has the patient had a suicide-related event? | Suicide | Suicide | ICD10 | | T63.712S | Toxic effect of contact with venomous marine plant, intentional self-harm, sequela |
| Suicide-related event | Has the patient had a suicide-related event? | Suicide | Suicide | ICD10 | | T63.792A | Toxic effect of contact with other venomous plant, intentional self-harm, initial encounter |
| Suicide-related event | Has the patient had a suicide-related event? | Suicide | Suicide | ICD10 | | T63.792D | Toxic effect of contact with other venomous plant, intentional self-harm, subsequent encounter |
| Suicide-related event | Has the patient had a suicide-related event? | Suicide | Suicide | ICD10 | | T63.792S | Toxic effect of contact with other venomous plant, intentional self-harm, sequela |
| Suicide-related event | Has the patient had a suicide-related event? | Suicide | Suicide | ICD10 | | T63.812A | Toxic effect of contact with venomous frog, intentional self-harm, initial encounter |
| Suicide-related event | Has the patient had a suicide-related event? | Suicide | Suicide | ICD10 | | T63.812D | Toxic effect of contact with venomous frog, intentional self-harm, subsequent encounter |
| Suicide-related event | Has the patient had a suicide-related event? | Suicide | Suicide | ICD10 | | T63.812S | Toxic effect of contact with venomous frog, intentional self-harm, sequela |
| Suicide-related event | Has the patient had a suicide-related event? | Suicide | Suicide | ICD10 | | T63.822A | Toxic effect of contact with venomous toad, intentional self-harm, initial encounter |
| Suicide-related event | Has the patient had a suicide-related event? | Suicide | Suicide | ICD10 | | T63.822D | Toxic effect of contact with venomous toad, intentional self-harm, subsequent encounter |
| Suicide-related event | Has the patient had a suicide-related event? | Suicide | Suicide | ICD10 | | T63.822S | Toxic effect of contact with venomous toad, intentional self-harm, sequela |
| Suicide-related event | Has the patient had a suicide-related event? | Suicide | Suicide | ICD10 | | T63.832A | Toxic effect of contact with other venomous amphibian, intentional self-harm, initial encounter |
| Suicide-related event | Has the patient had a suicide-related event? | Suicide | Suicide | ICD10 | | T63.832D | Toxic effect of contact with other venomous amphibian, intentional self-harm, subsequent encounter |
| Suicide-related event | Has the patient had a suicide-related event? | Suicide | Suicide | ICD10 | | T63.832S | Toxic effect of contact with other venomous amphibian, intentional self-harm, sequela |
| Suicide-related event | Has the patient had a suicide-related event? | Suicide | Suicide | ICD10 | | T63.892A | Toxic effect of contact with other venomous animals, intentional self-harm, initial encounter |
| Suicide-related event | Has the patient had a suicide-related event? | Suicide | Suicide | ICD10 | | T63.892D | Toxic effect of contact with other venomous animals, intentional self-harm, subsequent encounter |
| Suicide-related event | Has the patient had a suicide-related event? | Suicide | Suicide | ICD10 | | T63.892S | Toxic effect of contact with other venomous animals, intentional self-harm, sequela |
| Suicide-related event | Has the patient had a suicide-related event? | Suicide | Suicide | ICD10 | | T63.92XA | Toxic effect of contact with unspecified venomous animal, intentional self-harm, initial encounter |
| Suicide-related event | Has the patient had a suicide-related event? | Suicide | Suicide | ICD10 | | T63.92XD | Toxic effect of contact with unspecified venomous animal, intentional self-harm, subsequent encounter |
| Suicide-related event | Has the patient had a suicide-related event? | Suicide | Suicide | ICD10 | | T63.92XS | Toxic effect of contact with unspecified venomous animal, intentional self-harm, sequela |
| Suicide-related event | Has the patient had a suicide-related event? | Suicide | Suicide | ICD10 | | T64.02XA | Toxic effect of aflatoxin, intentional self-harm, initial encounter |
| Suicide-related event | Has the patient had a suicide-related event? | Suicide | Suicide | ICD10 | | T64.02XD | Toxic effect of aflatoxin, intentional self-harm, subsequent encounter |
| Suicide-related event | Has the patient had a suicide-related event? | Suicide | Suicide | ICD10 | | T64.02XS | Toxic effect of aflatoxin, intentional self-harm, sequela |
| Suicide-related event | Has the patient had a suicide-related event? | Suicide | Suicide | ICD10 | | T64.82XA | Toxic effect of other mycotoxin food contaminants, intentional self-harm, initial encounter |
| Suicide-related event | Has the patient had a suicide-related event? | Suicide | Suicide | ICD10 | | T64.82XD | Toxic effect of other mycotoxin food contaminants, intentional self-harm, subsequent encounter |
| Suicide-related event | Has the patient had a suicide-related event? | Suicide | Suicide | ICD10 | | T64.82XS | Toxic effect of other mycotoxin food contaminants, intentional self-harm, sequela |
| Suicide-related event | Has the patient had a suicide-related event? | Suicide | Suicide | ICD10 | | T65.0X2A | Toxic effect of cyanides, intentional self-harm, initial encounter |
| Suicide-related event | Has the patient had a suicide-related event? | Suicide | Suicide | ICD10 | | T65.0X2D | Toxic effect of cyanides, intentional self-harm, subsequent encounter |
| Suicide-related event | Has the patient had a suicide-related event? | Suicide | Suicide | ICD10 | | T65.0X2S | Toxic effect of cyanides, intentional self-harm, sequela |
| Suicide-related event | Has the patient had a suicide-related event? | Suicide | Suicide | ICD10 | | T65.1X2A | Toxic effect of strychnine and its salts, intentional self-harm, initial encounter |
| Suicide-related event | Has the patient had a suicide-related event? | Suicide | Suicide | ICD10 | | T65.1X2D | Toxic effect of strychnine and its salts, intentional self-harm, subsequent encounter |
| Suicide-related event | Has the patient had a suicide-related event? | Suicide | Suicide | ICD10 | | T65.1X2S | Toxic effect of strychnine and its salts, intentional self-harm, sequela |
| Suicide-related event | Has the patient had a suicide-related event? | Suicide | Suicide | ICD10 | | T65.212A | Toxic effect of chewing tobacco, intentional self-harm, initial encounter |
| Suicide-related event | Has the patient had a suicide-related event? | Suicide | Suicide | ICD10 | | T65.212D | Toxic effect of chewing tobacco, intentional self-harm, subsequent encounter |
| Suicide-related event | Has the patient had a suicide-related event? | Suicide | Suicide | ICD10 | | T65.212S | Toxic effect of chewing tobacco, intentional self-harm, sequela |
| Suicide-related event | Has the patient had a suicide-related event? | Suicide | Suicide | ICD10 | | T65.222A | Toxic effect of tobacco cigarettes, intentional self-harm, initial encounter |
| Suicide-related event | Has the patient had a suicide-related event? | Suicide | Suicide | ICD10 | | T65.222D | Toxic effect of tobacco cigarettes, intentional self-harm, subsequent encounter |
| Suicide-related event | Has the patient had a suicide-related event? | Suicide | Suicide | ICD10 | | T65.222S | Toxic effect of tobacco cigarettes, intentional self-harm, sequela |
| Suicide-related event | Has the patient had a suicide-related event? | Suicide | Suicide | ICD10 | | T65.292A | Toxic effect of other tobacco and nicotine, intentional self-harm, initial encounter |
| Suicide-related event | Has the patient had a suicide-related event? | Suicide | Suicide | ICD10 | | T65.292D | Toxic effect of other tobacco and nicotine, intentional self-harm, subsequent encounter |
| Suicide-related event | Has the patient had a suicide-related event? | Suicide | Suicide | ICD10 | | T65.292S | Toxic effect of other tobacco and nicotine, intentional self-harm, sequela |
| Suicide-related event | Has the patient had a suicide-related event? | Suicide | Suicide | ICD10 | | T65.3X2A | Toxic effect of nitroderivatives and aminoderivatives of benzene and its homologues, intentional self-harm, initial encounter |
| Suicide-related event | Has the patient had a suicide-related event? | Suicide | Suicide | ICD10 | | T65.3X2D | Toxic effect of nitroderivatives and aminoderivatives of benzene and its homologues, intentional self-harm, subsequent encounter |
| Suicide-related event | Has the patient had a suicide-related event? | Suicide | Suicide | ICD10 | | T65.3X2S | Toxic effect of nitroderivatives and aminoderivatives of benzene and its homologues, intentional self-harm, sequela |
| Suicide-related event | Has the patient had a suicide-related event? | Suicide | Suicide | ICD10 | | T65.4X2A | Toxic effect of carbon disulfide, intentional self-harm, initial encounter |
| Suicide-related event | Has the patient had a suicide-related event? | Suicide | Suicide | ICD10 | | T65.4X2D | Toxic effect of carbon disulfide, intentional self-harm, subsequent encounter |
| Suicide-related event | Has the patient had a suicide-related event? | Suicide | Suicide | ICD10 | | T65.4X2S | Toxic effect of carbon disulfide, intentional self-harm, sequela |
| Suicide-related event | Has the patient had a suicide-related event? | Suicide | Suicide | ICD10 | | T65.5X2A | Toxic effect of nitroglycerin and other nitric acids and esters, intentional self-harm, initial encounter |
| Suicide-related event | Has the patient had a suicide-related event? | Suicide | Suicide | ICD10 | | T65.5X2D | Toxic effect of nitroglycerin and other nitric acids and esters, intentional self-harm, subsequent encounter |
| Suicide-related event | Has the patient had a suicide-related event? | Suicide | Suicide | ICD10 | | T65.5X2S | Toxic effect of nitroglycerin and other nitric acids and esters, intentional self-harm, sequela |
| Suicide-related event | Has the patient had a suicide-related event? | Suicide | Suicide | ICD10 | | T65.6X2A | Toxic effect of paints and dyes, not elsewhere classified, intentional self-harm, initial encounter |
| Suicide-related event | Has the patient had a suicide-related event? | Suicide | Suicide | ICD10 | | T65.6X2D | Toxic effect of paints and dyes, not elsewhere classified, intentional self-harm, subsequent encounter |
| Suicide-related event | Has the patient had a suicide-related event? | Suicide | Suicide | ICD10 | | T65.6X2S | Toxic effect of paints and dyes, not elsewhere classified, intentional self-harm, sequela |
| Suicide-related event | Has the patient had a suicide-related event? | Suicide | Suicide | ICD10 | | T65.812A | Toxic effect of latex, intentional self-harm, initial encounter |
| Suicide-related event | Has the patient had a suicide-related event? | Suicide | Suicide | ICD10 | | T65.812D | Toxic effect of latex, intentional self-harm, subsequent encounter |
| Suicide-related event | Has the patient had a suicide-related event? | Suicide | Suicide | ICD10 | | T65.812S | Toxic effect of latex, intentional self-harm, sequela |
| Suicide-related event | Has the patient had a suicide-related event? | Suicide | Suicide | ICD10 | | T65.822A | Toxic effect of harmful algae and algae toxins, intentional self-harm, initial encounter |
| Suicide-related event | Has the patient had a suicide-related event? | Suicide | Suicide | ICD10 | | T65.822D | Toxic effect of harmful algae and algae toxins, intentional self-harm, subsequent encounter |
| Suicide-related event | Has the patient had a suicide-related event? | Suicide | Suicide | ICD10 | | T65.822S | Toxic effect of harmful algae and algae toxins, intentional self-harm, sequela |
| Suicide-related event | Has the patient had a suicide-related event? | Suicide | Suicide | ICD10 | | T65.832A | Toxic effect of fiberglass, intentional self-harm, initial encounter |
| Suicide-related event | Has the patient had a suicide-related event? | Suicide | Suicide | ICD10 | | T65.832D | Toxic effect of fiberglass, intentional self-harm, subsequent encounter |
| Suicide-related event | Has the patient had a suicide-related event? | Suicide | Suicide | ICD10 | | T65.832S | Toxic effect of fiberglass, intentional self-harm, sequela |
| Suicide-related event | Has the patient had a suicide-related event? | Suicide | Suicide | ICD10 | | T65.892A | Toxic effect of other specified substances, intentional self-harm, initial encounter |
| Suicide-related event | Has the patient had a suicide-related event? | Suicide | Suicide | ICD10 | | T65.892D | Toxic effect of other specified substances, intentional self-harm, subsequent encounter |
| Suicide-related event | Has the patient had a suicide-related event? | Suicide | Suicide | ICD10 | | T65.892S | Toxic effect of other specified substances, intentional self-harm, sequela |
| Suicide-related event | Has the patient had a suicide-related event? | Suicide | Suicide | ICD10 | | T65.92XA | Toxic effect of unspecified substance, intentional self-harm, initial encounter |
| Suicide-related event | Has the patient had a suicide-related event? | Suicide | Suicide | ICD10 | | T65.92XD | Toxic effect of unspecified substance, intentional self-harm, subsequent encounter |
| Suicide-related event | Has the patient had a suicide-related event? | Suicide | Suicide | ICD10 | | T65.92XS | Toxic effect of unspecified substance, intentional self-harm, sequela |
| Suicide-related event | Has the patient had a suicide-related event? | Suicide | Suicide | ICD10 | | T71.112A | Asphyxiation due to smothering under pillow, intentional self-harm, initial encounter |
| Suicide-related event | Has the patient had a suicide-related event? | Suicide | Suicide | ICD10 | | T71.112D | Asphyxiation due to smothering under pillow, intentional self-harm, subsequent encounter |
| Suicide-related event | Has the patient had a suicide-related event? | Suicide | Suicide | ICD10 | | T71.112S | Asphyxiation due to smothering under pillow, intentional self-harm, sequela |
| Suicide-related event | Has the patient had a suicide-related event? | Suicide | Suicide | ICD10 | | T71.122A | Asphyxiation due to plastic bag, intentional self-harm, initial encounter |
| Suicide-related event | Has the patient had a suicide-related event? | Suicide | Suicide | ICD10 | | T71.122D | Asphyxiation due to plastic bag, intentional self-harm, subsequent encounter |
| Suicide-related event | Has the patient had a suicide-related event? | Suicide | Suicide | ICD10 | | T71.122S | Asphyxiation due to plastic bag, intentional self-harm, sequela |
| Suicide-related event | Has the patient had a suicide-related event? | Suicide | Suicide | ICD10 | | T71.132A | Asphyxiation due to being trapped in bed linens, intentional self-harm, initial encounter |
| Suicide-related event | Has the patient had a suicide-related event? | Suicide | Suicide | ICD10 | | T71.132D | Asphyxiation due to being trapped in bed linens, intentional self-harm, subsequent encounter |
| Suicide-related event | Has the patient had a suicide-related event? | Suicide | Suicide | ICD10 | | T71.132S | Asphyxiation due to being trapped in bed linens, intentional self-harm, sequela |
| Suicide-related event | Has the patient had a suicide-related event? | Suicide | Suicide | ICD10 | | T71.152A | Asphyxiation due to smothering in furniture, intentional self-harm, initial encounter |
| Suicide-related event | Has the patient had a suicide-related event? | Suicide | Suicide | ICD10 | | T71.152D | Asphyxiation due to smothering in furniture, intentional self-harm, subsequent encounter |
| Suicide-related event | Has the patient had a suicide-related event? | Suicide | Suicide | ICD10 | | T71.152S | Asphyxiation due to smothering in furniture, intentional self-harm, sequela |
| Suicide-related event | Has the patient had a suicide-related event? | Suicide | Suicide | ICD10 | | T71.162A | Asphyxiation due to hanging, intentional self-harm, initial encounter |
| Suicide-related event | Has the patient had a suicide-related event? | Suicide | Suicide | ICD10 | | T71.162D | Asphyxiation due to hanging, intentional self-harm, subsequent encounter |
| Suicide-related event | Has the patient had a suicide-related event? | Suicide | Suicide | ICD10 | | T71.162S | Asphyxiation due to hanging, intentional self-harm, sequela |
| Suicide-related event | Has the patient had a suicide-related event? | Suicide | Suicide | ICD10 | | T71.192A | Asphyxiation due to mechanical threat to breathing due to other causes, intentional self-harm, initial encounter |
| Suicide-related event | Has the patient had a suicide-related event? | Suicide | Suicide | ICD10 | | T71.192D | Asphyxiation due to mechanical threat to breathing due to other causes, intentional self-harm, subsequent encounter |
| Suicide-related event | Has the patient had a suicide-related event? | Suicide | Suicide | ICD10 | | T71.192S | Asphyxiation due to mechanical threat to breathing due to other causes, intentional self-harm, sequela |
| Suicide-related event | Has the patient had a suicide-related event? | Suicide | Suicide | ICD10 | | T71.222A | Asphyxiation due to being trapped in a car trunk, intentional self-harm, initial encounter |
| Suicide-related event | Has the patient had a suicide-related event? | Suicide | Suicide | ICD10 | | T71.222D | Asphyxiation due to being trapped in a car trunk, intentional self-harm, subsequent encounter |
| Suicide-related event | Has the patient had a suicide-related event? | Suicide | Suicide | ICD10 | | T71.222S | Asphyxiation due to being trapped in a car trunk, intentional self-harm, sequela |
| Suicide-related event | Has the patient had a suicide-related event? | Suicide | Suicide | ICD10 | | T71.232A | Asphyxiation due to being trapped in a (discarded) refrigerator, intentional self-harm, initial encounter |
| Suicide-related event | Has the patient had a suicide-related event? | Suicide | Suicide | ICD10 | | T71.232D | Asphyxiation due to being trapped in a (discarded) refrigerator, intentional self-harm, subsequent encounter |
| Suicide-related event | Has the patient had a suicide-related event? | Suicide | Suicide | ICD10 | | T71.232S | Asphyxiation due to being trapped in a (discarded) refrigerator, intentional self-harm, sequela |
| Suicide-related event | Has the patient had a suicide-related event? | Suicide | Suicide | ICD10 | | X71.0XXA | Intentional self-harm by drowning and submersion while in bathtub, initial encounter |
| Suicide-related event | Has the patient had a suicide-related event? | Suicide | Suicide | ICD10 | | X71.0XXD | Intentional self-harm by drowning and submersion while in bathtub, subsequent encounter |
| Suicide-related event | Has the patient had a suicide-related event? | Suicide | Suicide | ICD10 | | X71.0XXS | Intentional self-harm by drowning and submersion while in bathtub, sequela |
| Suicide-related event | Has the patient had a suicide-related event? | Suicide | Suicide | ICD10 | | X71.1XXA | Intentional self-harm by drowning and submersion while in swimming pool, initial encounter |
| Suicide-related event | Has the patient had a suicide-related event? | Suicide | Suicide | ICD10 | | X71.1XXD | Intentional self-harm by drowning and submersion while in swimming pool, subsequent encounter |
| Suicide-related event | Has the patient had a suicide-related event? | Suicide | Suicide | ICD10 | | X71.1XXS | Intentional self-harm by drowning and submersion while in swimming pool, sequela |
| Suicide-related event | Has the patient had a suicide-related event? | Suicide | Suicide | ICD10 | | X71.2XXA | Intentional self-harm by drowning and submersion after jump into swimming pool, initial encounter |
| Suicide-related event | Has the patient had a suicide-related event? | Suicide | Suicide | ICD10 | | X71.2XXD | Intentional self-harm by drowning and submersion after jump into swimming pool, subsequent encounter |
| Suicide-related event | Has the patient had a suicide-related event? | Suicide | Suicide | ICD10 | | X71.2XXS | Intentional self-harm by drowning and submersion after jump into swimming pool, sequela |
| Suicide-related event | Has the patient had a suicide-related event? | Suicide | Suicide | ICD10 | | X71.3XXA | Intentional self-harm by drowning and submersion in natural water, initial encounter |
| Suicide-related event | Has the patient had a suicide-related event? | Suicide | Suicide | ICD10 | | X71.3XXD | Intentional self-harm by drowning and submersion in natural water, subsequent encounter |
| Suicide-related event | Has the patient had a suicide-related event? | Suicide | Suicide | ICD10 | | X71.3XXS | Intentional self-harm by drowning and submersion in natural water, sequela |
| Suicide-related event | Has the patient had a suicide-related event? | Suicide | Suicide | ICD10 | | X71.8XXA | Other intentional self-harm by drowning and submersion, initial encounter |
| Suicide-related event | Has the patient had a suicide-related event? | Suicide | Suicide | ICD10 | | X71.8XXD | Other intentional self-harm by drowning and submersion, subsequent encounter |
| Suicide-related event | Has the patient had a suicide-related event? | Suicide | Suicide | ICD10 | | X71.8XXS | Other intentional self-harm by drowning and submersion, sequela |
| Suicide-related event | Has the patient had a suicide-related event? | Suicide | Suicide | ICD10 | | X71.9XXA | Intentional self-harm by drowning and submersion, unspecified, initial encounter |
| Suicide-related event | Has the patient had a suicide-related event? | Suicide | Suicide | ICD10 | | X71.9XXD | Intentional self-harm by drowning and submersion, unspecified, subsequent encounter |
| Suicide-related event | Has the patient had a suicide-related event? | Suicide | Suicide | ICD10 | | X71.9XXS | Intentional self-harm by drowning and submersion, unspecified, sequela |
| Suicide-related event | Has the patient had a suicide-related event? | Suicide | Suicide | ICD10 | | X72.XXXA | Intentional self-harm by handgun discharge, initial encounter |
| Suicide-related event | Has the patient had a suicide-related event? | Suicide | Suicide | ICD10 | | X72.XXXD | Intentional self-harm by handgun discharge, subsequent encounter |
| Suicide-related event | Has the patient had a suicide-related event? | Suicide | Suicide | ICD10 | | X72.XXXS | Intentional self-harm by handgun discharge, sequela |
| Suicide-related event | Has the patient had a suicide-related event? | Suicide | Suicide | ICD10 | | X73.0XXA | Intentional self-harm by shotgun discharge, initial encounter |
| Suicide-related event | Has the patient had a suicide-related event? | Suicide | Suicide | ICD10 | | X73.0XXD | Intentional self-harm by shotgun discharge, subsequent encounter |
| Suicide-related event | Has the patient had a suicide-related event? | Suicide | Suicide | ICD10 | | X73.0XXS | Intentional self-harm by shotgun discharge, sequela |
| Suicide-related event | Has the patient had a suicide-related event? | Suicide | Suicide | ICD10 | | X73.1XXA | Intentional self-harm by hunting rifle discharge, initial encounter |
| Suicide-related event | Has the patient had a suicide-related event? | Suicide | Suicide | ICD10 | | X73.1XXD | Intentional self-harm by hunting rifle discharge, subsequent encounter |
| Suicide-related event | Has the patient had a suicide-related event? | Suicide | Suicide | ICD10 | | X73.1XXS | Intentional self-harm by hunting rifle discharge, sequela |
| Suicide-related event | Has the patient had a suicide-related event? | Suicide | Suicide | ICD10 | | X73.2XXA | Intentional self-harm by machine gun discharge, initial encounter |
| Suicide-related event | Has the patient had a suicide-related event? | Suicide | Suicide | ICD10 | | X73.2XXD | Intentional self-harm by machine gun discharge, subsequent encounter |
| Suicide-related event | Has the patient had a suicide-related event? | Suicide | Suicide | ICD10 | | X73.2XXS | Intentional self-harm by machine gun discharge, sequela |
| Suicide-related event | Has the patient had a suicide-related event? | Suicide | Suicide | ICD10 | | X73.8XXA | Intentional self-harm by other larger firearm discharge, initial encounter |
| Suicide-related event | Has the patient had a suicide-related event? | Suicide | Suicide | ICD10 | | X73.8XXD | Intentional self-harm by other larger firearm discharge, subsequent encounter |
| Suicide-related event | Has the patient had a suicide-related event? | Suicide | Suicide | ICD10 | | X73.8XXS | Intentional self-harm by other larger firearm discharge, sequela |
| Suicide-related event | Has the patient had a suicide-related event? | Suicide | Suicide | ICD10 | | X73.9XXA | Intentional self-harm by unspecified larger firearm discharge, initial encounter |
| Suicide-related event | Has the patient had a suicide-related event? | Suicide | Suicide | ICD10 | | X73.9XXD | Intentional self-harm by unspecified larger firearm discharge, subsequent encounter |
| Suicide-related event | Has the patient had a suicide-related event? | Suicide | Suicide | ICD10 | | X73.9XXS | Intentional self-harm by unspecified larger firearm discharge, sequela |
| Suicide-related event | Has the patient had a suicide-related event? | Suicide | Suicide | ICD10 | | X74.01XA | Intentional self-harm by airgun, initial encounter |
| Suicide-related event | Has the patient had a suicide-related event? | Suicide | Suicide | ICD10 | | X74.01XD | Intentional self-harm by airgun, subsequent encounter |
| Suicide-related event | Has the patient had a suicide-related event? | Suicide | Suicide | ICD10 | | X74.01XS | Intentional self-harm by airgun, sequela |
| Suicide-related event | Has the patient had a suicide-related event? | Suicide | Suicide | ICD10 | | X74.02XA | Intentional self-harm by paintball gun, initial encounter |
| Suicide-related event | Has the patient had a suicide-related event? | Suicide | Suicide | ICD10 | | X74.02XD | Intentional self-harm by paintball gun, subsequent encounter |
| Suicide-related event | Has the patient had a suicide-related event? | Suicide | Suicide | ICD10 | | X74.02XS | Intentional self-harm by paintball gun, sequela |
| Suicide-related event | Has the patient had a suicide-related event? | Suicide | Suicide | ICD10 | | X74.09XA | Intentional self-harm by other gas, air or spring-operated gun, initial encounter |
| Suicide-related event | Has the patient had a suicide-related event? | Suicide | Suicide | ICD10 | | X74.09XD | Intentional self-harm by other gas, air or spring-operated gun, subsequent encounter |
| Suicide-related event | Has the patient had a suicide-related event? | Suicide | Suicide | ICD10 | | X74.09XS | Intentional self-harm by other gas, air or spring-operated gun, sequela |
| Suicide-related event | Has the patient had a suicide-related event? | Suicide | Suicide | ICD10 | | X74.8XXA | Intentional self-harm by other firearm discharge, initial encounter |
| Suicide-related event | Has the patient had a suicide-related event? | Suicide | Suicide | ICD10 | | X74.8XXD | Intentional self-harm by other firearm discharge, subsequent encounter |
| Suicide-related event | Has the patient had a suicide-related event? | Suicide | Suicide | ICD10 | | X74.8XXS | Intentional self-harm by other firearm discharge, sequela |
| Suicide-related event | Has the patient had a suicide-related event? | Suicide | Suicide | ICD10 | | X74.9XXA | Intentional self-harm by unspecified firearm discharge, initial encounter |
| Suicide-related event | Has the patient had a suicide-related event? | Suicide | Suicide | ICD10 | | X74.9XXD | Intentional self-harm by unspecified firearm discharge, subsequent encounter |
| Suicide-related event | Has the patient had a suicide-related event? | Suicide | Suicide | ICD10 | | X74.9XXS | Intentional self-harm by unspecified firearm discharge, sequela |
| Suicide-related event | Has the patient had a suicide-related event? | Suicide | Suicide | ICD10 | | X75.XXXA | Intentional self-harm by explosive material, initial encounter |
| Suicide-related event | Has the patient had a suicide-related event? | Suicide | Suicide | ICD10 | | X75.XXXD | Intentional self-harm by explosive material, subsequent encounter |
| Suicide-related event | Has the patient had a suicide-related event? | Suicide | Suicide | ICD10 | | X75.XXXS | Intentional self-harm by explosive material, sequela |
| Suicide-related event | Has the patient had a suicide-related event? | Suicide | Suicide | ICD10 | | X76.XXXA | Intentional self-harm by smoke, fire and flames, initial encounter |
| Suicide-related event | Has the patient had a suicide-related event? | Suicide | Suicide | ICD10 | | X76.XXXD | Intentional self-harm by smoke, fire and flames, subsequent encounter |
| Suicide-related event | Has the patient had a suicide-related event? | Suicide | Suicide | ICD10 | | X76.XXXS | Intentional self-harm by smoke, fire and flames, sequela |
| Suicide-related event | Has the patient had a suicide-related event? | Suicide | Suicide | ICD10 | | X77.0XXA | Intentional self-harm by steam or hot vapors, initial encounter |
| Suicide-related event | Has the patient had a suicide-related event? | Suicide | Suicide | ICD10 | | X77.0XXD | Intentional self-harm by steam or hot vapors, subsequent encounter |
| Suicide-related event | Has the patient had a suicide-related event? | Suicide | Suicide | ICD10 | | X77.0XXS | Intentional self-harm by steam or hot vapors, sequela |
| Suicide-related event | Has the patient had a suicide-related event? | Suicide | Suicide | ICD10 | | X77.1XXA | Intentional self-harm by hot tap water, initial encounter |
| Suicide-related event | Has the patient had a suicide-related event? | Suicide | Suicide | ICD10 | | X77.1XXD | Intentional self-harm by hot tap water, subsequent encounter |
| Suicide-related event | Has the patient had a suicide-related event? | Suicide | Suicide | ICD10 | | X77.1XXS | Intentional self-harm by hot tap water, sequela |
| Suicide-related event | Has the patient had a suicide-related event? | Suicide | Suicide | ICD10 | | X77.2XXA | Intentional self-harm by other hot fluids, initial encounter |
| Suicide-related event | Has the patient had a suicide-related event? | Suicide | Suicide | ICD10 | | X77.2XXD | Intentional self-harm by other hot fluids, subsequent encounter |
| Suicide-related event | Has the patient had a suicide-related event? | Suicide | Suicide | ICD10 | | X77.2XXS | Intentional self-harm by other hot fluids, sequela |
| Suicide-related event | Has the patient had a suicide-related event? | Suicide | Suicide | ICD10 | | X77.3XXA | Intentional self-harm by hot household appliances, initial encounter |
| Suicide-related event | Has the patient had a suicide-related event? | Suicide | Suicide | ICD10 | | X77.3XXD | Intentional self-harm by hot household appliances, subsequent encounter |
| Suicide-related event | Has the patient had a suicide-related event? | Suicide | Suicide | ICD10 | | X77.3XXS | Intentional self-harm by hot household appliances, sequela |
| Suicide-related event | Has the patient had a suicide-related event? | Suicide | Suicide | ICD10 | | X77.8XXA | Intentional self-harm by other hot objects, initial encounter |
| Suicide-related event | Has the patient had a suicide-related event? | Suicide | Suicide | ICD10 | | X77.8XXD | Intentional self-harm by other hot objects, subsequent encounter |
| Suicide-related event | Has the patient had a suicide-related event? | Suicide | Suicide | ICD10 | | X77.8XXS | Intentional self-harm by other hot objects, sequela |
| Suicide-related event | Has the patient had a suicide-related event? | Suicide | Suicide | ICD10 | | X77.9XXA | Intentional self-harm by unspecified hot objects, initial encounter |
| Suicide-related event | Has the patient had a suicide-related event? | Suicide | Suicide | ICD10 | | X77.9XXD | Intentional self-harm by unspecified hot objects, subsequent encounter |
| Suicide-related event | Has the patient had a suicide-related event? | Suicide | Suicide | ICD10 | | X77.9XXS | Intentional self-harm by unspecified hot objects, sequela |
| Suicide-related event | Has the patient had a suicide-related event? | Suicide | Suicide | ICD10 | | X78.0XXA | Intentional self-harm by sharp glass, initial encounter |
| Suicide-related event | Has the patient had a suicide-related event? | Suicide | Suicide | ICD10 | | X78.0XXD | Intentional self-harm by sharp glass, subsequent encounter |
| Suicide-related event | Has the patient had a suicide-related event? | Suicide | Suicide | ICD10 | | X78.0XXS | Intentional self-harm by sharp glass, sequela |
| Suicide-related event | Has the patient had a suicide-related event? | Suicide | Suicide | ICD10 | | X78.1XXA | Intentional self-harm by knife, initial encounter |
| Suicide-related event | Has the patient had a suicide-related event? | Suicide | Suicide | ICD10 | | X78.1XXD | Intentional self-harm by knife, subsequent encounter |
| Suicide-related event | Has the patient had a suicide-related event? | Suicide | Suicide | ICD10 | | X78.1XXS | Intentional self-harm by knife, sequela |
| Suicide-related event | Has the patient had a suicide-related event? | Suicide | Suicide | ICD10 | | X78.2XXA | Intentional self-harm by sword or dagger, initial encounter |
| Suicide-related event | Has the patient had a suicide-related event? | Suicide | Suicide | ICD10 | | X78.2XXD | Intentional self-harm by sword or dagger, subsequent encounter |
| Suicide-related event | Has the patient had a suicide-related event? | Suicide | Suicide | ICD10 | | X78.2XXS | Intentional self-harm by sword or dagger, sequela |
| Suicide-related event | Has the patient had a suicide-related event? | Suicide | Suicide | ICD10 | | X78.8XXA | Intentional self-harm by other sharp object, initial encounter |
| Suicide-related event | Has the patient had a suicide-related event? | Suicide | Suicide | ICD10 | | X78.8XXD | Intentional self-harm by other sharp object, subsequent encounter |
| Suicide-related event | Has the patient had a suicide-related event? | Suicide | Suicide | ICD10 | | X78.8XXS | Intentional self-harm by other sharp object, sequela |
| Suicide-related event | Has the patient had a suicide-related event? | Suicide | Suicide | ICD10 | | X78.9XXA | Intentional self-harm by unspecified sharp object, initial encounter |
| Suicide-related event | Has the patient had a suicide-related event? | Suicide | Suicide | ICD10 | | X78.9XXD | Intentional self-harm by unspecified sharp object, subsequent encounter |
| Suicide-related event | Has the patient had a suicide-related event? | Suicide | Suicide | ICD10 | | X78.9XXS | Intentional self-harm by unspecified sharp object, sequela |
| Suicide-related event | Has the patient had a suicide-related event? | Suicide | Suicide | ICD10 | | X79.XXXA | Intentional self-harm by blunt object, initial encounter |
| Suicide-related event | Has the patient had a suicide-related event? | Suicide | Suicide | ICD10 | | X79.XXXD | Intentional self-harm by blunt object, subsequent encounter |
| Suicide-related event | Has the patient had a suicide-related event? | Suicide | Suicide | ICD10 | | X79.XXXS | Intentional self-harm by blunt object, sequela |
| Suicide-related event | Has the patient had a suicide-related event? | Suicide | Suicide | ICD10 | | X80.XXXA | Intentional self-harm by jumping from a high place, initial encounter |
| Suicide-related event | Has the patient had a suicide-related event? | Suicide | Suicide | ICD10 | | X80.XXXD | Intentional self-harm by jumping from a high place, subsequent encounter |
| Suicide-related event | Has the patient had a suicide-related event? | Suicide | Suicide | ICD10 | | X80.XXXS | Intentional self-harm by jumping from a high place, sequela |
| Suicide-related event | Has the patient had a suicide-related event? | Suicide | Suicide | ICD10 | | X81.0XXA | Intentional self-harm by jumping or lying in front of motor vehicle, initial encounter |
| Suicide-related event | Has the patient had a suicide-related event? | Suicide | Suicide | ICD10 | | X81.0XXD | Intentional self-harm by jumping or lying in front of motor vehicle, subsequent encounter |
| Suicide-related event | Has the patient had a suicide-related event? | Suicide | Suicide | ICD10 | | X81.0XXS | Intentional self-harm by jumping or lying in front of motor vehicle, sequela |
| Suicide-related event | Has the patient had a suicide-related event? | Suicide | Suicide | ICD10 | | X81.1XXA | Intentional self-harm by jumping or lying in front of (subway) train, initial encounter |
| Suicide-related event | Has the patient had a suicide-related event? | Suicide | Suicide | ICD10 | | X81.1XXD | Intentional self-harm by jumping or lying in front of (subway) train, subsequent encounter |
| Suicide-related event | Has the patient had a suicide-related event? | Suicide | Suicide | ICD10 | | X81.1XXS | Intentional self-harm by jumping or lying in front of (subway) train, sequela |
| Suicide-related event | Has the patient had a suicide-related event? | Suicide | Suicide | ICD10 | | X81.8XXA | Intentional self-harm by jumping or lying in front of other moving object, initial encounter |
| Suicide-related event | Has the patient had a suicide-related event? | Suicide | Suicide | ICD10 | | X81.8XXD | Intentional self-harm by jumping or lying in front of other moving object, subsequent encounter |
| Suicide-related event | Has the patient had a suicide-related event? | Suicide | Suicide | ICD10 | | X81.8XXS | Intentional self-harm by jumping or lying in front of other moving object, sequela |
| Suicide-related event | Has the patient had a suicide-related event? | Suicide | Suicide | ICD10 | | X82.0XXA | Intentional collision of motor vehicle with other motor vehicle, initial encounter |
| Suicide-related event | Has the patient had a suicide-related event? | Suicide | Suicide | ICD10 | | X82.0XXD | Intentional collision of motor vehicle with other motor vehicle, subsequent encounter |
| Suicide-related event | Has the patient had a suicide-related event? | Suicide | Suicide | ICD10 | | X82.0XXS | Intentional collision of motor vehicle with other motor vehicle, sequela |
| Suicide-related event | Has the patient had a suicide-related event? | Suicide | Suicide | ICD10 | | X82.1XXA | Intentional collision of motor vehicle with train, initial encounter |
| Suicide-related event | Has the patient had a suicide-related event? | Suicide | Suicide | ICD10 | | X82.1XXD | Intentional collision of motor vehicle with train, subsequent encounter |
| Suicide-related event | Has the patient had a suicide-related event? | Suicide | Suicide | ICD10 | | X82.1XXS | Intentional collision of motor vehicle with train, sequela |
| Suicide-related event | Has the patient had a suicide-related event? | Suicide | Suicide | ICD10 | | X82.2XXA | Intentional collision of motor vehicle with tree, initial encounter |
| Suicide-related event | Has the patient had a suicide-related event? | Suicide | Suicide | ICD10 | | X82.2XXD | Intentional collision of motor vehicle with tree, subsequent encounter |
| Suicide-related event | Has the patient had a suicide-related event? | Suicide | Suicide | ICD10 | | X82.2XXS | Intentional collision of motor vehicle with tree, sequela |
| Suicide-related event | Has the patient had a suicide-related event? | Suicide | Suicide | ICD10 | | X82.8XXA | Other intentional self-harm by crashing of motor vehicle, initial encounter |
| Suicide-related event | Has the patient had a suicide-related event? | Suicide | Suicide | ICD10 | | X82.8XXD | Other intentional self-harm by crashing of motor vehicle, subsequent encounter |
| Suicide-related event | Has the patient had a suicide-related event? | Suicide | Suicide | ICD10 | | X82.8XXS | Other intentional self-harm by crashing of motor vehicle, sequela |
| Suicide-related event | Has the patient had a suicide-related event? | Suicide | Suicide | ICD10 | | X83.0XXA | Intentional self-harm by crashing of aircraft, initial encounter |
| Suicide-related event | Has the patient had a suicide-related event? | Suicide | Suicide | ICD10 | | X83.0XXD | Intentional self-harm by crashing of aircraft, subsequent encounter |
| Suicide-related event | Has the patient had a suicide-related event? | Suicide | Suicide | ICD10 | | X83.0XXS | Intentional self-harm by crashing of aircraft, sequela |
| Suicide-related event | Has the patient had a suicide-related event? | Suicide | Suicide | ICD10 | | X83.1XXA | Intentional self-harm by electrocution, initial encounter |
| Suicide-related event | Has the patient had a suicide-related event? | Suicide | Suicide | ICD10 | | X83.1XXD | Intentional self-harm by electrocution, subsequent encounter |
| Suicide-related event | Has the patient had a suicide-related event? | Suicide | Suicide | ICD10 | | X83.1XXS | Intentional self-harm by electrocution, sequela |
| Suicide-related event | Has the patient had a suicide-related event? | Suicide | Suicide | ICD10 | | X83.2XXA | Intentional self-harm by exposure to extremes of cold, initial encounter |
| Suicide-related event | Has the patient had a suicide-related event? | Suicide | Suicide | ICD10 | | X83.2XXD | Intentional self-harm by exposure to extremes of cold, subsequent encounter |
| Suicide-related event | Has the patient had a suicide-related event? | Suicide | Suicide | ICD10 | | X83.2XXS | Intentional self-harm by exposure to extremes of cold, sequela |
| Suicide-related event | Has the patient had a suicide-related event? | Suicide | Suicide | ICD10 | | X83.8XXA | Intentional self-harm by other specified means, initial encounter |
| Suicide-related event | Has the patient had a suicide-related event? | Suicide | Suicide | ICD10 | | X83.8XXD | Intentional self-harm by other specified means, subsequent encounter |
| Suicide-related event | Has the patient had a suicide-related event? | Suicide | Suicide | ICD10 | | X83.8XXS | Intentional self-harm by other specified means, sequela |
| Suicide-related event | Has the patient had a suicide-related event? | Suicide | Suicide | ICD9 | | E950.0 | SUICIDE AND SELF-INFLICTED POISONING BY ANALGESICS, ANTIPYRETICS, AND ANTIRHEUMATICS |
| Suicide-related event | Has the patient had a suicide-related event? | Suicide | Suicide | ICD9 | | E950.01 | SUICIDE ATTEMPTED BY ANALGESICS/ANTIPYRETICS/ANTIRHEUMATICS |
| Suicide-related event | Has the patient had a suicide-related event? | Suicide | Suicide | ICD9 | | E950.02 | SUICIDE ACTUAL BY ANALGESICS/ANTIPYRETICS/ANTIRHEUMATICS |
| Suicide-related event | Has the patient had a suicide-related event? | Suicide | Suicide | ICD9 | | E950.1 | SUICIDE AND SELF-INFLICTED POISONING BY BARBITURATES |
| Suicide-related event | Has the patient had a suicide-related event? | Suicide | Suicide | ICD9 | | E950.11 | SUICIDE ATTEMPTED BY BARBITURATES |
| Suicide-related event | Has the patient had a suicide-related event? | Suicide | Suicide | ICD9 | | E950.12 | SUICIDE ACTUAL BY BARBITURATES |
| Suicide-related event | Has the patient had a suicide-related event? | Suicide | Suicide | ICD9 | | E950.2 | SUICIDE AND SELF-INFLICTED POISONING BY OTHER SEDATIVES AND HYPNOTICS |
| Suicide-related event | Has the patient had a suicide-related event? | Suicide | Suicide | ICD9 | | E950.21 | SUICIDE ATTEMPTED BY OTHER SEDATIVES OR HYPNOTICS |
| Suicide-related event | Has the patient had a suicide-related event? | Suicide | Suicide | ICD9 | | E950.22 | SUICIDE ACTUAL BY OTHER SEDATIVES OR HYPNOTICS |
| Suicide-related event | Has the patient had a suicide-related event? | Suicide | Suicide | ICD9 | | E950.3 | SUICIDE AND SELF-INFLICTED POISONING BY TRANQUILIZERS AND OTHER PSYCHOTROPIC AGENTS |
| Suicide-related event | Has the patient had a suicide-related event? | Suicide | Suicide | ICD9 | | E950.31 | SUICIDE ATTEMPTED BY TRANQUILIZERS AND OTHER PSYCHOTROPHIC AGENTS |
| Suicide-related event | Has the patient had a suicide-related event? | Suicide | Suicide | ICD9 | | E950.32 | SUICIDE ACTUAL BY BY TRANQUILIZERS AND OTHER PSYCHOTROPHIC AGENTS |
| Suicide-related event | Has the patient had a suicide-related event? | Suicide | Suicide | ICD9 | | E950.4 | SUICIDE AND SELF-INFLICTED POISONING BY OTHER SPECIFIED DRUGS AND MEDICINAL SUBSTANCES |
| Suicide-related event | Has the patient had a suicide-related event? | Suicide | Suicide | ICD9 | | E950.41 | SUICIDE ATTEMPTED BY OTHER SPECIFIED DRUGS AND MEDICINAL SUBSTANCES |
| Suicide-related event | Has the patient had a suicide-related event? | Suicide | Suicide | ICD9 | | E950.42 | SUICIDE ACTUAL BY OTHER SPECIFIED DRUGS AND MEDICINAL SUBSTANCES |
| Suicide-related event | Has the patient had a suicide-related event? | Suicide | Suicide | ICD9 | | E950.5 | SUICIDE AND SELF-INFLICTED POISONING BY UNSPECIFIED DRUG OR MEDICINAL SUBSTANCE |
| Suicide-related event | Has the patient had a suicide-related event? | Suicide | Suicide | ICD9 | | E950.51 | SUICIDE ATTEMPTED BY UNSPECIFIED DRUG OR MEDICINAL SUBSTANCE |
| Suicide-related event | Has the patient had a suicide-related event? | Suicide | Suicide | ICD9 | | E950.52 | SUICIDE ACTUAL BY UNSPECIFIED DRUG OR MEDICINAL SUBSTANCE |
| Suicide-related event | Has the patient had a suicide-related event? | Suicide | Suicide | ICD9 | | E950.6 | SUICIDE AND SELF-INFLICTED POISONING BY AGRICULTURAL AND HORTICULTURAL CHEMICAL AND PHARMACEUTICAL PREPARATIONS OTHER |
| Suicide-related event | Has the patient had a suicide-related event? | Suicide | Suicide | ICD9 | | E950.61 | SUICIDE ATTEMPTED BY AGRI/HORTI CHEM/PHARM OTH THAN PLANT FOODS/FERTZ |
| Suicide-related event | Has the patient had a suicide-related event? | Suicide | Suicide | ICD9 | | E950.62 | SUICIDE ACTUAL BY AGRI/HORTI CHEM/PHARM OTH THAN PLANT FOODS/FERTILIZ |
| Suicide-related event | Has the patient had a suicide-related event? | Suicide | Suicide | ICD9 | | E950.7 | SUICIDE AND SELF-INFLICTED POISONING BY CORROSIVE AND CAUSTIC SUBSTANCES |
| Suicide-related event | Has the patient had a suicide-related event? | Suicide | Suicide | ICD9 | | E950.71 | SUICIDE ATTEMPTED BY CORROSIVE AND CAUSTIC SUBSTANCES |
| Suicide-related event | Has the patient had a suicide-related event? | Suicide | Suicide | ICD9 | | E950.72 | SUICIDE ACTUAL BY CORROSIVE AND CAUSTIC SUBSTANCES |
| Suicide-related event | Has the patient had a suicide-related event? | Suicide | Suicide | ICD9 | | E950.8 | SUICIDE AND SELF-INFLICTED POISONING BY ARSENIC AND ITS COMPOUNDS |
| Suicide-related event | Has the patient had a suicide-related event? | Suicide | Suicide | ICD9 | | E950.81 | SUICIDE ATTEMPTED BY ARSENIC AND ITS COMPOUNDS |
| Suicide-related event | Has the patient had a suicide-related event? | Suicide | Suicide | ICD9 | | E950.82 | SUICIDE ACTUAL BY ARSENIC AND ITS COMPOUNDS |
| Suicide-related event | Has the patient had a suicide-related event? | Suicide | Suicide | ICD9 | | E950.9 | SUICIDE AND SELF-INFLICTED POISONING BY OTHER AND UNSPECIFIED SOLID AND LIQUID SUBSTANCES |
| Suicide-related event | Has the patient had a suicide-related event? | Suicide | Suicide | ICD9 | | E950.91 | SUICIDE ATTEMPTED BY OTH/UNSPECIFIED SOLID AND LIQUID SUBSTANCES |
| Suicide-related event | Has the patient had a suicide-related event? | Suicide | Suicide | ICD9 | | E950.92 | SUICIDE ACTUAL BY OTH/UNSPECIFIED SOLID AND LIQUID SUBSTANCES |
| Suicide-related event | Has the patient had a suicide-related event? | Suicide | Suicide | ICD9 | | E951.0 | SUICIDE AND SELF-INFLICTED POISONING BY GAS DISTRIBUTED BY PIPELINE |
| Suicide-related event | Has the patient had a suicide-related event? | Suicide | Suicide | ICD9 | | E951.01 | SUICIDE ATTEMPTED BY GAS DISTRIBUTED BY PIPELINE |
| Suicide-related event | Has the patient had a suicide-related event? | Suicide | Suicide | ICD9 | | E951.02 | SUICIDE ACTUAL BY GAS DISTRIBUTED BY PIPELINE |
| Suicide-related event | Has the patient had a suicide-related event? | Suicide | Suicide | ICD9 | | E951.1 | SUICIDE AND SELF-INFLICTED POISONING BY LIQUEFIED PETROLEUM GAS DISTRIBUTED IN MOBILE CONTAINERS |
| Suicide-related event | Has the patient had a suicide-related event? | Suicide | Suicide | ICD9 | | E951.11 | SUICIDE ATTEMPTED BY LIQUEFIED PETROLEUM GAS DIST IN MOBILE CONTAINER |
| Suicide-related event | Has the patient had a suicide-related event? | Suicide | Suicide | ICD9 | | E951.12 | SUICIDE ACTUAL BY LIQUEFIED PETROLEUM GAS DIST IN MOBILE CONTAINERS |
| Suicide-related event | Has the patient had a suicide-related event? | Suicide | Suicide | ICD9 | | E951.8 | SUICIDE AND SELF-INFLICTED POISONING BY OTHER UTILITY GAS |
| Suicide-related event | Has the patient had a suicide-related event? | Suicide | Suicide | ICD9 | | E951.81 | SUICIDE ATTEMPTED BY OTHER UTILITY GAS |
| Suicide-related event | Has the patient had a suicide-related event? | Suicide | Suicide | ICD9 | | E951.82 | SUICIDE ACTUAL BY OTHER UTILITY GAS |
| Suicide-related event | Has the patient had a suicide-related event? | Suicide | Suicide | ICD9 | | E952.0 | SUICIDE AND SELF-INFLICTED POISONING BY MOTOR VEHICLE EXHAUST GAS |
| Suicide-related event | Has the patient had a suicide-related event? | Suicide | Suicide | ICD9 | | E952.01 | SUICIDE ATTEMPTED BY MOTOR VEHICLE EXHAUST GAS |
| Suicide-related event | Has the patient had a suicide-related event? | Suicide | Suicide | ICD9 | | E952.02 | SUICIDE ACTUAL BY MOTOR VEHICLE EXHAUST GAS |
| Suicide-related event | Has the patient had a suicide-related event? | Suicide | Suicide | ICD9 | | E952.1 | SUICIDE AND SELF-INFLICTED POISONING BY OTHR CARBON MONOXIDE |
| Suicide-related event | Has the patient had a suicide-related event? | Suicide | Suicide | ICD9 | | E952.11 | SUICIDE ATTEMPTED BY OTHER CARBON MONOXIDE |
| Suicide-related event | Has the patient had a suicide-related event? | Suicide | Suicide | ICD9 | | E952.12 | SUICIDE ACTUAL BY OTHER CARBON MONOXIDE |
| Suicide-related event | Has the patient had a suicide-related event? | Suicide | Suicide | ICD9 | | E952.8 | SUICIDE AND SELF-INFLICTED POISONING BY OTHER SPECIFIED GASES AND VAPORS |
| Suicide-related event | Has the patient had a suicide-related event? | Suicide | Suicide | ICD9 | | E952.81 | SUICIDE ATTEMPTED BY OTHER SPECIFIED GASES AND VAPORS |
| Suicide-related event | Has the patient had a suicide-related event? | Suicide | Suicide | ICD9 | | E952.82 | SUICIDE ACTUAL BY OTHER SPECIFIED GASES AND VAPORS |
| Suicide-related event | Has the patient had a suicide-related event? | Suicide | Suicide | ICD9 | | E952.9 | SUICIDE AND SELF-INFLICTED POISONING BY UNSPECIFIED GASES AND VAPORS |
| Suicide-related event | Has the patient had a suicide-related event? | Suicide | Suicide | ICD9 | | E952.91 | SUICIDE ATTEMPTED BY UNSPECIFIED GASES AND VAPORS |
| Suicide-related event | Has the patient had a suicide-related event? | Suicide | Suicide | ICD9 | | E952.92 | SUICIDE ACTUAL BY UNSPECIFIED GASES AND VAPORS |
| Suicide-related event | Has the patient had a suicide-related event? | Suicide | Suicide | ICD9 | | E953.0 | SUICIDE AND SELF-INFLICTED INJURY BY HANGING |
| Suicide-related event | Has the patient had a suicide-related event? | Suicide | Suicide | ICD9 | | E953.01 | SUICIDE ATTEMPTED BY HANGING |
| Suicide-related event | Has the patient had a suicide-related event? | Suicide | Suicide | ICD9 | | E953.02 | SUICIDE ACTUAL BY HANGING |
| Suicide-related event | Has the patient had a suicide-related event? | Suicide | Suicide | ICD9 | | E953.1 | SUICIDE AND SELF-INFLICTED INJURY BY SUFFOCATION BY PLASTIC BAG |
| Suicide-related event | Has the patient had a suicide-related event? | Suicide | Suicide | ICD9 | | E953.11 | SUICIDE ATTEMPTED BY SUFFOCATION USING PLASTIC BAG |
| Suicide-related event | Has the patient had a suicide-related event? | Suicide | Suicide | ICD9 | | E953.12 | SUICIDE ACTUAL BY SUFFOCATION USING PLASTIC BAG |
| Suicide-related event | Has the patient had a suicide-related event? | Suicide | Suicide | ICD9 | | E953.8 | SUICIDE AND SELF-INFLICTED INJURY BY OTHER SPECIFIED MEANS |
| Suicide-related event | Has the patient had a suicide-related event? | Suicide | Suicide | ICD9 | | E953.81 | SUICIDE ATTEMPTED BY OTHER SPECIFIED MEANS |
| Suicide-related event | Has the patient had a suicide-related event? | Suicide | Suicide | ICD9 | | E953.82 | SUICIDE ACTUAL BY OTHER SPECIFIED MEANS |
| Suicide-related event | Has the patient had a suicide-related event? | Suicide | Suicide | ICD9 | | E953.9 | SUICIDE AND SELF-INFLICTED INJURY BY UNSPECIFIED MEANS |
| Suicide-related event | Has the patient had a suicide-related event? | Suicide | Suicide | ICD9 | | E954. | SUICIDE AND SELF-INFLICTED INJURY BY SUBMERSION (DROWNING) |
| Suicide-related event | Has the patient had a suicide-related event? | Suicide | Suicide | ICD9 | | E954.1 | SUICIDE ATTEMPTED BY SUBMERSION (DROWNING) |
| Suicide-related event | Has the patient had a suicide-related event? | Suicide | Suicide | ICD9 | | E954.2 | SUICIDE ACTUAL BY SUBMERSION (DROWNING) |
| Suicide-related event | Has the patient had a suicide-related event? | Suicide | Suicide | ICD9 | | E955.0 | SUICIDE AND SELF-INFLICTED INJURY BY HANDGUN |
| Suicide-related event | Has the patient had a suicide-related event? | Suicide | Suicide | ICD9 | | E955.01 | SUICIDE ATTEMPTED BY HANDGUN |
| Suicide-related event | Has the patient had a suicide-related event? | Suicide | Suicide | ICD9 | | E955.02 | SUICIDE ACTUAL BY HANDGUN |
| Suicide-related event | Has the patient had a suicide-related event? | Suicide | Suicide | ICD9 | | E955.1 | SUICIDE AND SELF-INFLICTED INJURY BY SHOTGUN |
| Suicide-related event | Has the patient had a suicide-related event? | Suicide | Suicide | ICD9 | | E955.11 | SUICIDE ATTEMPTED BY SHOTGUN |
| Suicide-related event | Has the patient had a suicide-related event? | Suicide | Suicide | ICD9 | | E955.12 | SUICIDE ACTUAL BY SHOTGUN |
| Suicide-related event | Has the patient had a suicide-related event? | Suicide | Suicide | ICD9 | | E955.2 | SUICIDE AND SELF-INFLICTED INJURY BY HUNTING RIFLE |
| Suicide-related event | Has the patient had a suicide-related event? | Suicide | Suicide | ICD9 | | E955.21 | SUICIDE ATTEMPTED BY HUNTING RIFLE |
| Suicide-related event | Has the patient had a suicide-related event? | Suicide | Suicide | ICD9 | | E955.22 | SUICIDE ACTUAL BY HUNTING RIFLE |
| Suicide-related event | Has the patient had a suicide-related event? | Suicide | Suicide | ICD9 | | E955.3 | SUICIDE AND SELF-INFLICTED INJURY BY MILITARY FIREARMS |
| Suicide-related event | Has the patient had a suicide-related event? | Suicide | Suicide | ICD9 | | E955.31 | SUICIDE ATTEMPTED BY MILITARY FIREARMS |
| Suicide-related event | Has the patient had a suicide-related event? | Suicide | Suicide | ICD9 | | E955.32 | SUICIDE ACTUAL BY MILITARY FIREARMS |
| Suicide-related event | Has the patient had a suicide-related event? | Suicide | Suicide | ICD9 | | E955.4 | SUICIDE AND SELF-INFLICTED INJURY BY OTHER AND UNSPECIFIED FIREARM |
| Suicide-related event | Has the patient had a suicide-related event? | Suicide | Suicide | ICD9 | | E955.41 | SUICIDE ATTEMPTED BY OTHER AND UNSPECIFIED FIREARM |
| Suicide-related event | Has the patient had a suicide-related event? | Suicide | Suicide | ICD9 | | E955.42 | SUICIDE ACTUAL BY OTHER AND UNSPECIFIED FIREARM |
| Suicide-related event | Has the patient had a suicide-related event? | Suicide | Suicide | ICD9 | | E955.5 | SUICIDE AND SELF-INFLICTED INJURY BY EXPLOSIVES |
| Suicide-related event | Has the patient had a suicide-related event? | Suicide | Suicide | ICD9 | | E955.51 | SUICIDE ATTEMPTED BY EXPLOSIVES |
| Suicide-related event | Has the patient had a suicide-related event? | Suicide | Suicide | ICD9 | | E955.52 | SUICIDE ACTUAL BY EXPLOSIVES |
| Suicide-related event | Has the patient had a suicide-related event? | Suicide | Suicide | ICD9 | | E955.6 | SUICIDE AND SELF-INFLICTED INJURY BY AIR GUN |
| Suicide-related event | Has the patient had a suicide-related event? | Suicide | Suicide | ICD9 | | E955.7 | SUICIDE AND SELF-INFLICTED INJURY BY FIREARMS, AIR GUNS AND EXPLOSIVES, PAINTBALL GUN |
| Suicide-related event | Has the patient had a suicide-related event? | Suicide | Suicide | ICD9 | | E955.9 | SUICIDE AND SELF-INFLICTED INJURY BY FIREARMS AND EXPLOSIVES, UNSPECIFIED |
| Suicide-related event | Has the patient had a suicide-related event? | Suicide | Suicide | ICD9 | | E956. | SUICIDE AND SELF-INFLICTED INJURY BY CUTTING AND PIERCING INSTRUMENT |
| Suicide-related event | Has the patient had a suicide-related event? | Suicide | Suicide | ICD9 | | E956.1 | SUICIDE ATTEMPTED BY CUTTING AND PIERCING INSTRUMENT |
| Suicide-related event | Has the patient had a suicide-related event? | Suicide | Suicide | ICD9 | | E956.2 | SUICIDE ACTUAL BY CUTTING AND PIERCING INSTRUMENT |
| Suicide-related event | Has the patient had a suicide-related event? | Suicide | Suicide | ICD9 | | E957.0 | SUICIDE AND SELF-INFLICTED INJURIES BY JUMPING FROM RESIDENTIAL PREMISES |
| Suicide-related event | Has the patient had a suicide-related event? | Suicide | Suicide | ICD9 | | E957.01 | SUICIDE ATTEMPTED BY JUMPING FROM RESIDENTIAL PREMISES |
| Suicide-related event | Has the patient had a suicide-related event? | Suicide | Suicide | ICD9 | | E957.02 | SUICIDE ACTUAL BY JUMPING FROM RESIDENTIAL PREMISES |
| Suicide-related event | Has the patient had a suicide-related event? | Suicide | Suicide | ICD9 | | E957.1 | SUICIDE AND SELF-INFLICTED INJURIES BY JUMPING FROM OTHER MAN-MADE STRUCTURES |
| Suicide-related event | Has the patient had a suicide-related event? | Suicide | Suicide | ICD9 | | E957.11 | SUICIDE ATTEMPTED BY JUMPING FROM OTHER MAN-MADE STRUCTURES |
| Suicide-related event | Has the patient had a suicide-related event? | Suicide | Suicide | ICD9 | | E957.12 | SUICIDE ACTUAL BY JUMPING FROM OTHER MAN-MADE STRUCTURES |
| Suicide-related event | Has the patient had a suicide-related event? | Suicide | Suicide | ICD9 | | E957.2 | SUICIDE AND SELF-INFLICTED INJURIES BY JUMPING FROM NATURAL SITES |
| Suicide-related event | Has the patient had a suicide-related event? | Suicide | Suicide | ICD9 | | E957.21 | SUICIDE ATTEMPTED BY JUMPING FROM NATURAL SITES |
| Suicide-related event | Has the patient had a suicide-related event? | Suicide | Suicide | ICD9 | | E957.22 | SUICIDE ACTUAL BY JUMPING FROM NATURAL SITES |
| Suicide-related event | Has the patient had a suicide-related event? | Suicide | Suicide | ICD9 | | E957.9 | SUICIDE AND SELF-INFLICTED INJURIES BY JUMPING FROM UNSPECIFIED SITE |
| Suicide-related event | Has the patient had a suicide-related event? | Suicide | Suicide | ICD9 | | E957.91 | SUICIDE ATTEMPTED BY JUMPING FROM UNSPECIFIED HIGH PLACE |
| Suicide-related event | Has the patient had a suicide-related event? | Suicide | Suicide | ICD9 | | E957.92 | SUICIDE ACTUAL BY JUMPING FROM UNSPECIFIED HIGH PLACE |
| Suicide-related event | Has the patient had a suicide-related event? | Suicide | Suicide | ICD9 | | E958.0 | SUICIDE AND SELF-INFLICTED INJURY BY JUMPING OR LYING BEFORE MOVING OBJECT |
| Suicide-related event | Has the patient had a suicide-related event? | Suicide | Suicide | ICD9 | | E958.01 | SUICIDE ATTEMPTED BY JUMPING OR LYING BEFORE MOVING OBJECT |
| Suicide-related event | Has the patient had a suicide-related event? | Suicide | Suicide | ICD9 | | E958.02 | SUICIDE ACTUAL BY JUMPING OR LYING BEFORE MOVING OBJECT |
| Suicide-related event | Has the patient had a suicide-related event? | Suicide | Suicide | ICD9 | | E958.1 | SUICIDE AND SELF-INFLICTED INJURY BY BURNS, FIRE |
| Suicide-related event | Has the patient had a suicide-related event? | Suicide | Suicide | ICD9 | | E958.11 | SUICIDE ATTEMPTED BY BURNS OR FIRE |
| Suicide-related event | Has the patient had a suicide-related event? | Suicide | Suicide | ICD9 | | E958.12 | SUICIDE ACTUAL BY BURNS OR FIRE |
| Suicide-related event | Has the patient had a suicide-related event? | Suicide | Suicide | ICD9 | | E958.2 | SUICIDE AND SELF-INFLICTED INJURY BY SCALD |
| Suicide-related event | Has the patient had a suicide-related event? | Suicide | Suicide | ICD9 | | E958.21 | SUICIDE ATTEMPTED BY SCALD |
| Suicide-related event | Has the patient had a suicide-related event? | Suicide | Suicide | ICD9 | | E958.22 | SUICIDE ACTUAL BY SCALD |
| Suicide-related event | Has the patient had a suicide-related event? | Suicide | Suicide | ICD9 | | E958.3 | SUICIDE AND SELF-INFLICTED INJURY BY EXTREMES OF COLD |
| Suicide-related event | Has the patient had a suicide-related event? | Suicide | Suicide | ICD9 | | E958.31 | SUICIDE ATTEMPTED BY EXTREMES OF COLD |
| Suicide-related event | Has the patient had a suicide-related event? | Suicide | Suicide | ICD9 | | E958.32 | SUICIDE ACTUAL BY EXTREMES OF COLD |
| Suicide-related event | Has the patient had a suicide-related event? | Suicide | Suicide | ICD9 | | E958.4 | SUICIDE AND SELF-INFLICTED INJURY BY ELECTROCUTION |
| Suicide-related event | Has the patient had a suicide-related event? | Suicide | Suicide | ICD9 | | E958.41 | SUICIDE ATTEMPTED BY ELECTROCUTION |
| Suicide-related event | Has the patient had a suicide-related event? | Suicide | Suicide | ICD9 | | E958.42 | SUICIDE ACTUAL BY ELECTROCUTION |
| Suicide-related event | Has the patient had a suicide-related event? | Suicide | Suicide | ICD9 | | E958.5 | SUICIDE AND SELF-INFLICTED INJURY BY CRASHING OF MOTOR VEHICLE |
| Suicide-related event | Has the patient had a suicide-related event? | Suicide | Suicide | ICD9 | | E958.51 | SUICIDE ATTEMPTED BY CRASHING OF MOTOR VEHICLE |
| Suicide-related event | Has the patient had a suicide-related event? | Suicide | Suicide | ICD9 | | E958.52 | SUICIDE ACTUAL BY CRASHING OF MOTOR VEHICLE |
| Suicide-related event | Has the patient had a suicide-related event? | Suicide | Suicide | ICD9 | | E958.6 | SUICIDE AND SELF-INFLICTED INJURY BY CRASHING OF AIRCRAFT |
| Suicide-related event | Has the patient had a suicide-related event? | Suicide | Suicide | ICD9 | | E958.61 | SUICIDE ATTEMPTED BY CRASHING OF AIRCRAFT |
| Suicide-related event | Has the patient had a suicide-related event? | Suicide | Suicide | ICD9 | | E958.62 | SUICIDE ACTUAL BY CRASHING OF AIRCRAFT |
| Suicide-related event | Has the patient had a suicide-related event? | Suicide | Suicide | ICD9 | | E958.7 | SUICIDE AND SELF-INFLICTED INJURY BY CAUSTIC SUBSTANCES, EXCEPT POISONING |
| Suicide-related event | Has the patient had a suicide-related event? | Suicide | Suicide | ICD9 | | E958.71 | SUICIDE ATTEMPTED BY CAUSTIC SUBSTANCES EXCEPT POISONINGS |
| Suicide-related event | Has the patient had a suicide-related event? | Suicide | Suicide | ICD9 | | E958.72 | SUICIDE ACTUAL BY CAUSTIC SUBSTANCES EXCEPT POISONINGS |
| Suicide-related event | Has the patient had a suicide-related event? | Suicide | Suicide | ICD9 | | E958.8 | SUICIDE AND SELF-INFLICTED INJURY BY OTHER SPECIFIED MEANS |
| Suicide-related event | Has the patient had a suicide-related event? | Suicide | Suicide | ICD9 | | E958.81 | SUICIDE ATTEMPTED BY OTHER SPECIFIC MEANS |
| Suicide-related event | Has the patient had a suicide-related event? | Suicide | Suicide | ICD9 | | E958.82 | SUICIDE ACTUAL BY OTHER SPECIFIC MEANS |
| Suicide-related event | Has the patient had a suicide-related event? | Suicide | Suicide | ICD9 | | E958.9 | SUICIDE AND SELF-INFLICTED INJURY BY UNSPECIFIED MEANS |
| Suicide-related event | Has the patient had a suicide-related event? | Suicide | Suicide | ICD9 | | E959. | LATE EFFECTS OF SELF-INFLICTED INJURY |
| Suicide-related event | Has the patient had a suicide-related event? | Suicide | Suicide | ICD9 | | E980.6 | POISONING BY CORROSIVE AND CAUSTIC SUBSTANCES, UNDETERMINED WHETHER ACCIDENTALLY OR PURPOSELY INFLICTED |
| Suicide-related event | Has the patient had a suicide-related event? | Suicide | Suicide | ICD9 | | E980.8 | POISONING BY ARSENIC AND ITS COMPOUNDS, UNDETERMINED WHETHER ACCIDENTALLY OR PURPOSELY INFLICTED |
| Suicide-related event | Has the patient had a suicide-related event? | Suicide | Suicide | ICD9 | | E981.0 | POISONING BY GAS DISTRIBUTED BY PIPELINE, UNDETERMINED WHETHER ACCIDENTALLY OR PURPOSELY INFLICTED |
| Suicide-related event | Has the patient had a suicide-related event? | Suicide | Suicide | ICD9 | | E981.1 | POISONING BY LIQUEFIED PETROLEUM GAS DISTRIBUTED IN MOBILE CONTAINERS, UNDETERMINED WHETHER ACCIDENTALLY OR PURPOSELY |
| Suicide-related event | Has the patient had a suicide-related event? | Suicide | Suicide | ICD9 | | E981.8 | POISONING BY OTHER UTILITY GAS, UNDETERMINED WHETHER ACCIDENTALLY OR PURPOSELY INFLICTED |
| Suicide-related event | Has the patient had a suicide-related event? | Suicide | Suicide | ICD9 | | E982.0 | POISONING BY MOTOR VEHICLE EXHAUST GAS, UNDETERMINED WHETHER ACCIDENTALLY OR PURPOSELY INFLICTED |
| Suicide-related event | Has the patient had a suicide-related event? | Suicide | Suicide | ICD9 | | E982.1 | POISONING BY OTHER CARBON MONOXIDE, UNDETERMINED WHETHER ACCIDENTALLY OR PURPOSELY INFLICTED |
| Suicide-related event | Has the patient had a suicide-related event? | Suicide | Suicide | ICD9 | | E982.8 | POISONING BY OTHER SPECIFIED GASES AND VAPORS, UNDETERMINED WHETHER ACCIDENTALLY OR PURPOSELY INFLICTED |
| Suicide-related event | Has the patient had a suicide-related event? | Suicide | Suicide | ICD9 | | E982.9 | POISONING BY UNSPECIFIED GASES AND VAPORS, UNDETERMINED WHETHER ACCIDENTALLY OR PURPOSELY INFLICTED |
| Suicide-related event | Has the patient had a suicide-related event? | Suicide | Suicide | ICD9 | | E983.0 | HANGING, UNDETERMINED WHETHER ACCIDENTALLY OR PURPOSELY INFLICTED |
| Suicide-related event | Has the patient had a suicide-related event? | Suicide | Suicide | ICD9 | | E983.1 | SUFFOCATION BY PLASTIC BAG, UNDETERMINED WHETHER ACCIDENTALLY OR PURPOSELY INFLICTED |
| Suicide-related event | Has the patient had a suicide-related event? | Suicide | Suicide | ICD9 | | E983.8 | STRANGULATION OR SUFFOCATION BY OTHER SPECIFIED MEANS, UNDETERMINED WHETHER ACCIDENTALLY OR PURPOSELY INFLICTED |
| Suicide-related event | Has the patient had a suicide-related event? | Suicide | Suicide | ICD9 | | E983.9 | STRANGULATION OR SUFFOCATION BY UNSPECIFIED MEANS, UNDETERMINED WHETHER ACCIDENTALLY OR PURPOSELY INFLICTED |
| Suicide-related event | Has the patient had a suicide-related event? | Suicide | Suicide | ICD9 | | E984. | SUBMERSION (DROWNING), UNDETERMINED WHETHER ACCIDENTALLY OR PURPOSELY INFLICTED |
| Suicide-related event | Has the patient had a suicide-related event? | Suicide | Suicide | ICD9 | | E988.0 | INJURY BY JUMPING OR LYING BEFORE MOVING OBJECT, UNDETERMINED WHETHER ACCIDENTALLY OR PURPOSELY INFLICTED |
| Suicide-related event | Has the patient had a suicide-related event? | Suicide | Suicide | ICD9 | | E988.1 | INJURY BY BURNS OR FIRE, UNDETERMINED WHETHER ACCIDENTALLY OR PURPOSELY INFLICTED |
| Suicide-related event | Has the patient had a suicide-related event? | Suicide | Suicide | ICD9 | | E988.2 | INJURY BY SCALD, UNDETERMINED WHETHER ACCIDENTALLY OR PURPOSELY INFLICTED |
| Suicide-related event | Has the patient had a suicide-related event? | Suicide | Suicide | ICD9 | | E988.3 | INJURY BY EXTREMES OF COLD, UNDETERMINED WHETHER ACCIDENTALLY OR PURPOSELY INFLICTED |
| Suicide-related event | Has the patient had a suicide-related event? | Suicide | Suicide | ICD9 | | E988.4 | INJURY BY ELECTROCUTION, UNDETERMINED WHETHER ACCIDENTALLY OR PURPOSELY INFLICTED |
| Suicide-related event | Has the patient had a suicide-related event? | Suicide | Suicide | ICD9 | | E988.5 | INJURY BY CRASHING OF MOTOR VEHICLE, UNDETERMINED WHETHER ACCIDENTALLY OR PURPOSELY INFLICTED |
| Suicide-related event | Has the patient had a suicide-related event? | Suicide | Suicide | ICD9 | | E988.6 | INJURY BY CRASHING OF AIRCRAFT, UNDETERMINED WHETHER ACCIDENTALLY OR PURPOSELY INFLICTED |
| Suicide-related event | Has the patient had a suicide-related event? | Suicide | Suicide | ICD9 | | E988.7 | INJURY BY CAUSTIC SUBSTANCES, EXCEPT POISONING, UNDETERMINED WHETHER ACCIDENTALLY OR PURPOSELY INFLICTED |
| Suicide-related event | Has the patient had a suicide-related event? | Suicide | Suicide | ICD9 | | E988.8 | INJURY BY OTHER SPECIFIED MEANS, UNDETERMINED WHETHER ACCIDENTALLY OR PURPOSELY INFLICTED |
| Suicide-related event | Has the patient had a suicide-related event? | Suicide | Suicide | ICD9 | | E988.9 | INJURY BY UNSPECIFIED MEANS, UNDETERMINED WHETHER ACCIDENTALLY OR PURPOSELY INFLICTED |
| Suicide-related event | Has the patient had a suicide-related event? | Suicide | Suicide | ICD9 | | V62.84 | SUICIDAL IDEATION |
