## Appendix B for "A Natural Language Processing Algorithm for Classifying Suicidal Behaviors in Alzheimer’s Disease and Related Dementia Patients: Development and Validation Using Electronic Health Records Data"

| ICD-10 | Definition | ICD-9 | Definition |
| --- | --- | --- | --- |
| G30.0 | Alzheimer's disease with early onset | 290.12 | Presenile dementia with delusional features |
| G30.1 | Alzheimer's disease with late onset | 331 | Alzheimer's disease |
| G30.8 | Other Alzheimer's disease | 290.4 | Vascular dementia, uncomplicated |
| G30.9 | Alzheimer's disease, unspecified | 290.41 | Vascular dementia, with delirium |
| F02.80 | Dementia in other diseases classified elsewhere without behavioral disturbance | 290.42 | Vascular dementia, with delusions |
| F02.81 | Dementia in other diseases classified elsewhere with | 290.43 | Vascular dementia, with depressed mood |
|  | behavioral disturbance, unspecifieed severity | 294 | Amnestic disorder in conditions classified elsewhere |
| F02.90 | Unspecified dementia without behavioral disturbance | 294.8 | Other persistent mental disorders due to conditions classified elsewhere |
| F03.91 | Unspecified dementia with behavioral disturbance | 294.9 | Unspecified persistent mental disorders due to conditions classified elsewhere |
| F04 | Amnestic disorder due to known physiological condition | 294.1 | Dementia in conditions classified elsewhere without behavioral disturbance |
| F06.0 | Psychotic disorder with hallucinations due to known physiological condition | 294.11 | Dementia in conditions classified elsewhere with behavioral disturbance |
| F06.8 | Other specified mental disorders due to known physiological condition | 294.2 | Dementia, unspecified, without behavioral disturbance |
| G31.01 | Pick's disease | 294.21 | Dementia, unspecified, with behavioral disturbance |
| G31.09 | Other frontotemporaldementia | 331.2 | Senile degeneration of brain |
| G31.1 | Senile degeneration of brain, not elsewhere classified | 331.6 | Corticobasal degeneration |
| G31.83 | Dementia with Lewy bodies | 331.7 | Cerebral degeneration in diseases classified elsewhere |
| G31.85 | Corticobasal degeneration | 331.11 | Pick's disease |
| G31.89 | Other specified degenerative diseases of nervous system | 331.19 | Other frontotemporal dementia |
| G31.9 | Degenerative disease of nervous system, unspecified | 331.81 | Reye's syndrome |
| G45.4 | Transient global amnesia | 331.82 | Dementia with lewy bodies |
| G93.7 | Reye's syndrome | 331.89 | Other cerebral degeneration |
| G94 | Other disorders of brain in diseases classified elsewhere | 437.7 | Transient global amnesia |
| G91.0 | Communicating hydrocephalus | 331.3 | Communicating hydrocephalus |
| G91.1 | Obstructive hydrocephalus | 331.4 | Obstructive hydrocephalus |
| G91.2 | (Idiopathic) normal pressure hydrocephalus | 331.5 | Idiopathic normal pressure hydrocephalus (INPH) |
| F01.50 | Vascular dementia without behavioral disturbance | 437 | Cerebral atherosclerosis |
| F01.51 | Vascular dementia with behavioral disturbance | 437.1 | Other generalized ischernic cerebrovascular disease |
| I67.5 | Moyamoya disease | 437.2 | Hypertensive encephalopathy |
| I67.1 | Cerebral aneurysm, nonruptured | 437.3 | Cerebral aneurysm, nonruptured |
| I67.2 | Cerebral atherosclerosis | 437.4 | Cerebral arteritis |
| I67.4 | Hypertensive encephalopathy | 437.5 | Moyamoya disease |
| I67.6 | Nonpyogenic thrombosis ofintracranialvenous system | 437.6 | Nonpyogenic thrombosis of intracranial venous sinus |
| I67.7 | Cerebral arteritis, not elsewhere classified | 437.8 | Other ill-defined cerebrovascular disease |
| I67.81 | Acute cerebrovascular insufficiency | 437.9 | Unspecified cerebrovascular disease |
| I67.82 | Cerebral ischemia | 331.83 | Mild cognitive impairment |
| I67.89 | Other cerebrovascular disease | 331 | Alzheimer’s disease |
| I67.9 | Cerebrovascular disease, unspecified | 331.1 | frontotemporal dementia |
