## Supplementary material for "A Natural Language Processing Algorithm for Classifying Suicidal Behaviors in Alzheimer’s Disease and Related Dementia Patients: Development and Validation Using Electronic Health Records Data": Table 3

Table 3:Performance of machine learning, and deep learning classifiers of ADRD patient population extracted from MIMIC dataset documented in unstructured narrative clinical notes.

| **Class** | **LR** | | | **SVM** | | | **Decision Tree** | | | **Random Forest** | | | **BERT** | | | **RoBERTa** | | | **Clinical BERT** | | | **Support (#N)** |
| --- | --- | --- | --- | --- | --- | --- | --- | --- | --- | --- | --- | --- | --- | --- | --- | --- | --- | --- | --- | --- | --- | --- |
|  | P† | R* | F§ | P† | R* | F§ | P† | R* | F§ | P† | R* | F§ | P† | R* | F§ | P† | R* | F§ | P† | R* | F§ |  |
| Suicide attempt | .74 | .78 | .76 | .75 | .82 | .78 | .78 | .83 | .80 | .82 | .72 | .77 | .99 | .98 | .99 | .98 | 1 | .99 | .97 | 1 | .98 | 129 |
| Passive suicidal ideation | .77 | .54 | .64 | .81 | .66 | .32 | .63 | .49 | .55 | .77 | .41 | .53 | 1 | .97 | .99 | .99 | .97 | .98 | 1 | .96 | .98 | 116 |
| Active suicidal ideation | .92 | .55 | .69 | .90 | .45 | .60 | .71 | .50 | .59 | 86 | .30 | .44 | .95 | 1 | .98 | 1 | .95 | .97 | 1 | 1 | 1 | 20 |
| Non-suicidal | .78 | .52 | .62 | .72 | .65 | .69 | .48 | .68 | .56 | .61 | .68 | .65 | .96 | .99 | .97 | .99 | 1 | .99 | .96 | .99 | .97 | 75 |
| NSSI | .0 | .0 | .0 | .50 | .15 | .24 | .56 | .38 | .45 | 1 | .08 | .14 | 1 | 1 | 1 | 1 | .92 | .96 | .91 | .77 | .83 | 13 |
| Mental disease | .60 | .66 | .63 | .66 | .58 | .62 | .47 | .59 | .52 | .68 | .38 | .49 | .95 | .96 | .96 | .91 | .95 | .93 | 1 | .97 | .99 | 79 |
| Overdose | .83 | .51 | .63 | .81 | .66 | .73 | .90 | .89 | .89 | .96 | .49 | .65 | 1 | .99 | .99 | .95 | .97 | .96 | .99 | .99 | .99 | 97 |
| Irrelevant | .81 | .93 | .87 | .83 | .93 | .88 | .86 | .89 | .88 | .74 | .96 | .83 | .98 | .99 | .98 | .98 | .98 | .98 | .96 | .99 | .99 | 419 |
| Micro Avg | .78 | .74 | .76 | .79 | .78 | .79 | .75 | .78 | .76 | .75 | .89 | .73 | .98 | .98 | .98 | .97 | .98 | .97 | .98 | .98 | .98 | 948 |
| Macro Avg | .68 | .56 | .60 | .75 | .61 | .66 | .67 | .66 | .66 | .80 | .89 | .56 | .98 | .99 | .98 | .87 | .86 | .86 | .98 | .96 | .97 | 948 |
| Weighted Avg | .77 | .74 | .75 | .78 | .78 | .78 | .75 | .78 | .76 | .77 | .89 | .71 | .98 | .98 | .98 | .97 | .98 | .97 | .98 | .98 | .98 | 948 |

- *Recall/ sensitivity.
- †Precision/ positive predictive value.
- §F score = 2 x [(Precision x Recall) / (Precision + Recall)].
- #Number of snippets included in model evaluation.
- Abbreviations: BERT, Bidirectional Encoder Representations from Transformers; DT, Decision Tree; LR, logistic regression; L-SVM, linear support vector machines; NSSI, non-suicidal self-harm injur
