## Supplementary material for "A Natural Language Processing Algorithm for Classifying Suicidal Behaviors in Alzheimer’s Disease and Related Dementia Patients: Development and Validation Using Electronic Health Records Data": Table 2

Table 2: The frequency and percentage of occurrence of each label in relevant subset and the document

| Labels | | Frequency of occurrence (f) | Percentage of occurrence in relevant snippets subset $\left( \frac{f}{N-n} \right)\times100$ | Percentage of occurrence in the document $\left( \frac{f}{N} \right)\times100$ |
| --- | --- | --- | --- | --- |
| Suicidal attempt | | 578 | 27.02 | 13.24 |
| Suicidal ideation | Passive suicidal ideation | 87 | 4.07 | 1.99 |
|  | Active suicidal ideation | 528 | 24.68 | 12.1 |
| Non-suicidal snippets | | 373 | 17.44 | 8.55 |
| Self-injurious behavior (NSSI) | | 59 | 2.76 | 1.35 |
| Mental disorder | | 418 | 19.54 | 9.58 |
| Overdose | | 529 | 24.73 | 12.12 |

- n= 2225
- N= 4364
- It is important to note that patients could have multiple annotations with different labels, which were not mutually exclusive with the exception that irrelevant snippets cannot be assigned any other labels during the annotation phase since only the snippets which provide no relevant information to the suicide topic will be annotated as irrelevant.
