## Supplementary material for "A Natural Language Processing Algorithm for Classifying Suicidal Behaviors in Alzheimer’s Disease and Related Dementia Patients: Development and Validation Using Electronic Health Records Data": Table 1

Table 1: Key properties (labels) and definitions for suicidal behavior used for the annotation of clinical notes.

| Labels | Definitions |
| --- | --- |
| Suicidal attempt | A potentially self-harming act, indicating at least some intention to cause death consequently. The evidence of the individual's intent to end their own life, to some extent, can be either explicit or deduced from their behavior or situation. A suicide attempt may or may not lead to actual harm^1^. |
| Passive suicidal ideation | This refers to a desire to be dead. It involves thoughts of not wanting to be alive anymore or wishing to fall asleep and not wake up^1^. |
| Active suicidal ideation | Active suicidal ideation is nonspecific and does not involve a specific method, intent, or plan. It entails general thoughts of wanting to end one's life or commit suicide, such as expressing thoughts like "I've thought about killing myself". However, there are no accompanying thoughts regarding specific methods, intent, or a plan during the assessment period^1^. |
| Self-injurious behaviors (non-suicidal self-injury (NSSI) | Self-injurious behavior without any intention to cause death is performed for reasons other than ending one's life. This behavior serves different purposes, such as relieving distress (referred to as self-mutilation) through actions like superficial cuts, scratches, hitting, banging, or burns. Additionally, it may also be aimed at influencing others or the environment to bring about a change^1^. |
| Non- suicidal snippets | In the absence of suicidal ideation or self-injurious behavior, it can be inferred that the patient is non-suicidal. To annotate a snippet as non-suicidal, we looked for a strong indication of negation of suicidal ideation and self-injurious behavior^1^. |
| Mental disorders | Mental disorders are characterized by significant disruptions in cognition, emotion, or behavior, often causing distress and impairing functioning. They are part of a broader category known as mental health conditions, which includes psychosocial disabilities and states involving distress, impaired functioning, or risk of self-harm^2^. |
| Overdose | An overdose occurs when an individual consumes an excessive amount of a substance, often drugs, surpassing the recommended dosage^3^. It can lead to severe symptoms or even death. Distinguishing intentional (suicidal) overdoses from unintentional ones is challenging due to incomplete information and overlapping features. Our annotation approach focuses on categorizing overdoses without explicitly labeling them as either suicidal or unintentional^4^. |
| Irrelevant | The "Irrelevant" category in our annotation refers to clinical notes that are unrelated to suicide or any defined categories. These notes cover different medical topics, treatments, or procedures, not relevant to the study's focus. By excluding irrelevant notes, we maintain the specificity and accuracy in analyzing suicidal behavior, mental health, and related aspects of interest. Annotators ensured consistent labeling to ensure the quality of data used for training and evaluation. |

^1^- Guidance for Industry: Suicidal Ideation and Behavior: Prospective Assessment of Occurrence in Clinical Trials. Clinical Trials 2012

^2^- Mental disorders. https://www.who.int/news-room/fact-sheets/detail/mental-disorders

^3^- Overdose: MedlinePlus Medical Encyclopedia. https://medlineplus.gov/ency/article/007287.htm

^4^- Stover AN, Rockett IRH, Smith GS, *et al.* Distinguishing clinical factors associated with unintentional overdose, suicidal ideation, and attempted suicide among opioid use disorder in-patients. *J Psychiatr Res* 2022;**153**:245–53. doi:10.1016/j.jpsychires.2022.06.039
